# Community awareness, access, and experiences of cervical screening following universal access to self-collection in Australia

**DOI:** 10.64898/2026.05.12.26353060

**Authors:** Chloe J. Jennett, Claire Bavor, Tessa Saunders, Lisa J. Whop, Louise Mitchell, Karen Canfell, Natalie Taylor, Louiza S. Velentzis, Sam Egger, Julia M.L Brotherton, Claire E. Nightingale, Megan A. Smith

## Abstract

**Background:** Since July 2022, Australian National Cervical Screening Guidelines have recommended anyone eligible for cervical screening be offered the choice between having their sample collected by a clinician with a speculum, or self-collection using a vaginal swab.

**Method:** We recruited screen-eligible people to an online survey between December 2023 and April 2024, via a paid social media (Meta) campaign, and stakeholder and community networks.

Using binary logistic regression, we assessed demographic and screening history factors associated with having previously heard of self-collection. In participants screened since July-2022, we assessed factors associated with being offered a choice between self-collection and clinician-collection; choosing self-collection (among those offered choice); and using self-collection (among all recently screened participants).

**Results:** Of the 9,928 participants, 70.2% had heard of self-collection. Among those screened since July 2022, 36.1% were offered a choice in screening method.

Awareness was associated with increasing age (p-trend **<0.001**), with participants aged >65 years most likely to have heard of self-collection (adjusted odds ratio (aOR): 1.69, 95% confidence interval (95%CI): 1.31-2.18).

Compared to participants who self-reported regularly attending cervical screening, both not-regular and never screeners (based on self-reported screening history, frequency, age and sexual history) were less likely to have heard of self-collection (aOR:0.80 [95%CI:0.72-0.89] and aOR:0.73 [95%CI:0.56-0.96], respectively; p***<0.001***). Participants who attended a specialised women’s/sexual health clinic were more likely to have heard of self-collection (aOR:1.32 (95%CI:1.06-1.64), p;***<0.001***), and to report being offered choice (aOR:1.62 (95%CI 1.22-2.14), p***<0.001***) at their last cervical screen.

Half of the participants who were offered a choice opted for self-collection (N=803/1,617; 49.7%). Not-regular screeners were twice as likely (aOR:2.31 (95%CI:1.74-3.07), p***<0.001***) to choose self-collection as regular-screeners.

**Conclusion:** Given almost 50% of women nationally are now choosing self-collection, these findings imply national uptake might be close to plateauing overall. In high income settings where a choice in screening methods is introduced with the aim of improving screening equity, resources, adequate training, and health promotion tools should be provided prior to program launch to support healthcare providers in offering choice and facilitate improved participation in screening programs. Raising community awareness of screening options is important and needs to reach under-screened groups.

## Introduction

Annual rates of cervical cancer in Australia are among the lowest in the world (1). Nationally, cervical cancer incidence was 6.4/100,000 women in 2020, and mortality was 1.3/100,000 women in 2018-2022 (1). This is largely due to the establishment of the National Cervical Screening Program (NCSP) in 1991, initially recommending 2-yearly cytology testing, and transitioning to 5-yearly primary HPV testing from December 2017(2). Immunisation against human papillomavirus (HPV), introduced to Australia’s National Immunisation Program in 2007(3), is anticipated to further reduce cancer incidence in future. This is supported by a decreasing trend in the detection of vaccine-targeted HPV types among vaccine-eligible age cohorts(1,4).

In Australia, women and people with a cervix aged 25–74 are invited by the National Cancer Screening Register (NCSR) for cervical screening every 5 years as part of the NCSP (5). The test is offered mainly through primary care/general practice and is largely reimbursable under Australia’s universal health coverage, Medicare. However, there can be out-of-pocket costs associated with the healthcare provider visit. Despite the longstanding cervical screening program, inequities in participation are evident, with disparities in both screening coverage and cancer incidence among Aboriginal and Torres Strait Islander women, by socioeconomic status and by geographic remoteness of residence (1,4,6).

Within the previous cytology-based program, never-screened or under-screened women made up approximately 70% of all cervical cancer cases among those of screening age (7). Studies have suggested a wide range of barriers to participation in organised screening programs, including practical factors, health systems, personal or cultural values, knowledge, fear, and embarrassment (8).

One innovation enabled by primary HPV testing is the ability to implement self-collection for cervical screening. Self-collection involves taking a lower vaginal sample using a swab, without a speculum, for HPV testing. It is an alternative to a clinician-collected cervical sample, which requires a speculum exam. Self-collection overcomes many barriers to cervical screening, and evidence suggests it is highly acceptable (9,10). Self-collection has been available to never- and under-screened people in Australia since early 2018, with the aim of increasing screening participation (11). Uptake during this time was low, likely due to limited laboratory availability to process samples, minimal promotion, lack of confidence among providers and difficulty in confirming participants’ history to assess eligibility (12). Strong evidence exists showing equivalent sensitivity between self-collection and clinician-collection (13). In response, Australian guidelines were updated in July 2022 to recommend that anyone eligible for screening be offered the choice to ‘use self-collection’ or clinician-collection (with the new recommendation referred to as ‘universal access’)(5).

Research assessing awareness and adoption of self-collection in Australia has been limited to pilot and sub-population studies largely prior to the policy change, and there is little evidence on whether screen-eligible people are consistently being offered a choice in screening method (14–16). Australia is one of the first countries to adopt a model of offering a choice between self-collection or clinician-collection. International evidence predicts the majority (69%) of women would opt for self-collection at home if provided a choice however actual uptake remains unknown (17).

Given this knowledge gap, the novel way self-collection is offered in Australia, and to further support the implementation of self-collection in the NCSP, we conducted a nation-wide survey of screen-eligible people. We aimed to assess awareness of self-collection, examine whether recently screened participants were offered a choice in screening method, whether they ‘used self-collection’ (among all recently screened), and ‘chose self-collection’ (among participants offered choice). We also report on general screening experiences and participants’ preferences for either self- or clinician-collection in the future.

## Methodology

Data analysed for this study were obtained from a cross-sectional, population-based electronic survey. The survey consisted of six sections; 1) information and consent; 2) demographics; 3) prior screening history; 4) experiences at their last cervical screen (participants screened since universal access); 5) future screening preferences; and 6) end of survey information and prize draw (supplementary file 1). Questions about gender, sex, intersex, and sexuality were developed in accordance with community indicators (18). The survey was hosted on Qualtrics and included closed and open questions informed by the COM-B model and Theoretical Domains Framework (19,20). The survey was piloted by consumers, taking approximately 15 minutes to complete.

### Recruitment

The survey was circulated across multiple channels: a 4-week Meta campaign (Instagram, Facebook), local stakeholder groups, and hard-copy flyers. Recruitment was open between December 2023 and April 2024 to women and people with a cervix aged 24-74 years of age living in Australia. The recruitment channels and materials were revised throughout, in response to monitoring of key demographic factors (age, state), with the aim of recruiting a sample representative of the eligible population, with over-sampling of Aboriginal and Torres Strait Islander people and people screened since July-2022.

Participants accessed the survey through an anonymous link. Informed consent was obtained via a checkbox prior to survey completion. Participants had the option to enter a draw to win 1 of 24 vouchers valued at 100 Australian dollars each.

### Sample selection and data cleaning

The sample included in the final analysis is shown in Figure 1, which outlines the process for assessing data quality and identifying potentially fraudulent responses (21,22). Data from Indigenous respondents, excluded from Figure 1, was analysed in line with Indigenist research methodology, and is reported elsewhere (23). Analyses involving future screening preference are also reported on separately(24).

**Figure 1:**
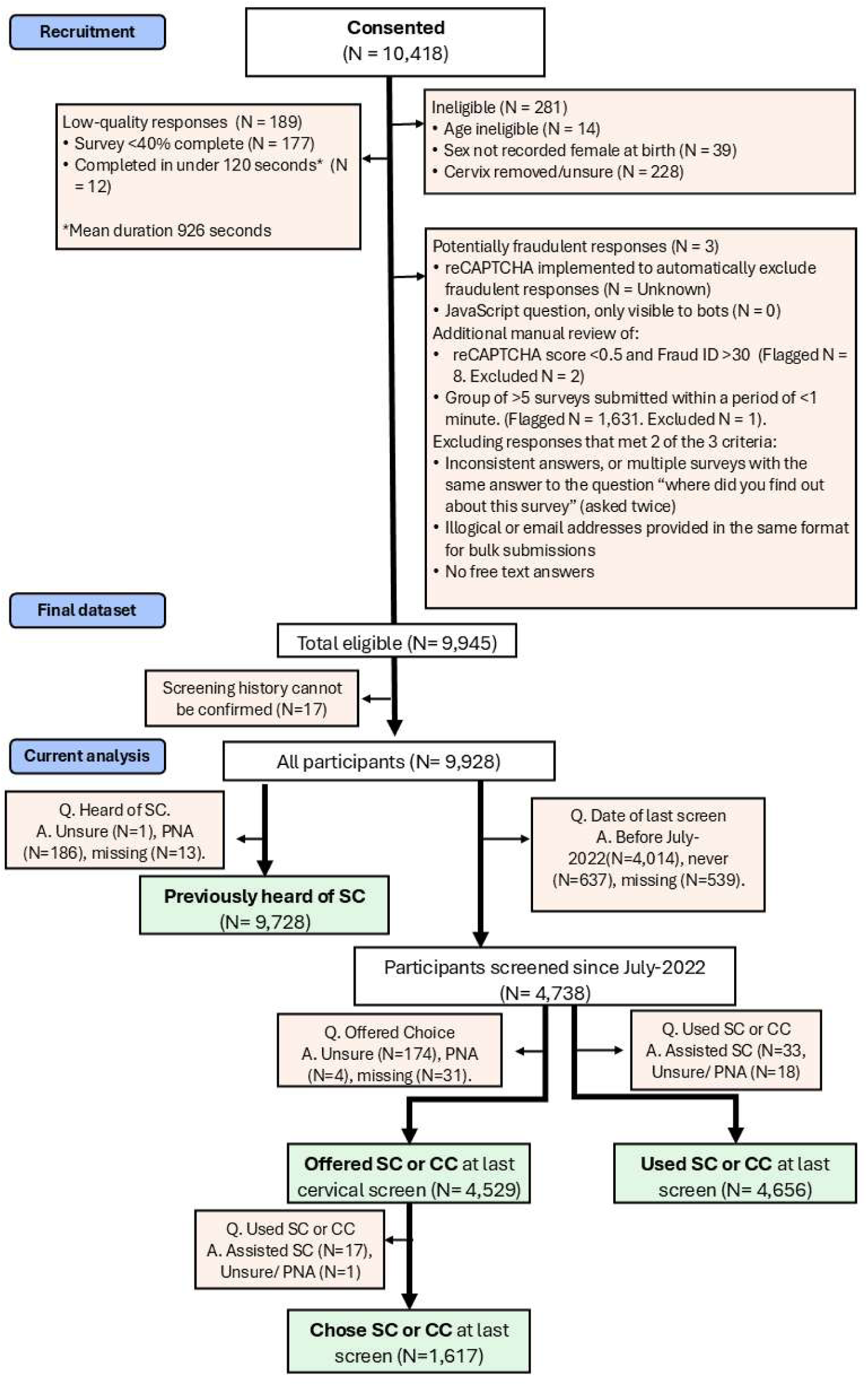
Recruitment and data cleaning process. **legend:** Abbreviations; SC; self-collection, CC; clinician-collection, PNA; Prefer not to answer, Q; question, A; answer. Orange shaded shapes indicate excluded responses, green shaded shapes indicate the number of participants included in key analyses. Where two arrows come out of a box, they are not mutually exclusive pathways.

### Statistical analysis

Data were analysed in STATA v18. Postcode was mapped to remoteness and area-level socioeconomic status using Accessibility/Remoteness Index of Australia Plus (ARIA+) 2021 and Socio-Economic Indexes for Areas (SEIFA) Index of Relative Socio-economic Disadvantage (IRSD) (25). Initially unmappable postcode data was manually reviewed against ABS data and updated/assigned where possible (26). Survey participants were categorised as regular screeners, not-regular screeners, or never-screened based on self-reported screening history, frequency, age and sexual history (supplementary Figure 1). Never-screeners were further stratified into those aged <27 (became eligible to screen relatively recently), those aged ≥27 (at least 2 years overdue for their first screen), and those who had never been sexually active. Participants for whom a screening history variable could not be created due to missing data were excluded from the analysis (Figure 1).

Supplementary Table 1 describes the primary outcome variables: awareness, offered choice in screening method, use of self-collection and chose self-collection, and how they were recoded for analysis. Binomial logistic regression was performed across all four primary outcomes. Demographic and prior screening history variables of interest were determined by the co-authors at the point of survey design based on prior research findings. After further discussion CJJ, MAS selected variables for each of the four primary outcomes for unadjusted analysis. The final adjusted regression for each outcome was determined based on evidence of an association (p-value <0.05) in the unadjusted model. Remoteness of residence was included in all adjusted models, as it is regarded as a key factor in participation in cervical screening in Australia (1). Where there was risk of multicollinearity, one variable was selected (investigator’s choice based on reasoning) to be carried forward to the final model. On this basis, SEIFA IRSD was excluded due to collinearity with remoteness. Missing data was coded as a category in each variable and included in the main analysis.

Sensitivity analyses using complete case data was conducted to assess whether results were similar to those obtained using missing indicators in the main adjusted regression analyses (supplementary Tables 3b – 6b).Sensitivity analysis was undertaken on the ‘chose self-collection’ and ‘used self-collection’ outcome variables excluding the screening history variable, to assess whether results were similar to those obtained in the main analysis, as screening history is potentially a mediator variable between, for example, remoteness of residence and desire to ‘use self-collection’ (supplementary tables 5a and 6a).

Descriptive statistics were used to summarise participants’ experience at their last cervical screening test (CST), including whether they received key information about the self-collection process, and their preferred sample collection method at their next CST.

## Results

10,418 people identified as non-Indigenous and consented to the study. Following review, 95% (9,945/10,418) of responses were included in the final dataset, and 9,928 reported on in this analysis (17 excluded from final dataset due to missing screening history data)

### Participant Characteristics

Table 1 describes the demographic and screening history variables for all 9,928 survey participants, alongside the 4,738 screened since universal access. Mean age of the total sample was 42.5 years (SD: 11.6), 87.6% were heterosexual, 63.9% lived in a major city, 82.5% were Australian born, and 61.4% reported having tertiary education. We determined 61.5% of participants to be regular screeners, 26.6% not-regular screeners, 2.6% never-screeners aged ≥27, 2.2% never-screeners aged <27 years, and 1.6% to be never-screeners who were never sexually active. Just over 5% of participants could not recall their last screen and were classified as ‘can’t remember’. The cohort screened after July-2022, when universal access came into effect, was broadly consistent in terms of demographic factors with the entire sample. Supplementary Table 2 includes demographics for all participants last screened prior to July-2022, those who were never screened, and those who could not remember.

**Table 1.** Demographic characteristics and prior screening experiences of all survey participants and those screened since universal access to self-collection.

|  | **Total**<br>**N (% within**<br>**category col)** | **Screened after July-22**<br>**n (% of row total)** |
| --- | --- | --- |
| **Total** | 9928 (100.0) | 4738 (47.7) |
| **Mean age (years) [SD]** | 42.53 [11.56] | 42.78 [11.74] |
| **Age group (years)** |  |  |
| 20-24 | 177 (1.8) | 41 (23.2) |
| 25-34 | 2631 (26.5) | 1312 (49.9) |
| 35-44 | 3202 (32.3) | 1470 (45.9) |
| 45-54 | 2203 (22.2) | 1057 (48.0) |
| 55-64 | 1288 (13.0) | 632 (49.1) |
| Over 65 | 427 (4.3) | 226 (52.9) |
| **Gender<sup>bc</sup>** |  |  |
| Cis | 9801 (98.7) | 4684 (47.8) |
| Trans | 118 (1.2) | 50 (42.4) |
| Not stated | 9 (0.1) | <5 (44.4) |
| **Intersex<sup>b</sup>** |  |  |
| No | 9823 (98.9) | 4692 (47.8) |
| Yes | 19 (0.2) | 11 (57.9) |
| PNA/Unsure | 74 (0.7) | 29 (39.2) |
| Missing data | 12 (0.1) | 6 (50.0) |
| **Sexuality<sup>b</sup>** |  |  |
| Heterosexual | 8692 (87.6) | 4190 (48.2) |
| Gay/Lesbian | 191 (1.9) | 64 (33.5) |
| Bisexual | 750 (7.6) | 358 (47.7) |
| I use a different term | 190 (1.9) | 78 (41.1) |
| Not stated | 104 (1.0) | 47 (45.2) |
| Missing data | <5 (0.0) | <5 (100.0) |
| **State** |  |  |
| ACT | 334 (3.4) | 154 (46.1) |
| NSW | 3107 (31.3) | 1433 (46.1) |
| NT | 181 (1.8) | 102 (56.4) |
| QLD | 2025 (20.4) | 950 (46.9) |
| SA | 571 (5.8) | 274 (48.0) |
| TAS | 344 (3.5) | 171 (49.7) |
| VIC | 2202 (22.2) | 1082 (49.1) |

|  | Total<br>N (% within<br>category col) | Screened after July-22<br>n (% of row total) |
| --- | --- | --- |
| WA | 1164 (11.7) | 572 (49.1) |
| **Remoteness of residence <sup>de</sup>** |  |  |
| Major city | 6346 (63.9) | 3028 (47.7) |
| Inner regional | 2015 (20.3) | 988 (49.0) |
| Outer regional/remote/very remote | 966 (9.7) | 440 (45.5) |
| Missing data | 601 (6.1) | 282 (46.9) |
| **SEIFA <sup>de</sup>** |  |  |
| 1 (most disadvantaged) | 1040 (10.5) | 470 (45.2) |
| 2 | 1695 (17.1) | 813 (48.0) |
| 3 | 2099 (21.1) | 996 (47.5) |
| 4 | 2271 (22.9) | 1054 (46.4) |
| 5 (most advantaged) | 2222 (22.4) | 1123 (50.5) |
| Missing data | 601 (6.1) | 282 (46.9) |
| **Country of birth** |  |  |
| Australia | 8195 (82.5) | 3897 (47.6) |
| Other | 1715 (17.3) | 834 (48.6) |
| Missing data | 18 (0.2) | 7 (38.9) |
| **Language spoken at home** |  |  |
| English | 9757 (98.3) | 4668 (47.8) |
| Other | 160 (1.6) | 67 (41.9) |
| PNA | 5 (0.1) | <5 (40.0) |
| Missing Data | 6 (0.1) | <5 (16.7) |
| **Duration of time in Australia** |  |  |
| Australian born | 8195 (82.5) | 3897 (47.6) |
| 0-4 years | 121 (1.2) | 54 (44.6) |
| 5-9 years | 182 (1.8) | 98 (53.8) |
| >10 years | 1380 (13.9) | 665 (48.2) |
| Missing data | 50 (0.5) | 24 (48.0) |
| **Highest education attained** |  |  |
| Tertiary | 6096 (61.4) | 2940 (48.2) |
| Tafe/certificate | 2551 (25.7) | 1194 (46.8) |
| School | 1233 (12.4) | 581 (47.1) |
| Other/PNA | 48 (0.5) | 23 (47.9) |
| **Prior screening history** |  |  |
| Regular screeners | 6110 (61.5) | 3715 (60.8) |
| Not-regular screeners | 2642 (26.6) | 1023 (38.7) |
| Never | 262 (2.6) | 0 (0.0) |
| Never <27yrs | 216 (2.2) | 0 (0.0) |
| Can't remember | 539 (5.4) | 0 (0.0) |
| Never sexually active | 159 (1.6) | 0 (0.0) |
| **Previously heard of SC** |  |  |
| No | 2898 (29.2) | 1053 (36.3) |
| Yes | 6830 (68.8) | 3636 (53.2) |
| PNA | 187 (1.9) | 44 (23.5) |
| Missing data | 13 (0.1) | 5 (38.5) |

### Primary Outcomes

Results for unadjusted and adjusted analyses are presented below. Results for variables included in the unadjusted regression model, including those which were not significantly associated with being offered a choice in screening method, prior awareness of self-collection, ‘used self-collection’ or ‘chose self-collection’ – and therefore excluded from the final adjusted regression for each outcome, are presented in supplementary Tables 3A to 6A.

For all outcomes (awareness, offer of choice, chose self-collection, and used self-collection) findings from the complete case analyses (supplementary Tables 3B to 6B) and findings from the sensitivity analyses removing prior screening history (chose self-collection, and used self-collection) (supplementary Tables 5A, 6A) were consistent with those from the primary analysis.

### Awareness of Self-Collection

Most participants reported having previously heard of self-collection (70.2%). Participants most frequently reported first hearing about self-collection through the media (36%) followed by a healthcare provider (28%) (supplementary Figure 2).

Although awareness was high across demographic subgroups, there were some significant differences (Table 2). There was a trend for increasing awareness with increasing categorical age group (p=<0.001, p-trend<0.001). When compared to the 25-29 age group, participants over 65 were significantly more likely to have heard of self-collection (AOR 1.69, 95%CI 1.31-2.18). ‘Awareness of self-collection’ was associated with the location where their last CST was conducted (p<0.001). Participants who attended cervical screening at a women’s/sexual health clinic, compared to a doctor’s clinic, were more likely to have heard of self-collection (AOR 1.32, 95% CI: 1.06-1.64). State of residence was also associated with prior awareness (p<0.001). Compared to participants living in the state of New South Wales, those in Western Australia were less likely to report prior awareness (AOR 0.86 95% CI: 0.74-0.99), whereas those in South Australia and Victoria (AOR 1.97, 95% CI: 1.57-2.47; AOR 1.54,95% CI: 1.35-1.75, respectively) were more likely. Language spoken at home was another significant factor (p=0.004), with participants who spoke a language other than English at home, less likely to be aware (AOR 0.57, 95% CI: 0.41-0.80). Finally, prior screening history (p=<0.001) and highest level of education attained (p=<0.001) were a significantly associated factor.

**Table 2.**
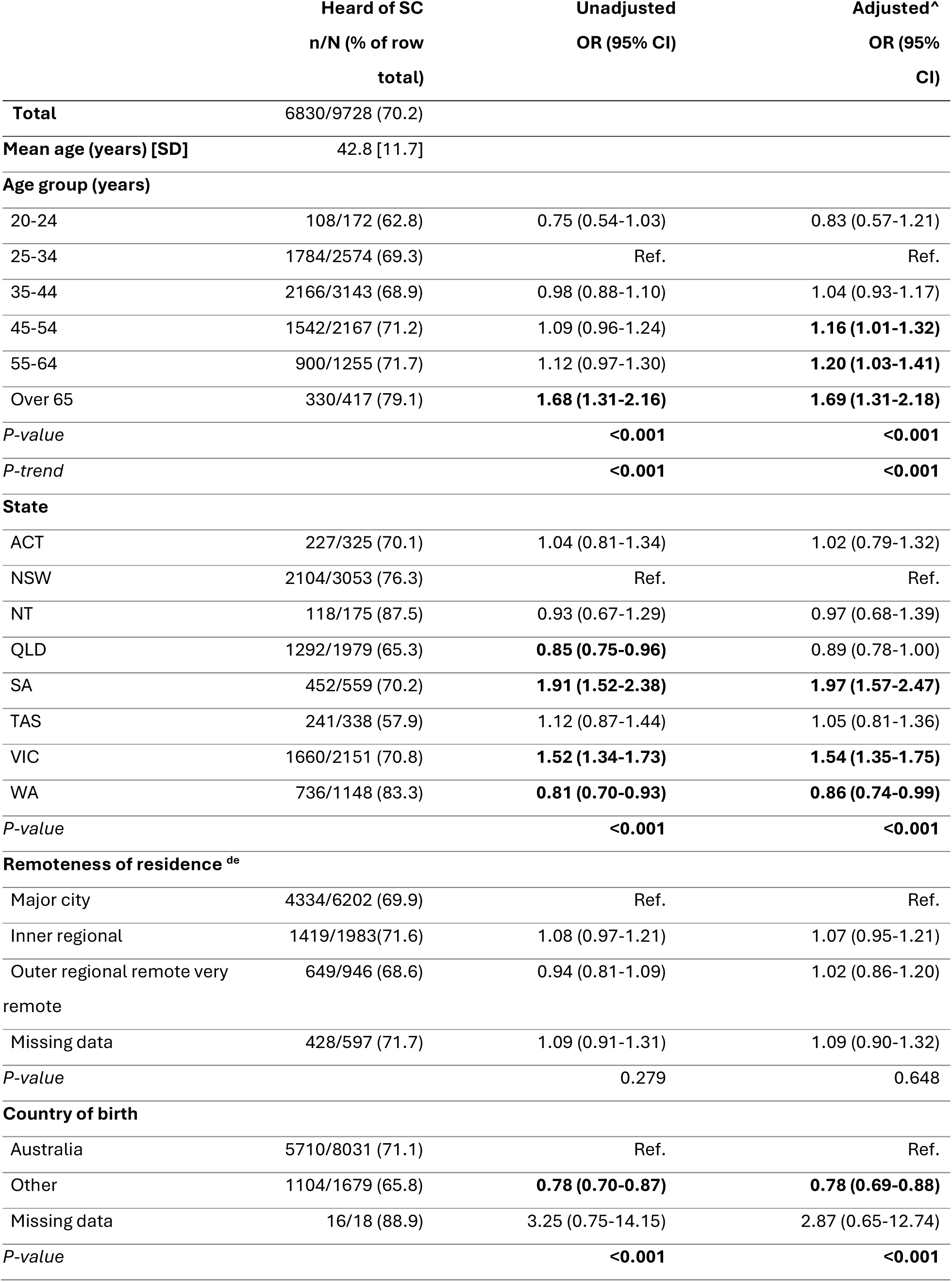
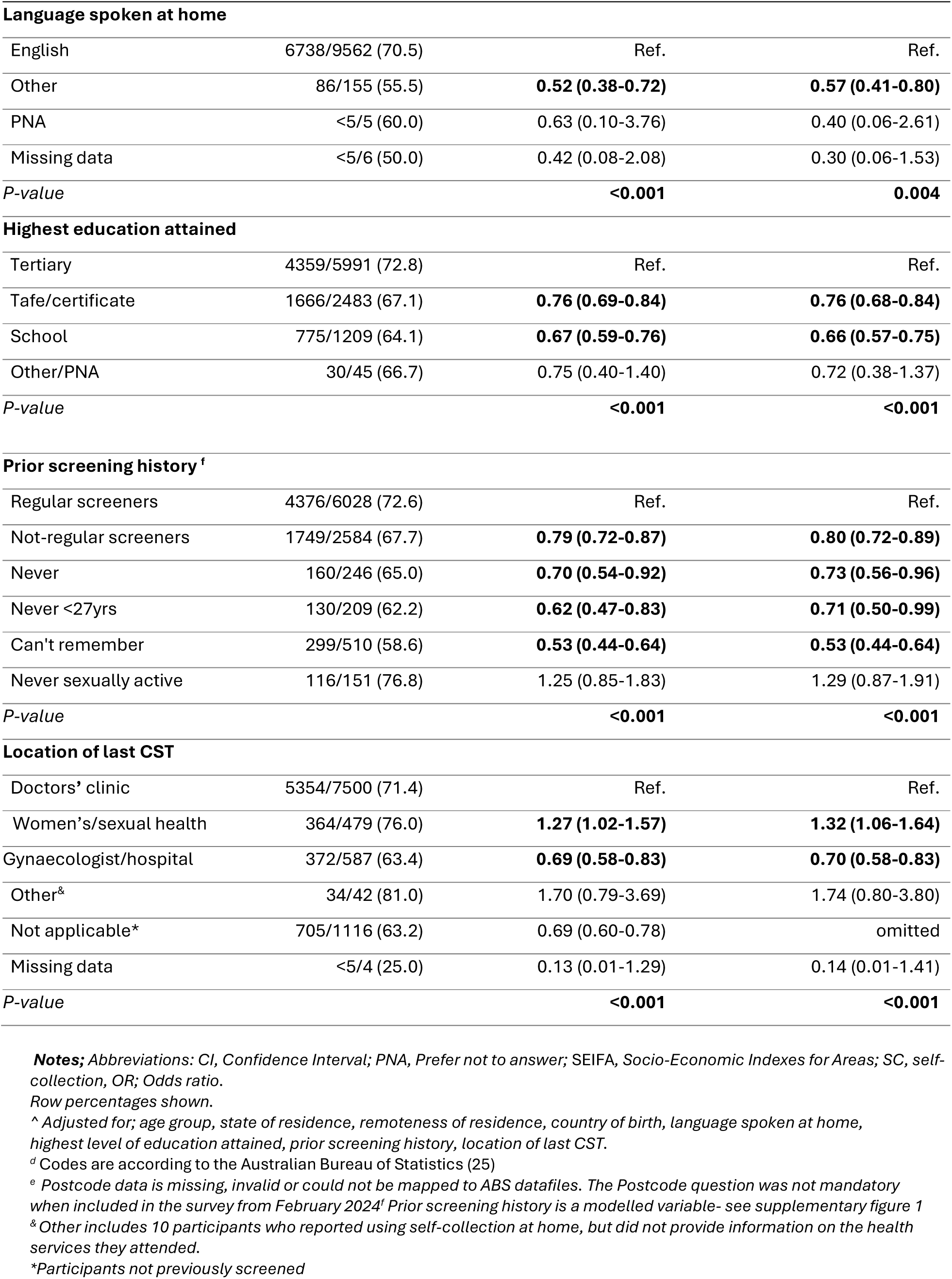
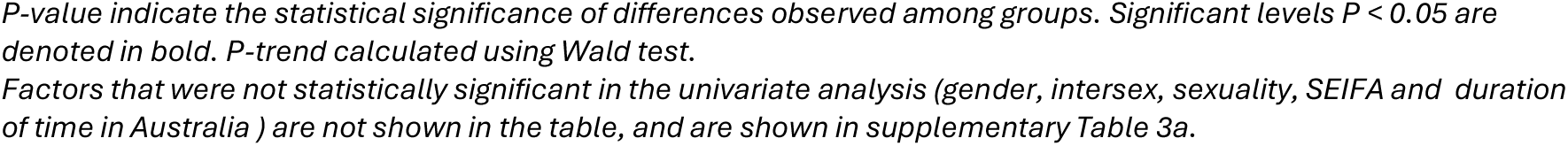
Demographic and screening history factors associated with prior awareness of self-collection.

### Offered Choice

Just over a third (36.1%) of participants who reported attending cervical screening since July-2022 recalled being offered a choice between self-collection and clinician-collection (Table 3).

**Table 3.** Demographic and screening history factors associated with being offered a choice between self-collection and clinician collection.

|  | Offered a choice<br>in screening<br>method n/N (%) | Unadjusted OR (95% CI) | Adjusted^<br>OR (95% CI) |
| --- | --- | --- | --- |
| **Total** | 1635/4529 (36.1) |  |  |
| **Mean age (years) [SD]** | 43.6 [12.2] |  |  |
| **Age group (years)** |  |  |  |
| 20-24 | 10/41 (24.4) | 0.59 (0.29-1.22) | 0.87 (0.40-1.88) |
| 25-34 | 439/1243 (35.3) | Ref. | Ref. |
| 35-44 | 468/1410 (33.2) | 0.91 (0.77-1.07) | **0.82 (0.69-0.97)** |
| 45-54 | 384/1014 (37.9) | 1.12 (0.94-1.33) | 0.92 (0.76-1.11) |
| 55-64 | 232/602 (38.5) | 1.15 (0.94-1.40) | 0.90 (0.72-1.12) |
| Over 65 | 102/219 (46.6) | **1.60 (1.19-2.13)** | 1.24 (0.91-1.70) |
| *P-value* |  | **<0.001** | 0.068 |
| *P-trend* |  | **<0.001** | 0.550 |
| **Sexuality<sup>b</sup>** |  |  |  |
| Heterosexual | 1448/4013 (36.1) | Ref. | Ref. |
| Gay/Lesbian | 28/60 (46.7) | 1.55 (0.93-2.58) | **1.72 (1.00-2.98)** |
| Bisexual | 114/339 (33.6) | 0.90 (0.71-1.13) | 0.96 (0.75-1.24) |
| I use a different term | 34/73 (46.6) | 1.54 (0.97-2.46) | 1.63 (0.99-2.69) |
| Not stated | 10/43 (23.3) | 0.54 (0.26-1.09) | 0.60 (0.29-1.27) |
| Missing Data | <5/<5 (100.0) | omitted. | omitted. |
| *P-value* |  | **0.038** | 0.050 |
| **State** |  |  |  |
| ACT | 50/145 (34.5) | 1.05 (0.74-1.51) | 1.06 (0.73-1.55) |
| NSW | 454/1364 (33.3) | Ref. | Ref. |
| NT | 42/100 (42.0) | 1.45 (0.96-2.19) | **1.71 (1.05-2.77)** |
| QLD | 297/909 (32.7) | 0.97 (0.81-1.16) | 1.05 (0.87-1.27) |
| SA | 92/258 (35.7) | 1.11 (0.84-1.47) | 1.18 (0.88-1.59) |
| TAS | 79/164 (48.2) | **1.86 (1.34-2.58)** | **1.51 (1.06-2.15)** |
| VIC | 433/1038 (41.7) | **1.43 (1.21-1.70)** | **1.49 (1.25-1.78)** |
| WA | 188/551 (34.1) | 1.04 (0.84-1.28) | 1.10 (0.88-1.37) |
| *P-value* |  | **<0.001** | **<0.001** |
| **Remoteness of**<br>**residence<sup>de</sup>** |  |  |  |
| Major city | 991/2879 (34.4) | Ref. | Ref. |
| Inner regional | 387/953(40.6) | **1.30 (1.12-1.51)** | **1.23 (1.04-1.46)** |
| Outer regional/remote/very remote | 160/429 (37.3) | 1.13 (0.92-1.40) | 1.03 (0.80-1.32) |
| Missing data | 97/268 (36.2) | 1.08 (0.83-1.40) | 1.09 (0.82-1.44) |
| *P-value* |  | **0.007** | 0.118 |
| **Duration of time in Australia** |  |  |  |
| Australian born | 1362/3731 (36.5) | Ref. | Ref. |
| 0-4 years | 8/49 (16.3) | **0.34 (0.16-0.73)** | **0.42 (0.19-0.91)** |
| 5-9 years | 23/95 (24.2) | **0.56 (0.35-0.89)** | 0.68 (0.42-1.13) |
| >10 years | 234/633 (37.0) | 1.02 (0.86-1.21) | 0.95 (0.79-1.15) |
| Missing Data | 8/21 (38.1) | 1.07 (0.44-2.59) | 1.02 (0.41-2.52) |
| *P-value* |  | **0.008** | 0.136 |
| **Location of last cervical screen** |  |  |  |
| Doctors' clinic | 1463/3917 (37.4) | Ref. | Ref. |
| Women's/Sexual health clinic | 111/236 (47.0) | **1.49 (1.14-1.94)** | **1.62 (1.22-2.14)** |
| Gynaecologist/Hospital | 45/351 (12.8) | **0.25 (0.18-0.34)** | **0.31 (0.23-0.44)** |
| Other<sup>a</sup> | 16/24 (66.7) | **3.35 (1.43-7.86)** | **2.72 (1.14-6.51)** |
| Missing Data | <5/<5 (0.0) | omitted | omitted |
| *P-value* |  | **<0.001** | **<0.001** |
| **Prompt for last CST** |  |  |  |
| Gynaecological appointment or symptomatic | 40/214 (18.7) | 0.29 (0.20-0.42) | **0.29 (0.20-0.43)** |
| Can't remember | <5/13 (23.1) | 0.38 (0.10-1.38) | 0.36 (0.10-1.35) |
| Follow-up for abnormal screen | 27/189 (14.3) | **0.21 (0.14-0.32)** | **0.25 (0.16-0.38)** |
| Health worker told me | 231/620 (37.3) | **0.75 (0.60-0.93)** | **0.76 (0.61-0.95)** |
| Other | 35/71 (49.3) | 1.22 (0.75-1.99) | 1.24 (0.75-2.05) |
| Remembered I was due | 187/521 (35.9) | **0.70 (0.56-0.89)** | **0.73 (0.58-0.92)** |
| Reminder from clinic | 229/574 (39.9) | 0.84 (0.67-1.04) | 0.83 (0.66-1.04) |
| Reminder letter from Register | 329/743 (44.3) | Ref. | Ref. |
| Saw something on media | 6/12 (50.0) | 1.26 (0.40-3.94) | 1.34 (0.41-4.36) |

|  | Offered a choice<br>in screening<br>method n/N (%) | Unadjusted OR (95% CI) | Adjusted<sup>^</sup><br>OR (95% CI) |
| --- | --- | --- | --- |
| Multiple- excluding<br>Gynaecological appointment<br>symptomatic or follow-up | 408/861 (47.4) | 1.13 (0.93-1.38) | 1.13 (0.92-1.38) |
| Multiple-including<br>Gynaecological appointment<br>symptomatic or follow-up | 140/711 (19.7) | **0.31 (0.24-0.39)** | **0.34 (0.26-0.43)** |
| *P-value* |  | **<0.001** | **<0.001** |

**Notes;** Abbreviations: CI, Confidence Interval; PNA, Prefer not to answer; SEIFA, Socio-Economic Indexes for Areas; SC, self-collection, OR; Odds ratio. Row percentages shown.
<sup>^</sup> Adjusted for; age group, sexuality, state of residence, remoteness of residence, duration of time in Australia, , location of last CST, prompt for last CST.
<sup>d</sup> Codes are according to the Australian Bureau of Statistics (25)
<sup>e</sup> Postcode data is missing, invalid or could not be mapped to ABS datafiles. The Postcode question was not mandatory when included in the survey from February 2024. & Other includes 10 participants who reported using self-collection at home, but did not provide information on the health services they attended.
*P-value indicate the statistical significance of differences observed among groups. Significant levels $P < 0.05$ are denoted in bold. P-trend calculated using Wald test.*
*Factors that were not statistically significant in the univariate analysis (gender, intersex, SEIFA, country of birth, language spoken at home, highest education attained, and prior screening history) are not shown in the table, and are shown in supplementary table 4a.*

In the adjusted model (Table 3), state of residence (p<0.001), location of last CST (p<0.001), and type of prompt received as a reminder for attending their last CST (p<0.001), were all significantly associated with being offered a choice in screening method. Participants who lived in the state of Victoria (AOR 1.49, 95% CI: 1.25-1.78), Tasmania (AOR 1.51, 95%CI: 1.06-2.15), the Northern Territory (AOR 1.71, 95%CI: 1.05-2.77), in an inner regional area (AOR 1.23, 95%CI: 1.04-1.46), or attended for screening at a women’s/sexual health clinic (AOR 1.62, 95% CI 1.22-2.14), were more likely to report being offered a choice in screening method.

In contrast, participants who attended a gynaecologist or hospital were significantly less likely to have been offered a choice (AOR 0.31, 95% CI: 0.23-0.44), as were those who had been living in Australia for less than 5 years (AOR 0.42, 95% CI: 0.19-0.91). In addition, compared to those who attended screening solely prompted by a reminder from the NCSR, participants were less likely to be offered a choice if their recent screen was prompted by a health worker (AOR 0.76, 95% CI: 0.61-0.95) or because they remembered they were due (AOR 0.73, 95%CI 0.58-0.92). Participants who were least likely to be offered a choice were those who attended as follow-up to a previous abnormal screen (AOR 0.25, 95% CI: 0.16-0.38), or attending for another gynaecological appointment or due to concern about symptoms (AOR 0.29, 95% CI: 0.20-0.43) (also see Supplementary Figure 3).

### Chose self-collection

Among the participants who were offered a choice of screening method, 50% chose to ‘use self-collection’ (Table 4). In the adjusted model, self-reported prior screening history, and prompt for last CST were significantly associated with choosing self-collection (p<0.001 in both instances). Non-regular screeners were significantly more likely to choose self-collection than regular screeners (AOR 2.31, 95% CI: 1.74-3.07). In contrast, compared to those who attended screening following a reminder from the NCSR, participants who were prompted to attend as part of another gynaecological appointment or due to symptoms (AOR 0.36, 95% CI: 0.17-0.76), or due to a previous abnormal test (AOR 0.17, 95% CI: 0.06-0.53) were less likely to have chosen self-collection even when offered.

**Table 4.** Demographic and screening history factors associated with choosing self-collection.

|  | 'Chose self-collection'<br>n (%) | Unadjusted OR<br>(95% CI) | Adjusted^ OR<br>(95% CI) |
| --- | --- | --- | --- |
| **Total** | 803/1617 (49.7) |  |  |
| **Meag age (years) [SD]** | 44.4 [12.1] |  |  |
| **Age group (years)** |  |  |  |
| 20-24 | <5/10 (40.0) | 0.83 (0.23-2.99) | 1.68 (0.42-6.78) |
| 25-34 | 193/434 (44.5) | Ref. | Ref. |
| 35-44 | 229/466 (49.1) | 1.21 (0.93-1.57) | 1.03 (0.78-1.36) |
| 45-54 | 200/380 (52.6) | **1.39 (1.05-1.83)** | 1.19 (0.89-1.59) |
| 55-64 | 127/228 (55.7) | **1.57 (1.14-2.17)** | 1.31 (0.93-1.85) |
| Over 65 | 50/99 (50.5) | 1.27 (0.82-1.97) | 1.02 (0.64-1.62) |
| *P-value* |  | 0.082 | 0.573 |
| *P-trend* |  | **0.007** | 0.248 |
| **Remoteness of residence <sup>de</sup>** |  |  |  |
| Major city | 471/979 (48.1) | Ref. | Ref. |
| Inner regional | 214/382 (55.9) | **1.37 (1.08-1.74)** | **1.30 (1.01-1.67)** |
| Outer regional/remote/very remote | 75/159 (47.2) | 0.96 (0.69-1.35) | 0.89 (0.62-1.28) |
| Missing data | 43/97 (44.9) | 0.85 (0.56-1.31) | 0.89 (0.56-1.36) |
| *P-value* |  | **0.035** | 0.105 |
| **Prior screening history<sup>f</sup>** |  |  |  |
| Regular screeners | 581/1279 (45.4) | Ref. | Ref. |
| Not-regular screeners | 222/338 (65.7) | **2.30 (1.79-2.95)** | **2.31 (1.74-3.07)** |
| *P-value* |  | **<0.001** | **<0.001** |
| **Location of last CST** |  |  |  |
| Doctors' clinic | 728/1446 (50.3) | Ref. | Ref. |
| Women's/Sexual health | 47/111 (42.3) | 0.72 (0.49-1.07) | 0.80 (0.53-1.21) |
| Gynaecologist/Hospital | 14/44 (31.8) | **0.46 (0.24-0.88)** | 0.63 (0.31-1.25) |
| Other<sup>g</sup> | 14/16 (87.5) | **6.90 (1.56-30.49)** | 4.45 (0.98-20.10) |
| *P-value* |  | **0.002** | 0.082 |
| **Prompt for last CST** |  |  |  |
| Gynaecological appointment or symptomatic | 12/40 (30.0) | **0.40 (0.20-0.81)** | **0.36 (0.17-0.76)** |
| Can't remember | <5/<5 (66.7) | 1.86 (0.17-20.69) | 2.06 (0.18-23.91) |
| Follow-up for abnormal screen | <5/27 (14.8) | **0.16 (0.05-0.48)** | **0.17 (0.06-0.53)** |
| Health worker told me | 148/229 (64.6) | **1.70 (1.20-2.40)** | 1.32 (0.92-1.91) |

|  | 'Chose self-collection'<br>n (%) | Unadjusted OR<br>(95% CI) | Adjusted^ OR<br>(95% CI) |
| --- | --- | --- | --- |
| Other | 23/35 (65.7) | 1.78 (0.86-3.70) | 1.05 (0.49-2.28) |
| Remembered I was due | 93/185 (50.3) | 0.94 (0.65-1.35) | 0.98 (0.68-1.42) |
| Reminder from clinic | 107/225 (47.6) | 0.84 (0.60-1.18) | 0.91 (0.64-1.29) |
| Reminder letter from Register | 169/326 (51.8) | Ref. | Ref. |
| Saw something on media | <5/5 (80.0) | 3.72 (0.41-33.61) | 2.43 (0.26-22.91) |
| Multiple- excluding<br>Gynaecological appointment<br>symptomatic or follow-up | 204/403 (50.6) | 0.95 (0.71-1.28) | 1.01 (0.75-1.36) |
| Multiple-including<br>Gynaecological appointment<br>symptomatic or follow-up | 37/139 (26.6) | **0.34 (0.22-0.52)** | **0.34 (0.22-0.54)** |
| *P-value* |  | **<0.001** | **<0.001** |

**Notes;** Abbreviations: CI, Confidence Interval; SEIFA, Socio-Economic Indexes for Areas; SC, self-collection, OR; Odds ratio. Row percentages shown
<sup>a</sup> Adjusted for; age group, remoteness of residence, prior screening history, location of last CST, prompt for last CST.
<sup>d</sup> Codes are according to the Australian Bureau of Statistics (25)
<sup>e</sup> Postcode data is missing, invalid or could not be mapped to ABS datafiles. The Postcode question was not mandatory when included in the survey from February 2024<sup>1</sup> Prior screening history is a modelled variable- see supplementary figure
& Other includes 10 participants who reported using self-collection at home, but did not provide information on the health services they attended
P-value indicate the statistical significance of differences observed among groups. Significant levels $P < 0.05$ are denoted in bold. P-trend calculated using Wald test.
Factors that were not statistically significant in the univariate analysis (gender, intersex, sexuality, state, SEIFA, country of birth, language spoken at home, duration of time in Australia, highest education attained, and prior screening history) are not shown in the table, and are shown in supplementary table 5a.

### Used self-collection

Overall, one fifth (21%) of all participants screened since July 2022 ‘used self-collection’ at their last cervical screen (Supplementary Table 6A). Amongst those who used clinician-collection, the most frequently reported reasons for doing so were because they had always had it done by a healthcare provider (43%), they were not offered self-collection (41%), and they wanted their healthcare provider to have a look (22%) (Supplementary Figure 4A). Among those who used SC, the top three reasons for doing so were that a healthcare provider suggested it as a good option (51%), it was less embarrassing (48%) and more convenient (43%) (Supplementary Figure 4B). In the adjusted model (Supplementary Table 6A) state of residence (p<0.001), remoteness (p=0.009), prior screening history (p<0.001), location of last CST (p<0.001) and prompt for last CST (p<0.001) were all significantly associated with using self-collection as the screening method.

### Self-collection experience at last CST and future screening preference

Among the 4,682 participants screened since July 2022 with known sampling method overall, a similar proportion reported that they would prefer clinician-collection, self-collection, or have no strong preference either way (30.6%, 28.1% and 29.2%, respectively), but preferences varied strongly by the sampling method used at their recent screen (Table 5). Regardless of sampling method at their recent screen, participants most frequently preferred to use the same sampling method at their next screen; however, this preference was much stronger for those who had used self-collection’ (67.4%) than those who had used clinician-collection (37.4%). For each sampling method group, approximately 20-30% stated they would be happy with either clinician-collection or self-collection next time. Very few participants who had used self-collection specifically wanted to use clinician-collection at their next test (5.3%)

**Table 5.** Future screening preferences by previous CST method.

| <u>Preference for next CST</u> | <u>Method for last CST</u> |  |  | <u>Total n (col %)</u> |
| --- | --- | --- | --- | --- |
|  | <u>Used clinician-collection n (%)</u> | <u>Used self-collection n (%)</u> | <u>Used assisted self-collection n (%)</u> |  |
| **Total (row %)** | **3,680 (78.6)** | **969 (20.6)** | **33 (0.7)** | **4,682 (100)** |
| **Collected by a healthcare provider using a speculum [i.e. clinician-collection]** | 1,370 (37.4) | 51 (5.3) | <5 (9.1) | 1,424 (30.6) |
| **Self-collection** | 653 (17.8) | 648 (67.4) | 9 (27.3) | 1,310 (28.1) |
| **Collected by a healthcare provider using a vaginal swab (but no speculum) [i.e. assisted self-collection]** | 399 (10.9) | 19 (2.0) | 14 (42.4) | 432 (9.3) |
| **I would be okay with either** | 1,117 (30.5) | 234 (24.3) | 7 (21.2) | 1,358 (29.2) |
| **Unsure** | 111 (3.0) | 6 (0.6) | - | 117 (2.5) |
| **I am not planning to screen** | 11 (0.3) | <5 (0.4) | - | 15 (0.3) |

**Notes;** N= all participants screened since July 2022 who provided a valid answer to 'method for last CST' and 'preference for next CST'. Numbers differ to Table 5 as participants who 'used self-collection' supported were excluded from regression analysis, and participants are only included if they answered the future preference question.

Amongst participants who used self-collection at their last CST, a large majority said they had enough information to decide between self-collection and clinician-collection (86.2%), understood how to collect the sample (94.8%), and were confident the test was accurate (77.2%) (Supplementary Figure 5 and 6). However, a significant minority did not recall being informed of how they would receive their test result (36.7%) or if they were told they could ask for assistance with self-collection if they needed to (36.5%). In addition, only 78.7% were informed they might need to come back for another test if HPV was detected.

## Discussion

Our national survey of over 9,000 participants provides the first data on awareness, offer of choice and choosing self-collection among screen-eligible people following the introduction of universal access in Australia in July 2022. We found 70% had previously heard of SC. Awareness was highest in older-age groups, including post-menopausal age ranges, who often find speculum insertion painful, making self-collection a particularly attractive option. It also varied by state/territory and prior screening history with lower awareness among never or not-regular screeners. Among participants who had attended screening since July 2022, over a third (36.1%) reported being offered a choice between self-collection and clinician-collection. Participants who lived in Victoria, Tasmania, the Northern Territory, or inner-regional areas, or who attended for screening at a women’s/sexual health clinic, were more likely to report being offered a choice. Our survey also found that 50% of all participants who were offered choice, chose self-collection and that non-regular screeners were significantly more likely to choose self-collection. Among survey participants screened since July 2022, whether offered choice or not, one fifth (21%) used self-collection at their last cervical screen. Use varied by state/territory, remoteness and prior screening history. Participants who used self-collection reported high acceptability overall of the information provided to them at their last screen, and when asked about future screening preferences, there was a strong preference to use self-collection again.

In 2026, Australia is one of a relatively small but increasing number of countries offering self-collection for routine cervical screening, not only to those who are overdue. The most recent 2022 systematic review reported 9 countries offered it to all women in their national programs (28), with variation in approach and uptake. In comparable settings with established screening programs, the Netherlands and Sweden offer self-collection as an ‘opt-in’ option through mailout (28–30), with the Netherlands additionally using an ‘opt-out’ approach for newly screen-eligible people (31). More recently, New Zealand introduced a universal option to use self-collection when attending for screening, concurrent with transitioning to HPV screening, including take-home tests at the discretion of a healthcare provider (32). The Australian setting also recommends that choice is offered at a clinic visit for cervical screening but guidelines enable out-of-clinic approaches facilitated by a HCP(33).

Our study presents the first national large-scale findings of consumer experiences with cervical screening in the context of a universal option to use self- or clinician-collection. Limited Australian data exist on self-collection awareness to consider whether this has changed since eligibility expanded, however, our data shows a substantial improvement in Victoria compared to survey data collected in early-2022 (prior to universal access). At that time, 9.7% of the 700 Victorians surveyed were aware of self-collection, compared to 70.8% in Victoria at the time of our survey (34). Our findings regarding offer of self-collection are consistent with findings from interviews with 43 recently screened people living in Victoria, conducted around the time of this survey, that found 44% of participants were offered a choice at their most recent screen, compared to 41.7% of Victorian respondents in our survey (16). In our analysis, prior screening attendance was not correlated with the offer of choice, consistent with updated guidelines that previous screening participation should not determine access to self-collection. However, as only one third of participants reported being offered a choice, the guideline to offer all screen-eligible individuals a choice of screening method was clearly not being followed consistently at the time of this survey. Recent studies suggest ongoing barriers include an outdated understanding of self-collection accuracy and pessimistic views about the ability of patients to perform self-collection amongst clinicians (35).

In our study, 50% of survey participants who had been offered choice had chosen self-collection. This is substantially less than the 69% predicted in an English survey that asked about future preferences, however the option queried was self-collection carried out at home (17). While use of self-collection at home is possible under the current Australian guidelines following an appointment with a HCP, it is far from standard practice (33). In our survey, use, defined as the percentage of all participants screened since July 2022 who used self-collection, was 21%, comparable with use reported from national registry data in the period when our survey participants were screened, which increased from 10% in July to September 2022, to 30% by January to March 2023, near the time our survey ended. Low use in our survey reflects the low rate of survey participants who reported being offered choice. National data on the proportion of participants using self-collection has continued to increase, reaching 46% in April to June 2025 (36). This is comparable with the proportion of participants in our survey who ‘chose self-collection’ when offered, potentially suggesting that the proportion being offered choice is increasing. Several factors identified as associated with use of self-collection in our survey are also consistent with national registry data, with reported use being highest among people aged 70-74 years, who are under- or never-screened, living in Tasmania, the most disadvantaged SEIFA quintiles and in very remote areas(12). Likewise, our study identified self-collection use to be highest among Tasmanian and Victorian participants, residents in inner regional areas, and non-regular screeners. Differences in how self-collection is accessed internationally make global comparison difficult, however, Australia’s registry data indicates that current uptake sits at a midpoint in the global context. In New Zealand, where it is offered in a comparable way, self-collection made up 80% of all screens in the first year it was offered (32,37), while in the Netherlands, where kits are mailed out to all newly screen-eligible people, uptake is 22.2% (31)

Our study offers reflections of the implementation of universal access, and key recommendations for future action and research. ‘Awareness of self-collection’ was significantly lower among never or not-regular screeners than in routine screeners. This underscores a critical communication gap with the highest-priority group, those under or unscreened, which must be addressed to improve equity within the NCSP. State differences in awareness of self-collection may be partly explained by differences in state-led awareness activities during the initial implementation of universal access and who they were directed to. Both states where awareness was higher (South Australia, Victoria) launched campaigns to improve HCP knowledge and confidence in offering self-collection in 2022, with Victoria leading this initiative prior to universal access (38–40). In contrast, community awareness campaigns were run in late 2022 and early 2023 in 4 states, including both states with higher awareness and also those with lower awareness (WA) (41,42). Coordinated statewide campaigns were not conducted in the remaining 4 states and territories, though in some cases localised advocacy occurred. Since this survey was conducted, national government-funded campaigns have been run (43,44). A HCP campaign began in June 2024 aiming to address knowledge of universal access and improve confidence in the accuracy of self-collection, followed by a community campaign to improve awareness from September 2024, with additional focus on groups known to be under-screened (43,44). The community campaign used social and print media, and public transport signage (43). Campaign evaluations are still to be published; however, our findings regarding most frequently selected sources for having heard about self-collection, traditional media, HCP, and social media, suggest the communication avenues used in the national campaigns will be effective, and this is supported by prior qualitative research (45).

Our data shows, regardless of screening method used, participants’ reasoning is often centred around the influence of the HCP. Participants who used clinician-collection frequently stated they have always had it done by their provider or their HCP did not tell them about self-collection. Those who ‘used self-collection’ reflected that their HCP presented self-collection as a good alternative option for them. The potential for universal access to positively impact participation rates in the NCSP, is entirely reliant on HCPs to, firstly, offer a choice, and secondly oversee the screening process. While HCP support in offering self-collection has been documented to be high among certain samples (46), only a third of our survey respondents reported being granted this choice, and hence it remains unclear how the offer is communicated, and whether current methods are engaging individuals of different demographics and prior screening experiences in a way that effectively communicates autonomy of choice. The variation in self-collection use and choice in our survey, across states and territories suggests differences in how HCPs communicate with consumers, and may reflect disparities in the support they receive from laboratories and networks (12). Self-collection information needs among screen-eligible people have been previously assessed at a sub-population level(47–49). However, the extent to which resources meet current needs should be a future research priority at a national level, particularly given our findings regarding the significant minority of participants not recalling being told they could ask for assistance to perform self-collection, how they would receive their result or that they might need to come back for follow-up.

Notably, a critical step prior to being offered choice in cervical screening is the ability to first access a provider, with 26% of Australians who required a general practitioner visit in 2024/2025 reporting delaying their appointment, and 10% of females reporting this delay was due to financial reasons (50). The requirement to attend a HCP to participate in cervical screening is likely an inadvertent but ongoing barrier to an equitable program. A systematic review of strategies for increasing participation using self-collection indicate opt-in strategies to be less effective than send-to-all strategies (10). More flexible models of accessing self-collection kits, via a range of non-appointment based models, including mail out, pharmacy collection, integration with other mobile services and community based approaches, have been shown to be acceptable in Australia (24), and should be explored and evaluated. A range of non-medical providers could potentially be more involved, as in other countries and consistent with a national medical workforce review (51)

### Strengths and limitations

Our survey recruited a large sample, which is broadly representative of the population eligible for screening, due to our agile recruitment method that required considerable monitoring of multiple demographic variables (4). Our primary recruitment method of a 4-week national Meta campaign minimised bot responses, and was cost effective, with a per survey completion cost of approximately 1.57 Australian dollars. Some groups are under-represented in our sample; notably 17.3% of survey participants were born outside Australia, compared to 31.5% in national data (52). While the survey was developed using plain language principles, it was only available in English. This survey is part of an overarching program of work, ‘Supporting Choice’, that also includes complementary qualitative work focussed on priority populations, including culturally diverse communities (53).

## Conclusion

Improving participation in the NCSP is essential for Australia to achieve its goal to eliminate cervical cancer as a public health problem by 2035 (54). Our findings support that self-collection is highly acceptable (though not universally preferred) in Australia, but at the time of this survey, choice was not being consistently offered, and non-regular screeners were less aware it was an option. Given the critical role of HCPs in both offering choice and the choice made, timely and adequate training, resources and health promotion tools, need to be provided to support them in offering choice and to avoid missed opportunities in the early stages of policy change. These need to be complemented by campaigns to raise community awareness. National community and HCP campaigns have since been run in Australia, and the effects of these will be examined in our follow-up survey conducted in late 2025.

## Data Availability

The datasets generated and/or analysed during the current study are not publicly available due to ethics requirements for this study, but are available from the corresponding author on reasonable request.

## List of abbreviations

CST: Cervical screening test
HCP: Healthcare provider
HPV: Human papillomavirus
NCSP: National Cervical Screening Program
NCSR: National Cancer Screening Register

## Declarations

### Ethics approval and consent to participant

Ethics approval was obtained from the Australian Institute of Aboriginal and Torres Strait Islander Studies (REC-0209) and ACON (202404). Informed consent was obtained from all participants prior to survey completion. The study was conducted in accordance with the principles of Good Clinical Practice and the Declaration of Helsinki.

### Consent for publication

Not applicable

### Competing interests

CJJ: None to declare CB: None to declare

TS: None to declare LJW: None to declare

LM: None to declare

KC: co-PI on a major implementation program ‘Elimination Partnership for Cervical Cancer in the Indo-Pacific’ which receives support from the Australian government, the Minderoo Foundation and equipment donations from Cepheid Inc. KC receive contract funding through her work institution from the Department of Health, Disability and Ageing, Australia to monitor the safety of the National Cervical Screening Program. KC is co-PI of an investigator-initiated trial of HPV screening in Australia (’Compass’), which is conducted by the ACPCC, which has previously received equipment and a funding contribution for the Compass trial from the Australian government, Roche Molecular Systems USA and Micobix. KC is a chair or member of a number of government or meetings convened by the World Health Organization (WHO), or philanthropic organisations such as Bill and Melinda Gates Foundation (BMGF). She is also a chair of the Expert Advisory Group to the Elimination Response for Australian Government, and Cancer Screening and Immunization Committee of Cancer Council Australia.

NT: None to declare LSV: None to declare SE: None to declare

JMLB: Principal investigator of a GACD/NHMRC funded self-collection study in India (SHE-CAN) which has received donated HPV tests and swabs from Copan, Abbott and Seegene.

CEN: None to declare

MAS: reports receipt of contract funding to her institution from the Australian Government Department of Health, Disability and Ageing and the Australian Centre for the Prevention of Cervical Cancer (ACPCC), a government-funded, not-for-profit, health promotion charity, for separate projects related to the NCSP

### Author contributions

CJJ: methodology, data curation, investigation, writing–original draft, writing–review and editing, formal analysis, project administration, funding acquisition.

CB: methodology, writing- review and editing

TS: methodology, writing- review and editing

LJW: conceptualization, methodology, supervision, writing- review and editing, funding acquisition.

LM: methodology, data curation, writing- review and editing

KC: methodology, supervision, writing- review and editing

NT: methodology, supervision, writing- review and editing

LSV: supervision, writing- review and editing

SE: methodology, writing- review and editing

JMLB: methodology, writing- review and editing, funding acquisition.

CEN: Conceptualization, methodology, supervision, writing- review and editing, funding acquisition.

MAS: Conceptualization, methodology, supervision, writing- review and editing, funding acquisition.

All authors have approved the submitted version and have agreed both to be personally accountable for the author’s own contributions and to ensure that questions related to the accuracy or integrity of any part of the work, even ones in which the author was not personally involved, are appropriately investigated, resolved, and the resolution documented in the literature.

## Acknowledgements

We thank Josephine Mondino, Cancer Council Australia, for providing support and advice for the Meta recruitment campaign and ACON for assisting with the recruitment of participants. We thank the Aboriginal and Torres Strait Islander Advisory Group Thittu Tharrmay, Supporting Choice Consumer and Community Advisory Panel, Provider and Implementation Advisory Group and Investigator team for their ongoing guidance. Finally, we thank all the participants who took the time to complete this survey.

