## Supplementary Data for "Community awareness, access, and experiences of cervical screening following universal access to self-collection in Australia"

### Supplementary File 1 of 1

### Supplementary Figures

#### Supplementary Figure 1: Derived Screening history categories for regression analysis.

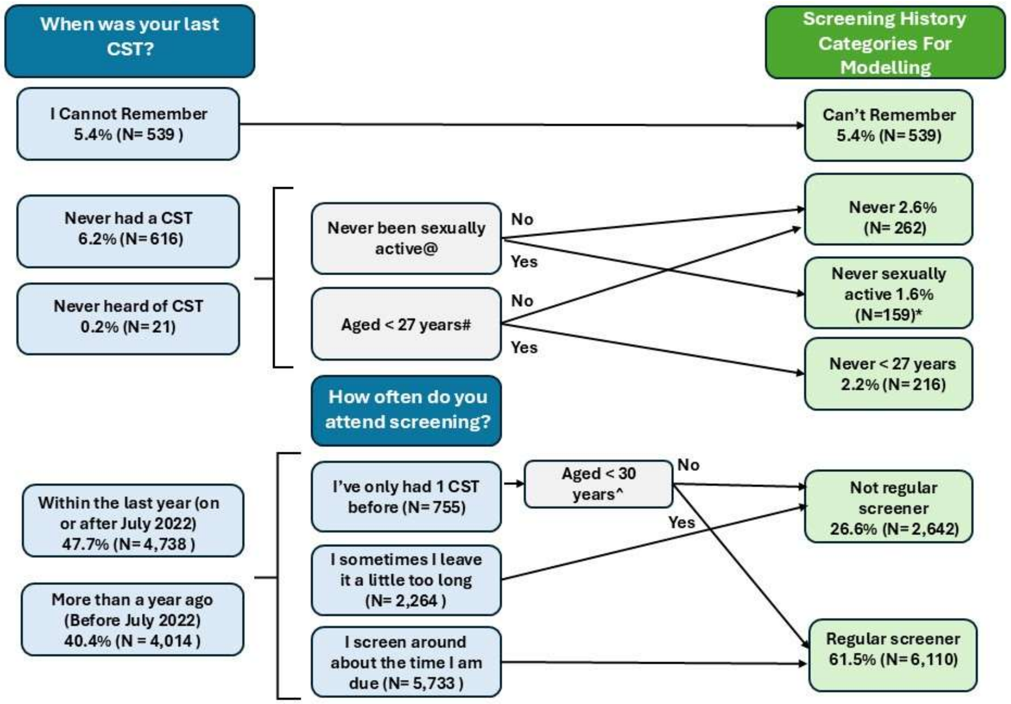

**Notes:** N=9,928 participants included in the current analysis – this is the denominator for the percentages shown. Abbreviations: CST; cervical screening test.
*40 participants who were both <27 years of age and never sexually active are coded here.
@ Individuals who have never been sexually active, do not require cervical screening in Australia
#Individuals are invited to participate in the National Program at 24 years and 9 months of age, and are considered under-screened if they are at least 2.5 years overdue for their CST
^Individuals aged <30 would be appropriately screening if they have had 1 CST.

**Supplementary Figure 2. How participants first heard of self-collection (N = 6,844).**

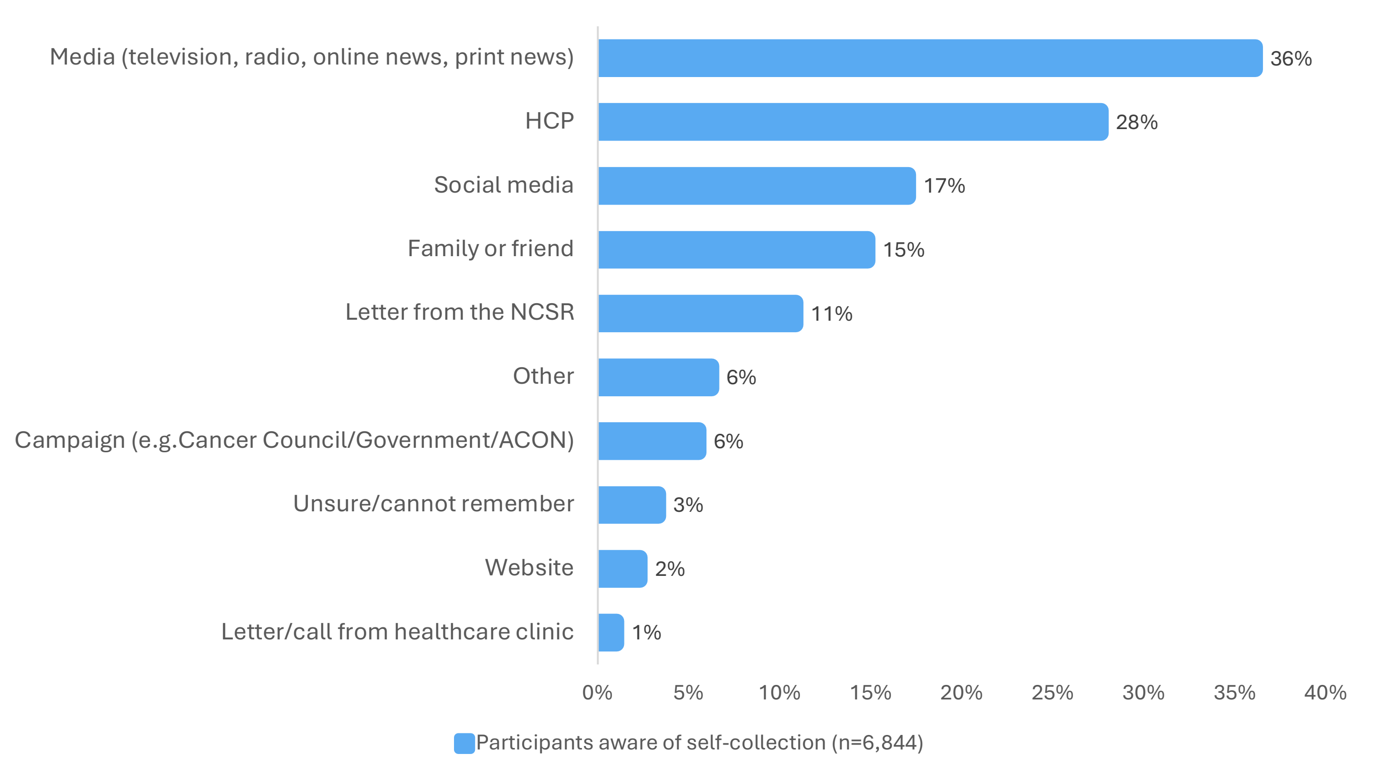

**Notes:** N = 6,844. % does not equal 100, and participants could select more than 1 option.
**Abbreviations:** HCP: Healthcare provider, NCSR: National cancer screening register.

#### Supplementary Figure 3. Prompt for last CST among participants screened since July-2022 (N= 4,529)

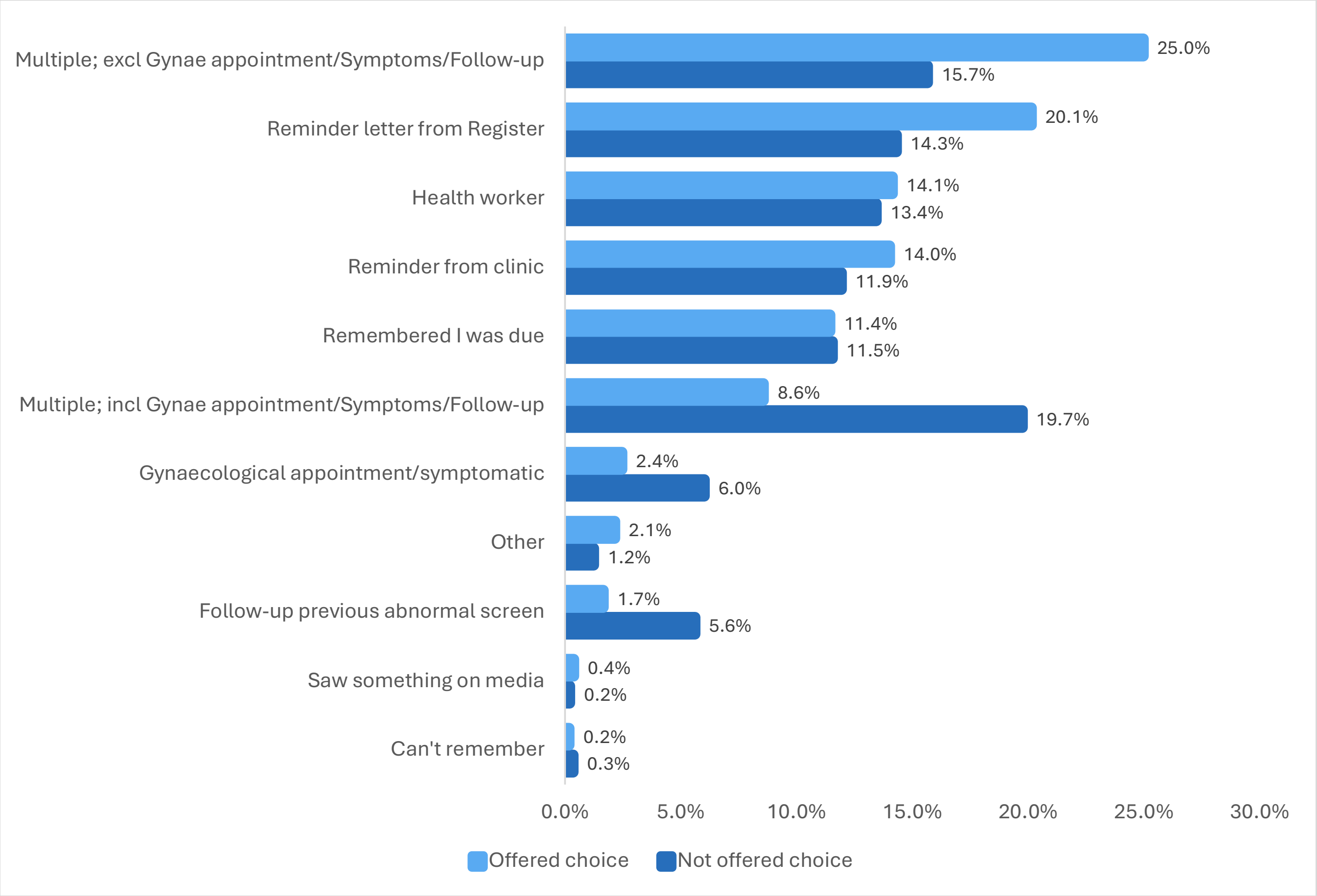

**Notes:** N= 4,529. Participants could select more than 1 option – those who selected more than 1 option are included in the multiple, including/excluding gynaecological appointment, symptoms and follow-up.

**Supplementary Figure 4: Reasons for using clinician-collection or self-collection**

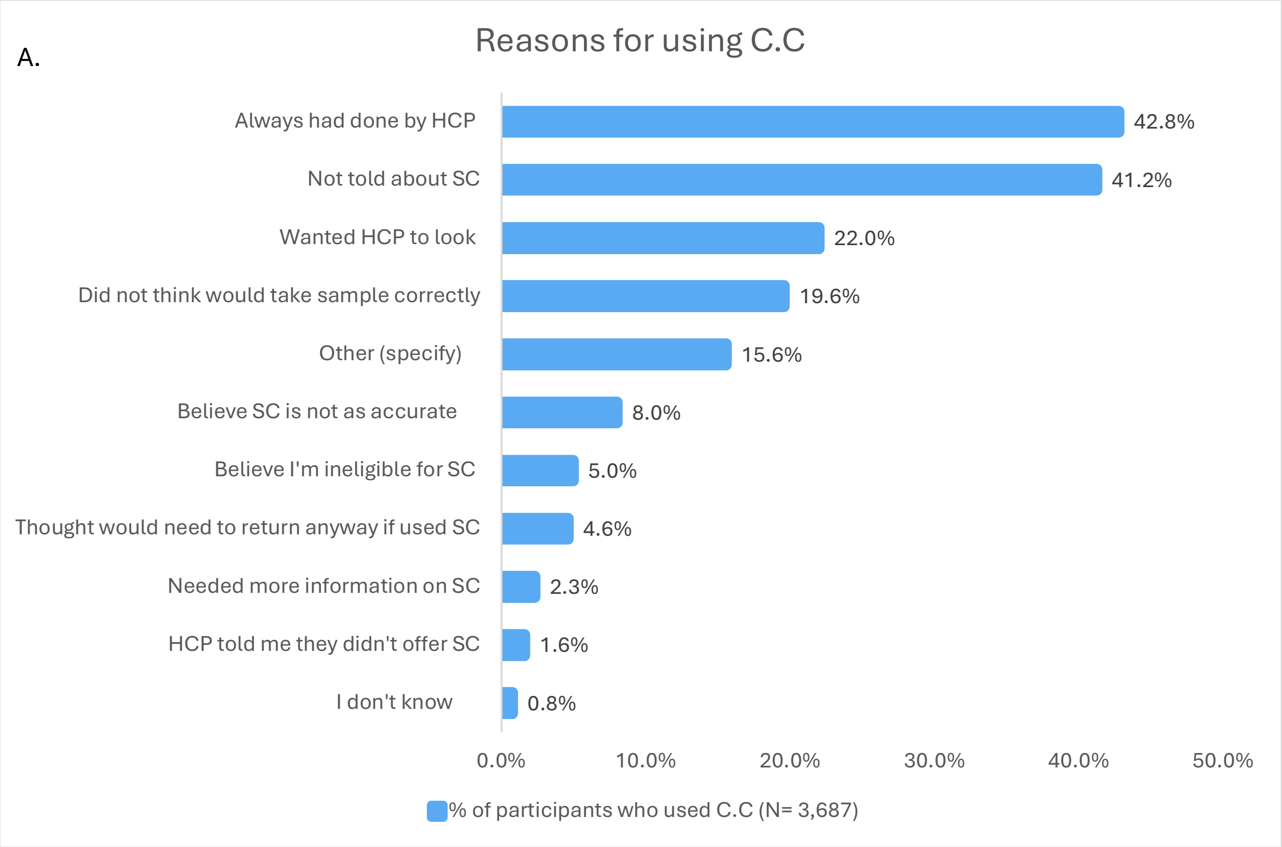

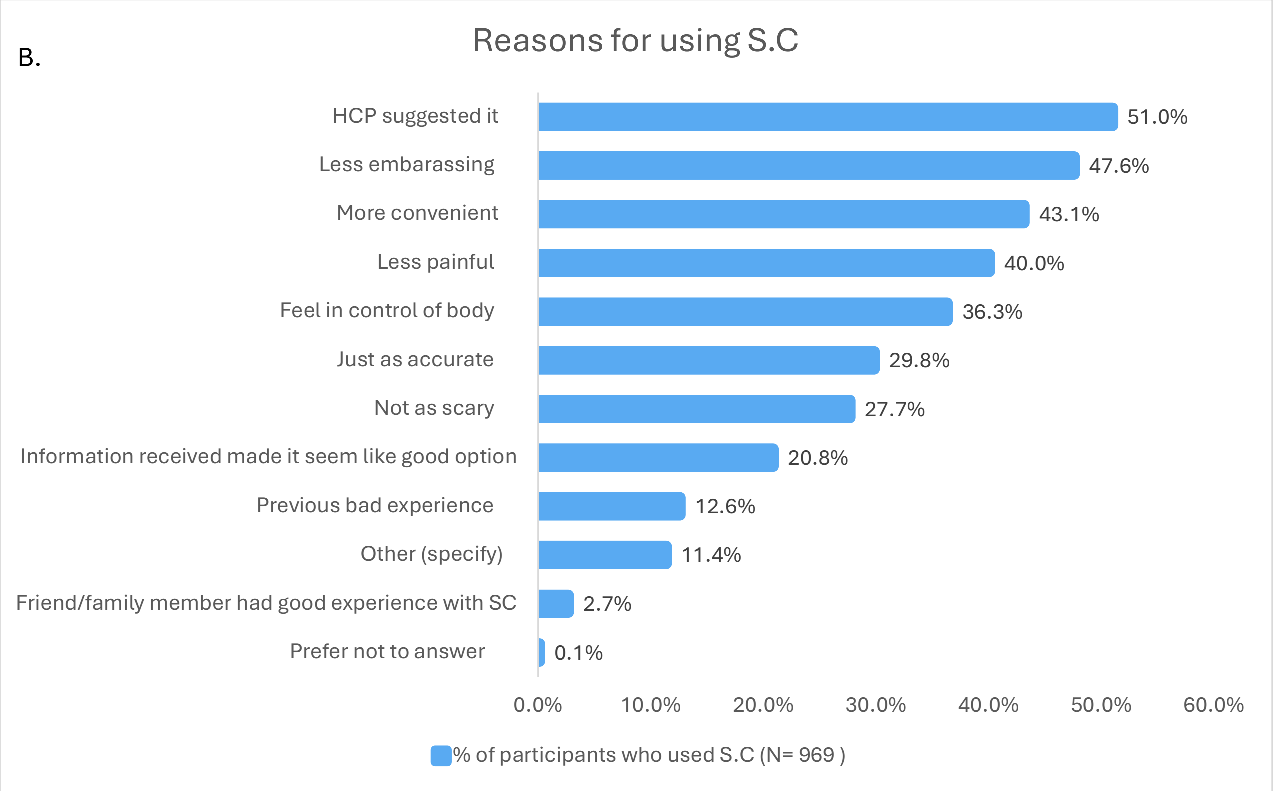

**Notes:** % Does not equal 100, as participants could select more than 1 option.
**Abbreviations:** C.C; Clinician-collected, S.C; Self-collected, HCP: Healthcare provider.

**Supplementary Figure 5. Recall among self-collection users that key information was received during their most recent cervical screening test (N=964 )**

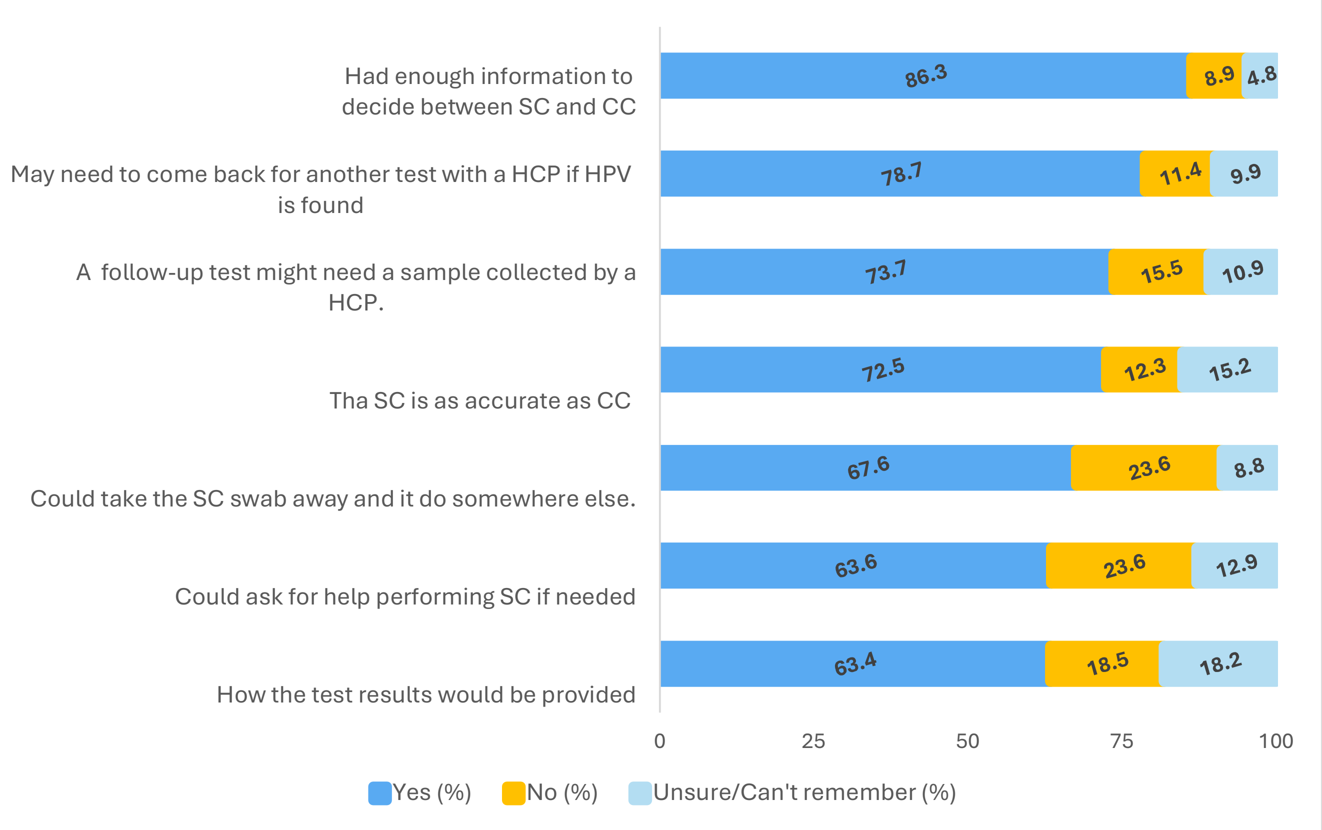

**Abbreviations:** C.C; Clinician-collected, S.C; Self-collected, HCP: Healthcare provider, HPV; Human Papillomavirus.

**Supplementary Figure 6. Statements of agreement among self-collection users regarding their experience (N=950-964)**

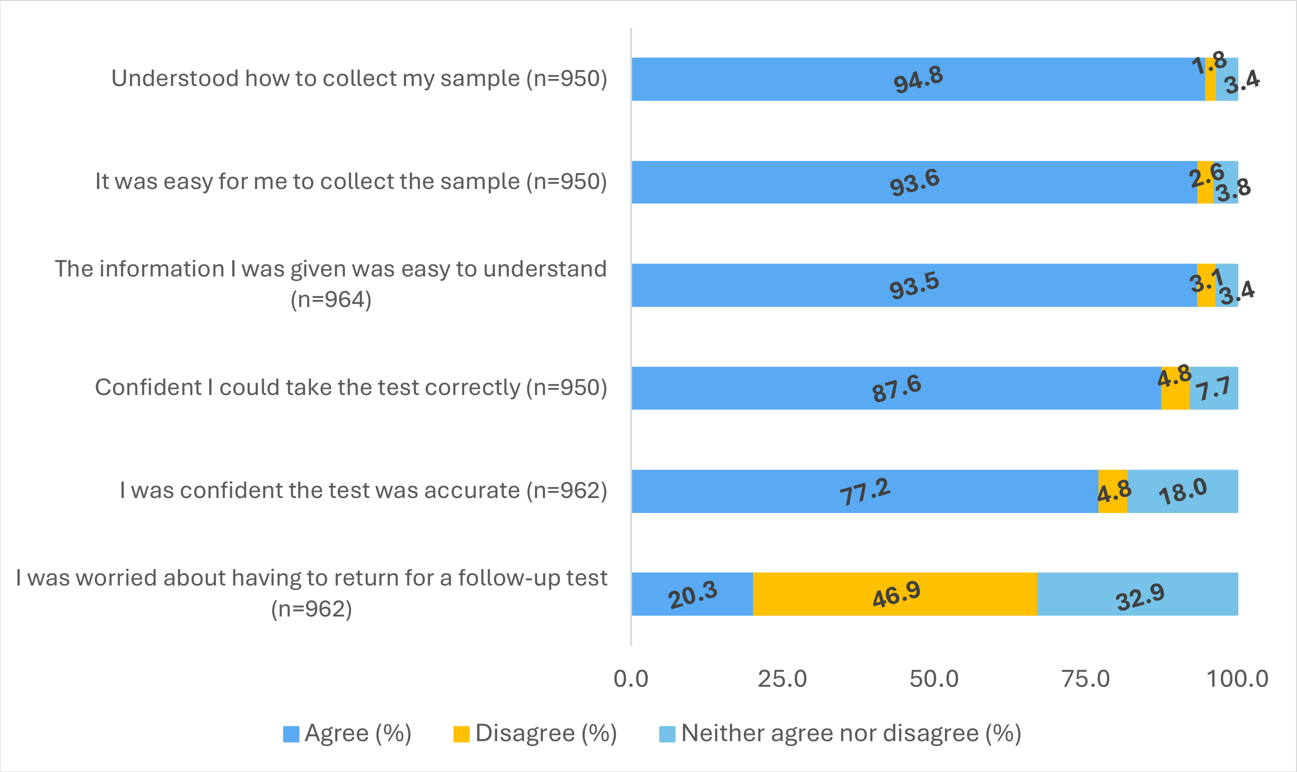

Abbreviations: C.C; Clinician-collected, S.C; Self-collected, HCP: Healthcare provider, HPV; Human Papillomavirus.

### Supplementary Tables

#### Supplementary Table 1: Research questions and modelled outcome variables

| Research Question | Survey Question | Participant group | Survey Options | Modelled options for analysis |
|---|---|---|---|---|
| 1. Awareness of Self-collection | "Have you ever heard of 'self-collection', 'self-sampling', or 'self-testing' as an option for cervical screening?". | All participants | **1.** Yes<br>**2.** No<br>**3.** Unsure<br>**4.** I prefer not to answer. | **0.** No<br>**1.** Yes<br>Excluded: Unsure and I prefer not to answer. |
| 2. Offered a choice in CST method | "For your last cervical screening test, were you offered the choice between self-collection or having a sample collected by the healthcare provider using a speculum?" | Participants screened since July 2022* | **1.** Yes, I was offered a choice<br>**2.** No, I was not offered a choice<br>**3.** I cannot remember<br>**4.** I prefer not to answer | **0.** No<br>**1.** Yes<br>Excluded: I cannot remember, and I prefer not to answer. |
| 3.'Use of self-collection' compared to clinician collection | "How did you do your last cervical screening test?". | Participants Screened since July 2022* | **1.** I 'used self-collection' in the clinic<br>**2.** I 'used self-collection' at home or somewhere else<br>**3.** A healthcare provider helped me to collect my own sample (without a speculum)<br>**4.** A healthcare provider collected the sample using a speculum<br>**5.** I cannot remember<br>**6.** I prefer not to answer | **0.** A healthcare provider collected the sample using a speculum<br>**1.** Used self-collection, includes those who responded with either of 'I 'used self-collection' in the clinic', or 'I 'used self-collection' at home or somewhere else'<br>Excluded; A healthcare provider helped me to collect my own sample (without a speculum), I cannot remember, and I prefer not to answer |
| 4.'Chose self-collection' compared to clinician collection | "How did you do your last cervical screening test?". | Participants who were offered choice† | **1.** I 'used self-collection' in the clinic<br>**2.** I 'used self-collection' at home or somewhere else<br>**3.** A healthcare provider helped me to collect my own sample (without a speculum)<br>**4.** A healthcare provider collected the sample using a speculum<br>**5.** I cannot remember<br>**6.** I prefer not to answer | **0.** A healthcare provider collected the sample using a speculum<br>**1.** Chose self-collection, includes those who responded with either 'I 'used self-collection' in the clinic' or 'I 'used self-collection' at home or somewhere else'<br>Excluded; A healthcare provider helped me to collect my own sample (without a speculum), I cannot remember, and I prefer not to answer |

#### Supplementary Table 2: Demographic Characteristics and Screening History of All Survey Participants by Date of Last Cervical Screen

| | Total N (%) | After July-22 n (%) | Before July-22 n (%) | Never n (%) | Can't remember n (%) |
|---|---|---|---|---|---|
| **Total** | 9928 (100.0%) | 4738 (47.7%) | 4014 (40.4%) | 637 (6.4%) | 539 (5.4%) |
| **Mean Age [SD]** | 42.53 [11.56] | 42.78 [11.74] | 43.54 [10.94] | 31.25 [8.05] | 46.03 [10.62] |
| **Age group (years)** | | | | | |
| **20-24** | 177 (1.8) | 41 (0.9) | 18 (0.4) | 114 (17.9) | 4 (0.7) |
| **25-34** | 2631 (26.5) | 1312 (27.7) | 908 (22.6) | 340 (53.4) | 71 (13.2) |
| **35-44** | 3202 (32.3) | 1470 (31.0) | 1420 (35.4) | 137 (21.5) | 175 (32.5) |
| **45-54** | 2203 (22.2) | 1057 (22.3) | 947 (23.6) | 36 (5.7) | 163 (30.2) |
| **55-64** | 1288 (13.0) | 632 (13.3) | 545 (13.6) | 7 (1.1) | 104 (19.3) |
| **Over 65** | 427 (4.3) | 226 (4.8) | 176 (4.4) | <5 (0.5) | 22 (4.1) |
| **Gender [b,c]** | | | | | |
| **Cis** | 9801 (98.7) | 4684 (98.9) | 3972 (99.0) | 609 (95.6) | 536 (99.4) |
| **Trans** | 118 (1.2) | 50 (1.1) | 39 (1.0) | 26 (4.1) | <5 (0.6) |
| **Not stated** | 9 (0.1) | <5 (0.1) | <5 (0.1) | <5 (0.3) | 0 (0.0) |
| **Intersex [b]** | | | | | |
| **No** | 9823 (98.9) | 4692 (99.0) | 3981 (99.2) | 623 (97.8) | 527 (97.8) |
| **Yes** | 19 (0.2) | 11 (0.2) | 8 (0.2) | 0 (0.0) | 0 (0.0) |
| **PNA/Unsure** | 74 (0.7) | 29 (0.6) | 21 (0.5) | 13 (2.0) | 11 (2.0) |
| **Missing Data** | 12 (0.1) | 6 (0.1) | <5 (0.1) | <5 (0.2) | <5 (0.2) |
| **Sexuality [b]** | | | | | |
| **Heterosexual** | 8692 (87.6) | 4190 (88.4) | 3566 (88.8) | 450 (70.6) | 486 (90.2) |
| **Gay/Lesbian** | 191 (1.9) | 64 (1.4) | 88 (2.2) | 28 (4.4) | 11 (2.0) |
| **Bisexual** | 750 (7.6) | 358 (7.6) | 277 (6.9) | 88 (13.8) | 27 (5.0) |
| **I use a different term** | 190 (1.9) | 78 (1.6) | 54 (1.3) | 47 (7.4) | 11 (2.0) |
| **Not stated** | 104 (1.0) | 47 (1.0) | 29 (0.7) | 24 (3.8) | <5 (0.7) |
| **Missing Data** | <5 (0.0) | <5 (0.0) | 0 (0.0) | 0 (0.0) | 0 (0.0) |
| **State** | | | | | |
| **ACT** | 334 (3.4) | 154 (3.3) | 137 (3.4) | 25 (3.9) | 18 (3.3) |
| **NSW** | 3107 (31.3) | 1433 (30.2) | 1317 (32.8) | 203 (31.9) | 154 (28.6) |
| **NT** | 181 (1.8) | 102 (2.2) | 65 (1.6) | 5 (0.8) | 9 (1.7) |
| **QLD** | 2025 (20.4) | 950 (20.1) | 824 (20.5) | 144 (22.6) | 107 (19.9) |
| **SA** | 571 (5.8) | 274 (5.8) | 222 (5.5) | 41 (6.4) | 34 (6.3) |
| **TAS** | 344 (3.5) | 171 (3.6) | 136 (3.4) | 22 (3.5) | 15 (2.8) |
| **VIC** | 2202 (22.2) | 1082 (22.8) | 839 (20.9) | 153 (24.0) | 128 (23.7) |
| **WA** | 1164 (11.7) | 572 (12.1) | 474 (11.8) | 44 (6.9) | 74 (13.7) |
| **Remoteness of residence [d,e]** | | | | | |
| **Major city** | 6346 (63.9) | 3028 (63.9) | 2548 (63.5) | 425 (66.7) | 345 (64.0) |
| **Inner regional** | 2015 (20.3) | 988 (20.9) | 807 (20.1) | 112 (17.6) | 108 (20.0) |
| **Outer regional, remote, very remote** | 966 (9.7) | 440 (9.3) | 417 (10.4) | 51 (8.0) | 58 (10.8) |
| **Missing data** | 601 (6.1) | 282 (6.0) | 242 (6.0) | 49 (7.7) | 28 (5.2) |
| **SEIFA [d,e]** | | | | | |
| **1 (most disadvantaged)** | 1040 (10.5) | 470 (9.9) | 410 (10.2) | 92 (14.4) | 68 (12.6) |
| **2** | 1695 (17.1) | 813 (17.2) | 670 (16.7) | 100 (15.7) | 112 (20.8) |
| **3** | 2099 (21.1) | 996 (21.0) | 847 (21.1) | 131 (20.6) | 125 (23.2) |
| **4** | 2271 (22.9) | 1054 (22.2) | 956 (23.8) | 136 (21.4) | 125 (23.2) |
| **5 (most advantaged)** | 2222 (22.4) | 1123 (23.7) | 889 (22.1) | 129 (20.3) | 81 (15.0) |
| **Missing Data** | 601 (6.1) | 282 (6.0) | 242 (6.0) | 49 (7.7) | 28 (5.2) |
| **Country of birth** | | | | | |
| **Australia** | 8195 (82.5) | 3897 (82.2) | 3295 (82.1) | 550 (86.3) | 453 (84.0) |
| **Other** | 1715 (17.3) | 834 (17.6) | 712 (17.7) | 84 (13.2) | 85 (15.8) |
| **Missing Data** | 18 (0.2) | 7 (0.1) | 7 (0.2) | <5 (0.5) | <5 (0.2) |
| **Language spoken at home** | | | | | |
| **English** | 9757 (98.3) | 4668 (98.5) | 3950 (98.4) | 602 (94.5) | 537 (99.6) |
| **Other** | 160 (1.6) | 67 (1.4) | 58 (1.4) | 33 (5.2) | <5 (0.4) |

| | | | | | |
|---|---|---|---|---|---|
| **I prefer not to answer** | 5 (0.1) | <5 (0.0) | <5 (0.0) | <5 (0.3) | 0 (0.0) |
| **Missing Data** | 6 (0.1) | <5 (0.0) | 5 (0.1) | 0 (0.0) | 0 (0.0) |
| | **Total N (%)** | **After July-22 n (%)** | **Before July-22 n (%)** | **Never n (%)** | **Can't remember n (%)** |
| **Duration of time in Australia** | | | | | |
| **Australian born** | 8195 (82.5) | 3897 (82.2) | 3295 (82.1) | 550 (86.3) | 453 (84.0) |
| **0-4 years** | 121 (1.2) | 54 (1.1) | 46 (1.1) | 19 (3.0) | <5 (0.4) |
| **5-9 years** | 182 (1.8) | 98 (2.1) | 70 (1.7) | 11 (1.7) | <5 (0.6) |
| **>10 years** | 1380 (13.9) | 665 (14.0) | 582 (14.5) | 52 (8.2) | 81 (15.0) |
| **Missing Data** | 50 (0.5) | 24 (0.5) | 21 (0.5) | 5 (0.8) | 0 (0.0) |
| **Highest education attained** | | | | | |
| **Tertiary** | 6096 (61.4) | 2940 (62.1) | 2466 (61.4) | 432 (67.8) | 258 (47.9) |
| **Tafe/Certificate** | 2551 (25.7) | 1194 (25.2) | 1058 (26.4) | 133 (20.9) | 166 (30.8) |
| **School** | 1233 (12.4) | 581 (12.3) | 474 (11.8) | 70 (11.0) | 108 (20.0) |
| **Other/PNA** | 48 (0.5) | 23 (0.5) | 16 (0.4) | <5 (0.3) | 7 (1.3) |
| **Prior screening history[f]** | | | | | |
| **Regular screeners** | 6110 (61.5) | 3715 (78.4) | 2395 (59.7) | 0 (0.0) | 0 (0.0) |
| **Not-Regular screeners** | 2642 (26.6) | 1023 (21.6) | 1619 (40.3) | 0 (0.0) | 0 (0.0) |
| **Never** | 262 (2.6) | 0 (0.0) | 0 (0.0) | 262 (41.1) | 0 (0.0) |
| **Never <27yrs** | 216 (2.2) | 0 (0.0) | 0 (0.0) | 216 (33.9) | 0 (0.0) |
| **Can't remember** | 539 (5.4) | 0 (0.0) | 0 (0.0) | 0 (0.0) | 539 (100.0) |
| **Never sexually active** | 159 (1.6) | 0 (0.0) | 0 (0.0) | 159 (25.0) | 0 (0.0) |
| **Heard of self-collection** | | | | | |
| **No** | 2898 (29.2) | 1053 (22.2) | 1434 (35.7) | 200 (31.4) | 211 (39.1) |
| **Yes** | 6830 (68.8) | 3636 (76.7) | 2489 (62.0) | 406 (63.7) | 299 (55.5) |
| **PNA/Unsure** | 187 (1.9) | 44 (0.9) | 88 (2.2) | 27 (4.2) | 28 (5.2) |
| **Missing data** | 13 (0.1) | 5 (0.1) | <5 (0.1) | <5 (0.6) | <5 (0.2) |

**Notes;** Abbreviations: CI, Confidence Interval; PNA, Prefer not to answer; SEIFA, Socio-Economic Indexes for Areas; SC, self-collection, SD, Standard Deviation; OR; Odds ratio.
Column percentages shown.
^ Adjusted for; age group, state of residence, remoteness of residence, country of birth, language spoken at home, highest level of education attained, prior screening history, location of last CST.
b Question development and reporting are according to ACON indicators (Recommended Community Indicators, n.d.)
c Analysed and reported on in line with the ABS two-step method (Standard for Sex, Gender, Variations of Sex Characteristics and Sexual Orientation Variables, 2020 | Australian Bureau of Statistics, 2023), Not stated refers to participants who selected prefer not to answer to the gender question.
[d] Codes are according to the Australian Bureau of Statistics (25)
[e] Postcode data is missing, invalid or could not be mapped to ABS datafiles. The Postcode question was not mandatory when included in the survey from February 2024
[f] Prior screening history is a modelled variable- see supplementary figure 1

#### Supplementary Results: key differences in unadjusted, adjusted, sensitivity and complete case analyses

All variables included in the unadjusted regression model, including those which were not significantly associated with being offered a choice in screening method, prior awareness of self-collection, or choosing self-collection, are shown in the supplementary text (supplementary table 3A – 6A). All results from the 'used self-collection' analysis are included in the supplementary text rather than the main text.

In terms of key differences in the unadjusted and adjusted regression models, while increasing age was associated with increasing prior 'awareness of self-collection' in the adjusted model, after controlling for other demographic and screening variables in the offer of choice, using and choosing self-collection analysis, it was not significant. Similarly, while 'offer of choice' was associated with sexuality in the unadjusted model, 95% confidence intervals were wide due to relatively small numbers among non-heterosexual groups, leading to a non-significant result in the adjusted analysis.

Furthermore, remoteness of residence was non-significant across unadjusted and adjusted models for awareness of self-collection, and significant in the unadjusted, but non-significant in the adjusted model for offer of choice. For 'use of self-collection' remoteness was significant in unadjusted, adjusted and the sensitivity analysis excluding prior screening history. Whereas, for 'chose self-collection' remoteness was significant in the unadjusted, but non-significant in the adjusted and the sensitivity analysis excluding prior screening history. Despite this, remoteness of residence was included in all adjusted models, as it is regarded as a key factor in participation in cervical screening in Australia.

##### Sensitivity Analysis Results

Sensitivity analysis was undertaken on the 'chose self-collection' and 'used self-collection' outcome variables excluding the screening history variable, as screening history is potentially a mediator variable between, for example, remoteness of residence and desire to use self-collection. Removing the prior screening history variable, resulted in one variation in the 'chose self-collection' and 'used self-collection' analyses (supplementary Table 5A and 6A). In the 'chose self-collection' analysis for "other" screening locations, the adjusted odds ratio increased moderately from 4.45 (95% CI 0.98–20.10; p = 0.082) to 5.18 (95% CI 1.16–23.10; p = 0.048), and the p-value crossed the threshold of 0.05. In the 'used self-collection' analysis for "other" screening locations, the odds ratio increased to a similar degree, from an adjusted odds ratio of 10.06 (95% CI 3.65-27.75; p=<0.001) to 10.87 (95% CI: 3.95-29.95; p=<0.001), with the p-value remaining significantly lower than the threshold of 0.05 in both instances. The modest increase in the point estimate change, and wide confidence intervals indicate limited precision. This is likely a reflection of the 'other' category. Free text data from participants who selected 'other', indicate a heterogenous group, which included 10 participants stating they used SC at home but not providing further details on the health practice they attended, and no 'usual' location. We therefore interpret this to be due to the heterogenous nature of the 'other' group, rather than a true change in effect.

##### Complete Case Analyses Results

Complete case analyses were performed on all outcomes (supplementary tables 3B to 6B). Minor differences in some the point estimates, among several outcomes, were observed. In the 'awareness of self-collection' analysis, language spoken differed slightly among participants who reported 'prefer not to answer' adjusted OR 1.11 (95% CI; 0.11-10.80) in the case control compared to adjusted 0.40 (95% CI; 0.06-2.61) in the final regression model. While the point estimate suggests slightly higher odds among the 'prefer not to answer group' the wide 95% confidence interval remains imprecise, and the sample in this group is very small <5 and therefore easily influenced by the complete case analysis. This is shown in Supplementary Table 3B.

When analysing complete case data for the ‘offer of choice’ outcome, as shown in Supplementary Table 4B, the variable sexuality, was non-significant in the unadjusted model (p=0.079), and was therefore not included in the complete case adjusted model analysis. In the regression model using all data, shown in Supplementary Table 4A, sexuality was significant in the unadjusted model, (p=0.038), and non-significant in the adjusted model (p=0.050). This likely reflects selection introduced by complete-case analysis, with the reduced sample size altering the distribution of sexuality and related sociodemographic factors, resulting in attenuated estimates and reduced precision, with 95% confidence intervals for each category crossing the null value of 1.

**Supplementary Table 3A. Demographic and Screening History Factors Associated with Awareness of Self-Collection.**

| | Total N (%) | Heard of SC n (%) | Unadjusted OR (95% CI) | Adjusted^ OR (95% CI) |
|---|---|---|---|---|
| **Total** | 9,728 (100.0) | 6,830 (70.2) | | |
| **Mean Age [SD]** | 42.5 [11.5] | 42.8 [11.7] | | |
| **Age group (years)** | | | | |
| 20-24 | 172 (1.8) | 108 (1.58) | 0.75 (0.54-1.03) | 0.83 (0.57-1.21) |
| 25-34 | 2,574 (26.5) | 1784 (26.12) | Ref. | Ref. |
| 35-44 | 3,143 (32.3) | 2166 (31.71) | 0.98 (0.88-1.10) | 1.04 (0.93-1.17) |
| 45-54 | 2,167 (22.3) | 1542 (22.58) | 1.09 (0.96-1.24) | **1.16 (1.01-1.32)** |
| 55-64 | 1,255 (12.9) | 900 (13.18) | 1.12 (0.97-1.30) | **1.20 (1.03-1.41)** |
| Over 65 | 417 (4.3) | 330 (4.83) | **1.68 (1.31-2.16)** | **1.69 (1.31-2.18)** |
| ***P-value*** | | | **<0.001** | **<0.001** |
| ***P-trend*** | | | **<0.001** | **<0.001** |
| **Gender** [b,c] | | | | |
| Cis | 9,606 (98.7) | 6736 (98.62) | Ref. | |
| Trans | 114 (1.2) | 87 (1.27) | 1.37 (0.89-2.12) | |
| Not stated | 8 (0.1) | 7 (0.10) | 2.98 (0.37-24.25) | |
| ***P-value*** | | | 0.214 | |
| **Intersex** [b] | | | | |
| No | 9,625 (98.9) | 6758 (98.95) | Ref. | |
| Yes | 19 (0.2) | 11 (0.16) | 0.58 (0.23-1.45) | |
| PNA/Unsure | 72 (0.7) | 51 (0.75) | 1.03 (0.62-1.72) | |
| Missing Data | 12 (0.1) | 10 (0.15) | 2.12 (0.46-9.69) | |
| ***P-value*** | | | 0.512 | |
| **Sexuality** [b] | | | | |
| Heterosexual | 8,521 (87.6) | 5950 (87.12) | Ref. | |
| Gay/Lesbian | 187 (1.9) | 136 (1.99) | 1.15 (0.83-1.59) | |
| Bisexual | 737 (7.6) | 528 (7.73) | 1.09 (0.92-1.29) | |
| I use a different term | 182 (1.9) | 143 (2.09) | **1.58 (1.11-2.26)** | |
| Not stated | 100 (1.0) | 72 (1.05) | 1.11 (0.72-1.72) | |
| Missing Data | <5 (0.0) | <5 (0.01) | Omitted. | |
| ***P-trend*** | | | 0.091 | |
| **State** | | | | |
| ACT | 325 (3.3) | 227 (3.32) | 1.04 (0.81-1.34) | 1.02 (0.79-1.32) |
| NSW | 3,053 (31.4) | 2104 (30.81) | Ref. | Ref. |
| NT | 175 (1.8) | 118 (1.73) | 0.93 (0.67-1.29) | 0.97 (0.68-1.39) |
| QLD | 1,979 (20.3) | 1292 (18.92) | **0.85 (0.75-0.96)** | 0.89 (0.78-1.00) |
| SA | 559 (5.7) | 452 (6.62) | **1.91 (1.52-2.38)** | **1.97 (1.57-2.47)** |
| TAS | 338 (3.5) | 241 (3.53) | 1.12 (0.87-1.44) | 1.05 (0.81-1.36) |
| VIC | 2,151 (22.1) | 1660 (24.30) | **1.52 (1.34-1.73)** | **1.54 (1.35-1.75)** |
| WA | 1,148 (11.8) | 736 (10.78) | **0.81 (0.70-0.93)** | **0.86 (0.74-0.99)** |
| ***P-value*** | | | **<0.001** | **<0.001** |

| | Total N (%) | Heard of SC n (%) | Unadjusted OR (95% CI) | Adjusted^ OR (95% CI) |
|---|---|---|---|---|
| **Remoteness of residence** [d,e] | | | | |
| Major city | 6,202 (63.8) | 4334 (63.46) | Ref. | Ref. |
| Inner regional | 1,983 (20.4) | 1419 (20.8) | 1.08 (0.97-1.21) | 1.07 (0.95-1.21) |
| Outer regional, remote, very remote | 946 (9.7) | 649 (9.50) | 0.94 (0.81-1.09) | 1.02 (0.86-1.20) |
| Missing data | 597(6.1) | 428 (6.3) | 1.09 (0.91-1.31) | 1.09 (0.90-1.32) |
| ***P-value*** | | | 0.279 | 0.648 |
| **SEIFA** [d,e] | | | | |
| 1 (most disadvantaged) | 1,017 (10.5) | 726 (10.63) | Ref. | |
| 2 | 1,653 (17.0) | 1166 (17.07) | 0.96 (0.81-1.14) | |
| 3 | 2,062 (21.2) | 1446 (21.17) | 0.94 (0.80-1.11) | |
| 4 | 2,225 (22.9) | 1528 (22.37) | 0.88 (0.75-1.03) | |
| 5 (most advantaged) | 2,174 (22.3) | 1536 (22.49) | 0.96 (0.82-1.14) | |
| Missing Data | 597 (6.1) | 428 (6.27) | 1.02 (0.81-1.27) | |
| ***P-value*** | | | 0.535 | |
| **Country of birth** | | | | |
| Australia | 8,031 (82.6) | 5710 (83.60) | Ref. | Ref. |
| Other | 1,679 (17.3) | 1104 (16.16) | **0.78 (0.70-0.87)** | **0.78 (0.69-0.88)** |
| Missing Data | 18 (0.2) | 16 (0.23) | 3.25 (0.75-14.15) | 2.87 (0.65-12.74) |
| ***P-value*** | | | **<0.001** | **<0.001** |
| **Language spoken at home** | | | | |
| English | 9,562 (98.3) | 6738 (98.65) | Ref. | Ref. |
| Other | 155 (1.6) | 86 (1.26) | **0.52 (0.38-0.72)** | **0.57 (0.41-0.80)** |
| PNA | 5 (0.1) | <5 (0.04) | 0.63 (0.10-3.76) | 0.40 (0.06-2.61) |
| Missing Data | 6 (0.1) | <5 (0.04) | 0.42 (0.08-2.08) | 0.30 (0.06-1.53) |
| ***P-value*** | | | **0.001** | **0.004** |
| **Duration of time in Australia** | | | | |
| Australian born | 8,031 (82.6) | 5710 (83.60) | Ref. | |
| 0-4 years | 118 (1.2) | 60 (0.88) | **0.42 (0.29-0.61)** | |
| 5-9 years | 174 (1.8) | 96 (1.41) | **0.50 (0.37-0.68)** | |
| >10 years | 1,357 (13.9) | 928 (13.59) | 0.88 (0.78-1.00) | |
| Missing Data | 48 (0.5) | 36 (0.53) | 1.22 (0.63-2.35) | |
| ***P-value*** | | | **<0.001** | |
| **Highest education attained** | | | | |
| Tertiary | 5,991 (61.6) | 4359 (63.82) | Ref. | Ref. |
| Tafe/Certificate | 2,483 (25.5) | 1666 (24.39) | **0.76 (0.69-0.84)** | **0.76 (0.68-0.84)** |
| School | 1,209 (12.4) | 775 (11.35) | **0.67 (0.59-0.76)** | **0.66 (0.57-0.75)** |
| Other/PNA | 45 (0.5) | 30 (0.44) | 0.75 (0.40-1.40) | 0.72 (0.38-1.37) |
| ***P-value*** | | | **<0.001** | **<0.001** |
| **Prior screening history** [f] | | | | |
| Regular screeners | 6,028 (62.0) | 4376 (64.07) | Ref. | Ref. |
| Not-Regular screeners | 2,584 (26.6) | 1749 (25.61) | **0.79 (0.72-0.87)** | **0.80 (0.72-0.89)** |
| Never | 246 (2.5) | 160 (2.34) | **0.70 (0.54-0.92)** | **0.73 (0.56-0.96)** |
| Never <27yrs | 209 (2.1) | 130 (1.90) | **0.62 (0.47-0.83)** | **0.71 (0.50-0.99)** |

| | Total N (%) | Heard of SC n (%) | Unadjusted OR (95% CI) | Adjusted^ OR (95% CI) |
|---|---|---|---|---|
| Can't remember | 510 (5.2) | 299 (4.38) | **0.53 (0.44-0.64)** | **0.53 (0.44-0.64)** |
| Never sexually active | 151 (1.6) | 116 (1.70) | 1.25 (0.85-1.83) | 1.29 (0.87-1.91) |
| ***P-value*** | | | **<0.001** | **<0.001** |
| **Location of last CST** | | | | |
| Doctors' clinic | 7,500 (77.1) | 5354 (78.39) | Ref. | Ref. |
| Women's/Sexual Health | 479 (4.9) | 364 (5.33) | **1.27 (1.02-1.57)** | **1.32 (1.06-1.64)** |
| Gynaecologist/Hospital | 587 (6.0) | 372 (5.45) | **0.69 (0.58-0.83)** | **0.70 (0.58-0.83)** |
| Other& | 42 (0.4) | 34 (0.50) | 1.70 (0.79-3.69) | 1.74 (0.80-3.80) |
| Not applicable* | 1,116 (11.5) | 705 (10.32) | **0.69 (0.60-0.78)** | omitted |
| Missing data | <5 (0.0) | <5 (0.01) | 0.13 (0.01-1.29) | 0.14 (0.01-1.41) |
| ***P-value*** | | | **<0.001** | **<0.001** |

**Notes;** Abbreviations: CI, Confidence Interval; PNA, Prefer not to answer; SEIFA, Socio-Economic Indexes for Areas; SC, self-collection, OR; Odds ratio.
Column percentages shown.
^ Adjusted for; age group, state of residence, remoteness of residence, country of birth, language spoken at home, highest level of education attained, prior screening history, location of last CST.
b Question development and reporting are according to ACON indicators (Recommended Community Indicators, n.d.)
c Analysed and reported on in line with the ABS two-step method (Standard for Sex, Gender, Variations of Sex Characteristics and Sexual Orientation Variables, 2020 | Australian Bureau of Statistics, 2023), Not stated refers to participants who selected prefer not to answer to the gender question.
[d] Codes are according to the Australian Bureau of Statistics (25)
[e] *Postcode data is missing, invalid or could not be mapped to ABS datafiles. The Postcode question was not mandatory when included in the survey from February 2024*
[f] Prior screening history is a modelled variable- see supplementary figure 1
[&] Other includes 10 participants who reported using self-collection at home, but did not provide information on the health services they attended.
*Participants not previously screened
P-value indicate the statistical significance of differences observed among groups. Significant levels P < 0.05 are denoted in bold. P-trend calculated using Wald test.

**Supplementary Table 3B. Demographic and Screening History Factors Associated with 'awareness of self-collection' – complete case analysis**

| | Heard of SC – complete case n (%) | Unadjusted OR (95%) Complete case | Adjusted^ OR (95%) Complete case | Adjusted^ OR (95% CI) – final regression* |
|---|---|---|---|---|
| **Total** | 6362 | | | |
| **Mean Age [SD]** | 42.9 [11.7] | | | |
| **Age group (years)** | | | | |
| 20-24 | 95 (1.5) | **0.69 (0.49-0.96)** | 0.74 (0.50-1.09) | 0.83 (0.57-1.21) |
| 25-34 | 1633 (25.7) | Ref. | Ref. | Ref. |
| 35-44 | 2039 (32.0) | 0.96 (0.85-1.08) | 1.01 (0.90-1.14) | 1.04 (0.93-1.17) |
| 45-54 | 1434 (22.6) | 1.07 (0.94-1.22) | 1.12 (0.98-1.28) | **1.16 (1.01-1.32)** |
| 55-64 | 853 (13.4) | 1.16 (0.99-1.35) | 1.23 (1.05-1.45) | **1.20 (1.03-1.41)** |
| Over 65 | 308 (4.8) | **1.76 (1.35-2.30)** | **1.77 (1.35-2.31)** | **1.69 (1.31-2.18)** |
| ***P-value*** | | **<0.001** | **<0.001** | **<0.001** |
| ***P-trend*** | | **<0.001** | **<0.001** | **<0.001** |
| **Gender** [b,c] | | | | |
| Cis | 6273 (98.6) | Ref. | | |
| Trans | 82 (1.3) | 1.40 (0.90-2.20) | | |
| Not stated | 7 (0.1) | 3.00 (0.37-24.37) | | |
| ***P-value*** | | 0.198 | | |
| **Intersex** [b] | | | | |
| No | 6308 (99.2) | Ref. | | |
| Yes | 10 (0.2) | 0.71 (0.26-1.96) | | |
| PNA/Unsure | 44 (0.7) | 0.99 (0.58-1.69) | | |
| ***P-value*** | | 0.802 | | |
| **Sexuality** [b] | | | | |
| Heterosexual | 5550 (87.2) | Ref. | | |
| Gay/Lesbian | 126 (2.0) | 1.17 (0.83-1.64) | | |
| Bisexual | 489 (7.7) | 1.12 (0.94-1.33) | | |
| I use a different term | 131 (2.1) | **1.50 (1.04-2.16)** | | |
| Not stated | 66 (1.0) | 1.25 (0.77-2.01) | | |
| ***P-value*** | | 0.118 | | |
| **State** | | | | |
| ACT | 209 (3.3) | 1.04 (0.80-1.35) | 1.03 (0.79-1.34) | 1.02 (0.79-1.32) |
| NSW | 1941 (30.5) | Ref. | Ref. | Ref. |
| NT | 109 (1.7) | 0.89 (0.64-1.24) | 0.92 (0.63-1.33) | 0.97 (0.68-1.39) |
| QLD | 1209 (19.0) | **0.83 (0.74-0.94)** | **0.87 (0.77-0.99)** | 0.89 (0.78-1.00) |
| SA | 430 (6.8) | **1.93 (1.53-2.43)** | **1.98 (1.57-2.51)** | **1.97 (1.57-2.47)** |
| TAS | 227 (3.6) | 1.12 (0.87-1.44) | 1.05 (0.80-1.37) | 1.05 (0.81-1.36) |
| VIC | 1535 (24.1) | **1.50 (1.32-1.71)** | **1.51 (1.33-1.73)** | **1.54 (1.35-1.75)** |
| WA | 702 (11.0) | **0.79 (0.69-0.92)** | **0.84 (0.72-0.98)** | **0.86 (0.74-0.99)** |
| ***P-value*** | | **<0.001** | **<0.001** | **<0.001** |
| **Remoteness of residence** [d,e] | | | | |

| | Heard of SC – complete case n (%) | Unadjusted OR (95%) Complete case | Adjusted^ OR (95%) Complete case | Adjusted^ OR (95% CI) – final regression* |
|---|---|---|---|---|
| Major city | 4305 (67.7) | Ref. | Ref. | Ref. |
| Inner regional | 1413 (22.2) | 1.08 (0.97-1.21) | 1.06 (0.94-1.20) | 1.07 (0.95-1.21) |
| Outer regional, remote, very remote | 644 (10.1) | 0.94 (0.81-1.09) | 1.02 (0.86-1.21) | 1.02 (0.86-1.20) |
| Missing data | | | | 1.09 (0.90-1.32) |
| ***P-value*** | | 0.200 | 0.6073 | 0.6475 |
| **SEIFA [d,e]** | | | | |
| 1 (most disadvantaged) | 722 (11.3) | Ref. | | |
| 2 | 1161 (18.3) | 0.97 (0.82-1.15) | | |
| 3 | 1437 (22.6) | 0.95 (0.80-1.12) | | |
| 4 | 1516 (23.8) | 0.88 (0.75-1.04) | | |
| 5 (most advantaged) | 1526 (24.0) | 0.97 (0.82-1.14) | | |
| ***P-value*** | | 0.482 | | |
| **Country of birth** | (0.0) | | | |
| Australia | 5341 (84.0) | Ref. | Ref. | Ref. |
| Other | 1021 (16.0) | **0.80 (0.71-0.90)** | **0.79 (0.70-0.90)** | **0.78 (0.69-0.88)** |
| Missing data | | | | 2.87 (0.65-12.74) |
| ***P-value*** | | **<0.001** | **<0.001** | **<0.001** |
| **Language spoken at home** | | | | |
| English | 6282 (98.7) | Ref. | Ref. | Ref. |
| Other | 77 (1.2) | **0.55 (0.39-0.78)** | **0.61 (0.43-0.88)** | **0.57 (0.41-0.80)** |
| PNA | <5 (0.1) | 1.27 (0.13-12.18) | 1.11 (0.11-10.80) | 0.40 (0.06-2.61) |
| Missing data | | | | 0.30 (0.06-1.53) |
| ***P-value*** | | **0.0029** | **0.0281** | **0.0034** |
| **Duration of time in Australia** | | | | |
| Australian born | 5341 (84.0) | Ref. | | |
| 0-4 years | 58 (0.9) | **0.44 (0.30-0.64)** | | |
| 5-9 years | 90 (1.4) | **0.53 (0.39-0.73)** | | |
| >10 years | 873 (13.7) | 0.89 (0.78-1.01) | | |
| Missing data | | | | |
| ***P-value*** | | **<0.001** | | |
| **Highest education attained** | | | | |
| Tertiary | 4041 (63.5) | Ref. | Ref. | Ref. |
| Tafe/Certificate | 1576 (24.8) | **0.76 (0.68-0.84)** | **0.75 (0.68-0.84)** | **0.76 (0.68-0.84)** |
| School | 720 (11.3) | **0.67 (0.59-0.77)** | **0.66 (0.57-0.75)** | **0.66 (0.57-0.75)** |
| Other/PNA | 25 (0.4) | 0.67 (0.35-1.29) | 0.66 (0.33-1.29) | 0.72 (0.38-1.37) |
| ***P-value*** | | **<0.001** | **<0.001** | **<0.001** |
| **Prior screening history[f]** | | | | |
| Regular screeners | 4074 (64.0) | Ref. | Ref. | Ref. |
| Not-Regular screeners | 1638 (25.8) | **0.80 (0.72-0.88)** | **0.81 (0.73-0.90)** | **0.80 (0.72-0.89)** |
| Never | 146 (2.3) | **0.68 (0.52-0.90)** | **0.72 (0.54-0.95)** | **0.73 (0.56-0.96)** |
| Never <27yrs | 116 (1.8) | **0.63 (0.47-0.85)** | 0.76 (0.53-1.09) | **0.71 (0.50-0.99)** |
| Can't remember | 281 (4.4) | **0.54 (0.44-0.65)** | **0.53 (0.44-0.65)** | **0.53 (0.44-0.64)** |

| | Heard of SC – complete case n (%) | Unadjusted OR (95%) Complete case | Adjusted^ OR (95%) Complete case | Adjusted^ OR (95% CI) – final regression* |
|---|---|---|---|---|
| Never sexually active | 107 (1.7) | 1.16 (0.79-1.71) | 1.18 (0.80-1.76) | 1.29 (0.87-1.91) |
| ***P-value*** | | **<0.001** | **<0.001** | **<0.001** |
| **Location of last CST** | | | | |
| Doctors' clinic | 4996 (78.5) | Ref. | Ref. | Ref. |
| Women's/Sexual Health | 342 (5.4) | **1.33 (1.06-1.66)** | **1.37 (1.09-1.73)** | **1.32 (1.06-1.64)** |
| Gynaecologist/Hospital | 342 (5.4) | **0.68 (0.57-0.82)** | **0.68 (0.57-0.82)** | **0.70 (0.58-0.83)** |
| **Other&** | 32 (0.5) | 1.84 (0.81-4.18) | 1.93 (0.84-4.41) | 1.74 (0.80-3.80) |
| **Not applicable*** | 650 (10.2) | **0.68 (0.59-0.78)** | omitted | omitted |
| **Missing data** | | | | 0.14 (0.01-1.41) |
| ***P-value*** | | **<0.001** | **<0.001** | **<0.001** |

**Notes;** Abbreviations: CI, Confidence Interval; PNA, Prefer not to answer; SEIFA, Socio-Economic Indexes for Areas; SC, self-collection, OR; Odds ratio.
Column percentages shown.
^ Adjusted for; age group, state of residence, remoteness of residence, country of birth, language spoken at home, highest level of education attained, prior screening history, location of last CST.
b Question development and reporting are according to ACON indicators (Recommended Community Indicators, n.d.)
c Analysed and reported on in line with the ABS two-step method (Standard for Sex, Gender, Variations of Sex Characteristics and Sexual Orientation Variables, 2020 | Australian Bureau of Statistics, 2023), Not stated refers to participants who selected prefer not to answer to the gender question.
[d] Codes are according to the Australian Bureau of Statistics (25)
[e] Postcode data is missing, invalid or could not be mapped to ABS datafiles. The Postcode question was not mandatory when included in the survey from February 2024
[f] Prior screening history is a modelled variable- see supplementary figure 1
[&] Other includes 10 participants who reported using self-collection at home, but did not provide information on the health services they attended.
*Participants not previously screened
P-value indicate the statistical significance of differences observed among groups. Significant levels P < 0.05 are denoted in bold. P-trend calculated using Wald test.

**Supplementary Table 4A. Demographic and Screening History Factors Associated with 'offer of choice' in screening method.**

| | Total N (%) | Offered Choice n (%) | Unadjusted OR (95% CI) | Adjusted^ OR (95% CI) |
|---|---|---|---|---|
| **Total** | 4,529 (100) | 1,635 (36.1) | | |
| **Mean Age [SD]** | 42.8 [11.7] | 43.6 [12.2] | | |
| **Age group (years)** | | | | |
| 20-24 | 41 (0.9) | 10 (0.6) | 0.59 (0.29-1.22) | 0.87 (0.40-1.88) |
| 25-34 | 1,243 (27.4) | 439 (26.9) | Ref. | Ref. |
| 35-44 | 1,410 (31.1) | 468 (28.6) | 0.91 (0.77-1.07) | **0.82 (0.69-0.97)** |
| 45-54 | 1,014 (22.4) | 384 (23.5) | 1.12 (0.94-1.33) | 0.92 (0.76-1.11) |
| 55-64 | 602 (13.3) | 232 (14.2) | 1.15 (0.94-1.40) | 0.90 (0.72-1.12) |
| Over 65 | 219 (4.8) | 102 (6.2) | **1.60 (1.19-2.13)** | 1.24 (0.91-1.70) |
| ***P-value*** | | | **<0.001** | 0.068 |
| ***P-trend*** | | | **<0.001** | 0.555 |
| **Gender** [b,c] | | | | |
| Cis | 4,477 (98.9) | 1,618 (99.0) | Ref. | |
| Trans | 48 (1.1) | 16 (1.0) | 0.88 (0.48-1.62) | |
| Not stated | <5 (0.1) | <5 (0.1) | 0.59 (0.06-5.67) | |
| ***P-value*** | | | 0.831 | |
| **Intersex** [b] | | | | |
| No | 4,488 (99.1) | 1,618 (99.0) | Ref. | |
| Yes | 10 (0.2) | 5 (0.3) | 1.77 (0.51-6.14) | |
| PNA/Unsure | 25 (0.6) | 10 (0.6) | 1.18 (0.53-2.64) | |
| Missing Data | 6 (0.1) | <5 (0.1) | 0.89 (0.16-4.85) | |
| ***P-value*** | | | 0.800 | |
| **Sexuality** [b] | | | | |
| Heterosexual | 4,013 (88.6) | 1,448 (88.6) | Ref. | Ref. |
| Gay/Lesbian | 60 (1.3) | 28 (1.7) | 1.55 (0.93-2.58) | **1.72 (1.00-2.98)** |
| Bisexual | 339 (7.5) | 114 (7.0) | 0.90 (0.71-1.13) | 0.96 (0.75-1.24) |
| I use a different term | 73 (1.6) | 34 (2.1) | 1.54 (0.97-2.46) | 1.63 (0.99-2.69) |
| Not stated | 43 (0.9) | 10 (0.6) | 0.54 (0.26-1.09) | 0.60 (0.29-1.27) |
| Missing Data | <5 (0.0) | <5 (0.1) | omitted. | omitted. |
| ***P-value*** | | | 0.038 | 0.050 |
| **State** | | | | |
| ACT | 145 (3.2) | 50 (3.1) | 1.05 (0.74-1.51) | 1.06 (0.73-1.55) |
| NSW | 1,364 (30.1) | 454 (27.8) | Ref. | Ref. |
| NT | 100 (2.2) | 42 (2.6) | 1.45 (0.96-2.19) | **1.71 (1.05-2.77)** |
| QLD | 909 (20.1) | 297 (18.2) | 0.97 (0.81-1.16) | 1.05 (0.87-1.27) |
| SA | 258 (5.7) | 92 (5.6) | 1.11 (0.84-1.47) | 1.18 (0.88-1.59) |
| TAS | 164 (3.6) | 79 (4.8) | **1.86 (1.34-2.58)** | **1.51 (1.06-2.15)** |

| | Total N (%) | Offered Choice n (%) | Unadjusted OR (95% CI) | Adjusted^ OR (95% CI) |
|---|---|---|---|---|
| VIC | 1,038 (22.9) | 433 (26.5) | **1.43 (1.21-1.70)** | **1.49 (1.25-1.78)** |
| WA | 551 (12.2) | 188 (11.5) | 1.04 (0.84-1.28) | 1.10 (0.88-1.37) |
| ***P-value*** | | | **<0.001** | **<0.001** |
| **Remoteness of residence** [d,e] | | | | |
| Major city | 2,879 (63.6) | 991 (60.6) | Ref. | Ref. |
| Inner regional | 953 (21.0) | 387 (23.7) | **1.30 (1.12-1.51)** | **1.23 (1.04-1.46)** |
| Outer regional, remote and very remote | 429 (9.5) | 160 (9.8) | 1.13 (0.92-1.40) | 1.03 (0.80-1.32) |
| Missing data | 268 (5.9) | 97 (5.9) | 1.08 (0.83-1.40) | 1.09 (0.82-1.44) |
| ***P-value*** | | | **0.007** | 0.118 |
| **SEIFA** [d,e] | | | | |
| 1 (most disadvantaged) | 446 (9.8) | 164 (10.0) | Ref. | |
| 2 | 778 (17.2) | 278 (17.0) | 0.96 (0.75-1.22) | |
| 3 | 959 (21.2) | 346 (21.2) | 0.97 (0.77-1.23) | |
| 4 | 1,015 (22.4) | 377 (23.1) | 1.02 (0.81-1.28) | |
| 5 (most advantaged) | 1,063 (23.5) | 373 (22.8) | 0.93 (0.74-1.17) | |
| Missing Data | 268 (5.9) | 97 (5.9) | 0.98 (0.71-1.34) | |
| ***P-value*** | | | 0.956 | |
| **Country of birth** | | | | |
| Australia | 3,731 (82.4) | 1,362 (83.3) | Ref. | |
| Other | 792 (17.5) | 269 (16.5) | 0.89 (0.76-1.05) | |
| Missing Data | 6 (0.1) | <5 (0.2) | 3.48 (0.64-19.02) | |
| ***P-value*** | | | 0.138 | |
| **Language spoken at home** | | | | |
| English | 4,464 (98.6) | 1,613 (98.7) | Ref. | |
| Other | 62 (1.4) | 21 (1.3) | 0.91 (0.53-1.54) | |
| PNA | <5 (0.0) | <5 (0.1) | 1.77 (0.11-28.28) | |
| Missing Data | <5 (0.0) | 0 (0.0) | omitted | |
| ***P-value*** | | | 0.861 | |
| **Duration of time in Australia** | | | | |
| Australian born | 3,731 (82.4) | 1,362 (83.3) | Ref. | Ref. |
| 0-4 years | 49 (1.1) | 8 (0.5) | **0.34 (0.16-0.73)** | **0.42 (0.19-0.91)** |
| 5-9 years | 95 (2.1) | 23 (1.4) | **0.56 (0.35-0.89)** | 0.68 (0.42-1.13) |
| >10 years | 633 (14.0) | 234 (14.3) | 1.02 (0.86-1.21) | 0.95 (0.79-1.15) |
| Missing Data | 21 (0.5) | 8 (0.5) | 1.07 (0.44-2.59) | 1.02 (0.41-2.52) |
| ***P-value*** | | | **0.008** | 0.136 |
| **Highest education attained** | | | | |
| Tertiary | 2,823 (62.3) | 1,039 (63.5) | Ref. | |
| Tafe/Certificate | 1,135 (25.1) | 391 (23.9) | 0.90 (0.78-1.04) | |
| School | 550 (12.1) | 197 (12.0) | 0.96 (0.79-1.16) | |

| | Total N (%) | Offered Choice n (%) | Unadjusted OR (95% CI) | Adjusted^ OR (95% CI) |
|---|---|---|---|---|
| Other/PNA | 21 (0.5) | 8 (0.5) | 1.06 (0.44-2.56) | |
| ***P-value*** | | | 0.572 | |
| **Prior screening history[f]** | | | | |
| Regular screeners | 3,548 (78.3) | 1,292 (79.0) | Ref. | |
| Not-Regular screeners | 981 (21.7) | 343 (21.0) | 0.94 (0.81-1.09) | |
| ***P-value*** | | | 0.403 | |
| **Location of last cervical screen** | | | | |
| Doctors' clinic | 3,917 (86.5) | 1,463 (89.5) | Ref. | Ref. |
| Women's/Sexual health clinic | 236 (5.2) | 111 (6.8) | **1.49 (1.14-1.94)** | **1.62 (1.22-2.14)** |
| Gynaecologist/Hospital setting | 351 (7.8) | 45 (2.8) | **0.25 (0.18-0.34)** | **0.31 (0.23-0.44)** |
| Other | 24 (0.5) | 16 (1.0) | **3.35 (1.43-7.86)** | **2.72 (1.14-6.51)** |
| Missing Data | <5 (0.0) | 0 (0.0) | omitted | omitted |
| ***P-value*** | | | **<0.001** | **<0.001** |
| **Prompt for last CST** | | | | |
| Another gynaecological appointment/worried about symptom | 214 (4.7) | 40 (2.4) | **0.29 (0.20-0.42)** | **0.29 (0.20-0.43)** |
| Can't remember | 189 (4.2) | 3 (0.2) | 0.38 (0.10-1.38) | 0.36 (0.10-1.35) |
| Follow-up previous abnormal screen | 620 (13.7) | 27 (1.7) | **0.21 (0.14-0.32)** | **0.25 (0.16-0.38)** |
| Health worker told me | 583 (12.9) | 231 (14.1) | **0.75 (0.60-0.93)** | **0.76 (0.61-0.95)** |
| Other | 71 (1.6) | 35 (2.1) | 1.22 (0.75-1.99) | 1.24 (0.75-2.05) |
| Remembered I was due | 12 (0.3) | 187 (11.4) | **0.70 (0.56-0.89)** | **0.73 (0.58-0.92)** |
| Reminder from clinic | 521 (11.5) | 229 (14.0) | 0.84 (0.67-1.04) | 0.83 (0.66-1.04) |
| Reminder letter from Register | 574 (12.7) | 329 (20.1) | Ref. | Ref. |
| Saw something on media | 743 (16.4) | 6 (0.4) | 1.26 (0.40-3.94) | 1.34 (0.41-4.36) |
| Multiple- excluding gynaecological appointment/symptoms/follow-up | 989 (21.8) | 408 (25.0) | 1.13 (0.93-1.38) | 1.13 (0.92-1.38) |
| Multiple-including gynaecological appointment/symptoms/follow-up/ | 13 (0.3) | 140 (8.6) | **0.31 (0.24-0.39)** | **0.34 (0.26-0.43)** |
| ***P-value*** | | | **<0.001** | **<0.001** |

**Notes;** Abbreviations: CI, Confidence Interval; PNA, prefer not to answer; SEIFA, Socio-Economic Indexes for Areas; SC, self-collection, SD, Standard Deviation; SEIFA, Socio-Economic Indexes for Areas.
Total = Column percentages shown, offered choice = row percentages shown.
^Adjusted for; Age group, sexuality, state, remoteness of residence, duration of time in Australia, modelled prior screening history, location of last CST, and prompt for last CST.
[b] Question development and reporting are according to ACON indicators (Recommended Community Indicators, n.d.)
[c] Analysed and reported on in line with the ABS two-step method (Standard for Sex, Gender, Variations of Sex Characteristics and Sexual Orientation Variables, 2020 | Australian Bureau of Statistics, 2023), Not stated refers to participants who selected prefer not to answer to the gender question.
[d] Codes are according to the Australian Bureau of Statistics (Australian Statistical Geography Standard (ASGS) Edition 3, July 2021 - June 2026 | Australian Bureau of Statistics, 2024)
[e] *Postcode data is missing, invalid or could not be mapped to ABS datafiles. The Postcode question was not mandatory when included in the survey from February 2024*
**[f]** Prior screening history is a modelled variable- see supplementary figure 1
P-value indicate the statistical significance of differences observed among groups. Significant levels P < 0.05 are denoted in bold. P-trend calculated using Cochran–Armitage test.

#### Supplementary Table 4B. Demographic and Screening History Factors Associated with 'offer of choice' in screening method – complete case analysis

| | Offered Choice n (%) Complete case | Unadjusted OR (95%) Complete case | Adjusted^ OR (95%) Complete case | Adjusted^ OR (95% CI) – final regression* |
|---|---|---|---|---|
| **Total** | 1529 (36.1%) | | | |
| **Mean Age [SD]** | 42.9 [11.7] | | | |
| **Age group (years)** | | | | |
| 20-24 | 8 (0.5) | 0.52 (0.24-1.16) | 0.80 (0.35-1.85) | 0.87 (0.40-1.88) |
| 25-34 | 405 (26.5) | Ref. | Ref. | Ref. |
| 35-44 | 447 (29.2) | 0.91 (0.78-1.09) | **0.82 (0.69-0.98)** | **0.82 (0.69-0.97)** |
| 45-54 | 351 (23) | 1.08 (0.91-1.30) | 0.88 (0.72-1.07) | 0.92 (0.76-1.11) |
| 55-64 | 221 (14.5) | 1.17 (0.95-1.44) | 0.89 (0.71-1.12) | 0.90 (0.72-1.12) |
| Over 65 | 97 (6.3) | **1.65 (1.22-2.22)** | 1.24 (0.90-1.72) | 1.24 (0.91-1.70) |
| ***P-value*** | | **0.001** | 0.069 | 0.068 |
| ***P-trend*** | | **<0.001** | 0.676 | 0.550 |
| **Gender** [b,c] | | | | |
| Cis | 1513 (99.0) | Ref. | | |
| Trans | 15 (1.0) | 0.91 (0.49-1.71) | | |
| Not stated | <5 (0.1) | 0.59 (0.06-5.67) | | |
| ***P-value*** | | 0.865 | | |
| **Intersex** [b] | | | | |
| No | 1515 (99.1) | Ref. | | |
| Yes | 5 (0.3) | 2.22 (0.60-8.28) | | |
| PNA/Unsure | 9 (0.6) | 1.23 (0.52-2.88) | | |
| ***P-value*** | | 0.444 | | |
| **Sexuality** [b] | | | | |
| Heterosexual | 1357 (88.7) | Ref. | | Ref. |
| Gay/Lesbian | 27 (1.8) | 1.59 (0.94-2.69) | | **1.72 (1.00-2.98)** |
| Bisexual | 105 (6.9) | 0.88 (0.69-1.12) | | 0.96 (0.75-1.24) |
| I use a different term | 30 (2.0) | 1.43 (0.88-2.33) | | 1.63 (0.99-2.69) |
| Not stated | 10 (0.7) | 0.59 (0.29-1.21) | | 0.60 (0.29-1.27) |
| Missing data | | | | omitted. |
| ***P-value*** | | 0.079 | | 0.050 |
| **State** | | | | |
| ACT | 46 (3.0) | 1.00 (0.69-1.46) | 1.03 (0.69-1.52) | 1.06 (0.73-1.55) |
| NSW | 427 (27.9) | Ref. | Ref. | Ref. |
| NT | 40 (2.6) | 1.53 (0.99-2.35) | **1.77 (1.06-2.96)** | **1.71 (1.05-2.77)** |
| QLD | 279 (18.3) | 0.94 (0.78-1.13) | 1.02 (0.84-1.24) | 1.05 (0.87-1.27) |
| SA | 89 (5.8) | 1.10 (0.83-1.46) | 1.19 (0.88-1.61) | 1.18 (0.88-1.59) |
| TAS | 73 (4.8) | **1.80 (1.28-2.53)** | 1.42 (0.98-2.06) | **1.51 (1.06-2.15)** |
| VIC | 400 (26.1) | **1.38 (1.16-1.64)** | **1.45 (1.21-1.74)** | **1.49 (1.25-1.78)** |
| WA | 175 (11.5) | 0.96 (0.78-1.19) | 1.01 (0.81-1.27) | 1.10 (0.88-1.37) |
| ***P-value*** | | **<0.001** | **<0.001** | **<0.001** |

| | Offered Choice n (%) Complete case | Unadjusted OR (95%) Complete case | Adjusted^ OR (95%) Complete case | Adjusted^ OR (95% CI) – final regression* |
|---|---|---|---|---|
| **Remoteness of residence [d,e]** | | | | |
| Major city | 985 (64.5) | Ref. | Ref. | Ref. |
| Inner regional | 385 (25.1) | **1.30 (1.12-1.51)** | **1.22 (1.03-1.45)** | **1.23 (1.04-1.46)** |
| Outer regional, remote and very remote | 159 (10.4) | 1.13 (0.92-1.40) | 1.01 (0.79-1.31) | 1.03 (0.80-1.32) |
| Missing data | | | | 1.09 (0.82-1.44) |
| ***P-value*** | | **0.003** | 0.065 | 0.118 |
| **SEIFA [d,e]** | | | | |
| 1 (most disadvantaged) | 163 (10.7) | Ref. | | |
| 2 | 277 (18.1) | 0.96 (0.75-1.22) | | |
| 3 | 344 (22.5) | 0.97 (0.77-1.23) | | |
| 4 | 373 (24.3) | 1.01 (0.80-1.28) | | |
| 5 (most advantaged) | 372 (24.3) | 0.93 (0.74-1.18) | | |
| Missing data | | | | |
| ***P-value*** | | 0.936 | | |
| **Country of birth** | | | | |
| Australia | 1277 (83.5) | Ref. | | |
| Other | 252 (16.5) | 0.92 (0.78-1.09) | | |
| Missing data | | | | |
| ***P-value*** | | 0.352 | | |
| **Language spoken at home** | | | | |
| English | 1509 (98.7) | Ref. | | |
| Language Other Than English | 19 (1.2) | 0.96 (0.55-1.68) | | |
| PNA | <5 (0.1) | omitted | | |
| Missing data | | | | |
| ***P-value*** | | 0.887 | | |
| **Duration of time in Australia** | | | | |
| Australian born | 1277 (83.5) | Ref. | Ref. | Ref. |
| 0-4 years | 8 (0.5) | **0.37 (0.17-0.79)** | **0.45 (0.20-0.99)** | **0.42 (0.19-0.91)** |
| 5-9 years | 23 (1.5) | **0.60 (0.37-0.97)** | 0.74 (0.44-1.22) | 0.68 (0.42-1.13) |
| >10 years | 221 (14.5) | 1.04 (0.87-1.24) | 0.97 (0.80-1.18) | 0.95 (0.79-1.15) |
| Missing data | | | | 1.02 (0.41-2.52) |
| ***P-value*** | | **0.011** | 0.157 | 0.136 |
| **Highest education attained** | | | | |
| Tertiary | 970 (63.4) | Ref. | | |
| Tafe/Certificate | 371 (24.3) | 0.90 (0.78-1.04) | | |
| School | 182 (11.9) | 0.97 (0.80-1.18) | | |
| Other/PNA | 6 (0.4) | 0.94 (0.34-2.54) | | |
| ***P-value*** | | 0.588 | | |
| **Prior screening history** | | | | |
| Regular screeners | 1210 (79.1) | Ref. | | |
| Not-Regular screeners | 319 (20.9) | 0.91 (0.78-1.07) | | |
| ***P-value*** | | 0.254 | | |

| | Offered Choice n (%) Complete case | Unadjusted OR (95%) Complete case | Adjusted^ OR (95%) Complete case | Adjusted^ OR (95% CI) – final regression* |
|---|---|---|---|---|
| **Location of last cervical screen** | | | | |
| Doctors' clinic | 1365 (89.3) | Ref. | Ref. | Ref. |
| Women's/Sexual health clinic | 107 (7.0) | **1.53 (1.17-2.00)** | **1.69 (1.27-2.25)** | **1.62 (1.22-2.14)** |
| Gynaecologist/Hospital setting | 41 (2.7) | **0.25 (0.18-0.34)** | **0.32 (0.22-0.44)** | **0.31 (0.23-0.44)** |
| Other | 16 (1.0) | **4.49 (1.75-11.51)** | **3.87 (1.49-10.10)** | **2.72 (1.14-6.51)** |
| Missing data | | | | omitted |
| ***P-value*** | | **<0.001** | **<0.001** | **<0.001** |
| **Prompt for last CST** | | | | |
| Another gynaecological appointment/worried about symptom | 35 (2.3) | **0.27 (0.18-0.40)** | **0.28 (0.19-0.42)** | **0.29 (0.20-0.43)** |
| Can't remember | <5 (0.2) | 0.41 (0.11-1.54) | 0.39 (0.10-1.48) | **0.36 (0.10-1.35)** |
| Follow-up previous abnormal screen | 24 (1.6) | **0.20 (0.12-0.31)** | **0.23 (0.14-0.37)** | **0.25 (0.16-0.38)** |
| Health worker told me | 217 (14.1) | **0.73 (0.59-0.92)** | **0.76 (0.60-0.95)** | **0.76 (0.61-0.95)** |
| Other | 33 (2.2) | 1.17 (0.71-1.92) | 1.17 (0.70-1.96) | **1.24 (0.75-2.05)** |
| Remembered I was due | 177 (11.6) | **0.70 (0.56-0.89)** | **0.73 (0.58-0.93)** | **0.73 (0.58-0.92)** |
| Reminder from clinic | 214 (14) | 0.82 (0.65-1.03) | 0.83 (0.66-1.05) | 0.83 (0.66-1.04) |
| Reminder letter from Register | 313 (20.5) | Ref. | Ref. | Ref. |
| Saw something on media | 5 (0.3) | 1.24 (0.35-4.31) | 1.50 (0.42-5.37) | 1.34 (0.41-4.36) |
| Multiple- excluding gynaecological appointment/symptoms/follow-up | 381 (24.9) | 1.11 (0.91-1.36) | 1.12 (0.91-1.37) | 1.13 (0.92-1.38) |
| Multiple-including gynaecological appointment/symptoms/follow-up | 127 (8.3) | **0.29 (0.23-0.37)** | **0.32 (0.25-0.41)** | **0.34 (0.26-0.43)** |
| ***P-value*** | | **<0.001** | **<0.001** | **<0.001** |

**Notes;** Abbreviations: CI, Confidence Interval; PNA, prefer not to answer; SEIFA, Socio-Economic Indexes for Areas; SC, self-collection, SD, Standard Deviation; SEIFA, Socio-Economic Indexes for Areas.
Total = Column percentages shown, offered choice = row percentages shown.
*Adjusted OR and 95% CI from regression analysis included from corresponding table a for easy comparison.
^Adjusted for; Age group, sexuality, state, remoteness of residence, duration of time in Australia, modelled prior screening history, location of last CST, and prompt for last CST.
[b] Question development and reporting are according to ACON indicators (Recommended Community Indicators, n.d.)
[c] Analysed and reported on in line with the ABS two-step method (Standard for Sex, Gender, Variations of Sex Characteristics and Sexual Orientation Variables, 2020 | Australian Bureau of Statistics, 2023), Not stated refers to participants who selected prefer not to answer to the gender question.
[d] Codes are according to the Australian Bureau of Statistics (Australian Statistical Geography Standard (ASGS) Edition 3, July 2021 - June 2026 | Australian Bureau of Statistics, 2024)
[e] *Postcode data is missing, invalid or could not be mapped to ABS datafiles. The Postcode question was not mandatory when included in the survey from February 2024*[f]
Prior screening history is a modelled variable- see supplementary figure 1
P-value indicate the statistical significance of differences observed among groups. Significant levels P < 0.05 are denoted in bold. P-trend calculated using Cochran–Armitage test.

**Supplementary Table 5A. Demographic and screening history factors associated with choosing self-collection.**

| | Total N (%) | Chose SC n (%) | Unadjusted OR (95% CI) | Adjusted^ OR (95% CI) | Adjusted^ OR (95%) Sensitivity Analysis[&] |
|---|---|---|---|---|---|
| **Total** | 1617 (100) | 803 (49.7) | | | |
| **Mean Age [SD]** | 43.5 [12.1] | 44.4 [12.1] | | | |
| **Age group (years)** | | | | | |
| 20-24 | 10 (0.6) | <5 (0.5) | 0.83 (0.23-2.99) | 1.68 (0.42-6.78) | 1.31 (0.33-5.23) |
| 25-34 | 434 (26.8) | 193 (24.0) | Ref. | Ref. | Ref. |
| 35-44 | 466 (28.8) | 229 (28.5) | 1.21 (0.93-1.57) | 1.03 (0.78-1.36) | 1.09 (0.83-1.44) |
| 45-54 | 380 (23.5) | 200 (24.9) | 1.39 (1.05-1.83) | 1.19 (0.89-1.59) | 1.14 (0.85-1.52) |
| 55-64 | 228 (14.1) | 127 (15.8) | **1.57 (1.14-2.17)** | 1.31 (0.93-1.85) | 1.27 (0.90-1.78) |
| Over 65 | 99 (6.1) | 50 (6.2) | 1.27 (0.82-1.97) | 1.02 (0.64-1.62) | 0.97 (0.62-1.54) |
| ***P-value*** | | | 0.082 | 0.573 | 0.792 |
| ***P-trend*** | | | **0.007** | 0.248 | 0.436 |
| **Gender** [b,c] | | | | | |
| Cis | 1,600 (98.9) | 794 (98.9) | Ref. | | |
| Trans | 16 (1.0) | 8 (1.0) | 1.02 (0.38-2.72) | | |
| Not stated | <5 (0.1) | <5 (0.1) | omitted. | | |
| ***P-value*** | | | 0.976 | | |
| **Intersex** [b] | | | | | |
| No | 1,600 (98.9) | 795 (99.0) | Ref. | | |
| Yes | 5 (0.3) | <5 (0.4) | 1.52 (0.25-9.11) | | |
| PNA/Unsure | 10 (0.6) | 5 (0.6) | 1.01 (0.29-3.51) | | |
| Missing Data | <5 (0.1) | 0 (0.0) | omitted. | | |
| ***P-value*** | | | 0.901 | | |
| **Sexuality** [b] | | | | | |
| Heterosexual | 1,431 (88.5) | 714 (88.9) | Ref. | | |
| Gay/Lesbian | 28 (1.7) | 13 (1.6) | 0.87 (0.41-1.84) | | |
| Bisexual | 113 (7.0) | 51 (6.4) | 0.83 (0.56-1.21) | | |
| I use a different term | 34 (2.1) | 19 (2.4) | 1.27 (0.64-2.52) | | |
| Not stated | 10 (0.6) | 5 (0.6) | 1.00 (0.29-3.48) | | |
| Missing Data | <5 (0.1) | <5 (0.1) | omitted. | | |
| ***P-value*** | | | 0.810 | | |
| **State** | | | | | |
| ACT | 50 (3.1) | 20 (2.5) | 0.63 (0.35-1.14) | | |
| NSW | 446 (27.6) | 230 (28.6) | Ref. | | |
| NT | 41 (2.5) | 20 (2.5) | 0.89 (0.47-1.70) | | |
| QLD | 295 (18.2) | 131 (16.3) | 0.75 (0.56-1.01) | | |
| SA | 90 (5.6) | 45 (5.6) | 0.94 (0.60-1.48) | | |
| TAS | 77 (4.8) | 42 (5.2) | 1.13 (0.69-1.83) | | |
| VIC | 430 (26.6) | 227 (28.3) | 1.05 (0.81-1.37) | | |
| WA | 188 (11.6) | 88 (11.0) | 0.83 (0.59-1.16) | | |
| ***P-value*** | | | 0.269 | | |
| **Remoteness of residence** [d,e] | | | | | |
| Major city | 979 (60.5) | 471 (58.7) | Ref. | Ref. | Ref. |
| Inner regional | 382 (23.6) | 214 (26.5) | **1.37 (1.08-1.74)** | **1.30 (1.01-1.67)** | **1.33 (1.04-1.70)** |
| Outer regional, remote, very remote | 159 (9.8) | 75 (9.3) | 0.96 (0.69-1.35) | 0.89 (0.62-1.28) | 0.96 (0.67-1.36) |
| Missing data | 97 (6.1) | 43 (5.5) | 0.85 (0.56-1.31) | 0.89 (0.56-1.36) | 0.91 (0.60-1.41) |
| ***P-value*** | | | **0.035** | 0.105 | 0.109 |
| **SEIFA** [d,e] | | | | | |
| 1 (most disadvantaged) | 161 (10.0) | 68 (8.5) | Ref. | | |
| 2 | 275 (17.0) | 148 (18.4) | **1.59 (1.08-2.36)** | | |

| | Total N (%) | Chose SC n (%) | Unadjusted OR (95% CI) | Adjusted^ OR (95% CI) | Adjusted^ OR (95%) Sensitivity Analysis[&] |
|---|---|---|---|---|---|
| 3 | 344 (21.3) | 183 (22.8) | **1.55 (1.07-2.27)** | | |
| 4 | 372 (23.0) | 183 (22.8) | 1.32 (0.91-1.92) | | |
| 5 (most advantaged) | 368 (22.8) | 178 (22.2) | 1.28 (0.88-1.86) | | |
| Missing Data | 97 (6.0) | 43 (5.4) | 1.09 (0.66-1.81) | | |
| ***P-value*** | | | 0.130 | | |
| **Country of birth** | | | | | |
| Australia | 1,348 (83.4) | 656 (81.7) | Ref. | | |
| Other | 265 (16.4) | 145 (18.1) | 1.27 (0.98-1.66) | | |
| Missing Data | <5 (0.2) | <5 (0.2) | 1.05 (0.15-7.51) | | |
| ***P-value*** | | | 0.198 | | |
| **Language spoken at home** | | | | | |
| English | 1,595 (98.6) | 790 (98.4) | Ref. | | |
| Language Other Than English | 21 (1.3) | 12 (1.5) | 1.36 (0.57-3.24) | | |
| I prefer not to answer | <5 (0.1) | <5 (0.1) | omitted. | | |
| ***P-value*** | | | 0.490 | | |
| **Duration of time in Australia** | | | | | |
| Australian born | 1,348 (83.4) | 656 (81.7) | Ref. | | |
| 0-4 years | 8 (0.5) | <5 (0.4) | 0.63 (0.15-2.66) | | |
| 5-9 years | 22 (1.4) | 11 (1.4) | 1.05 (0.45-2.45) | | |
| >10 years | 231 (14.3) | 126 (15.7) | 1.27 (0.96-1.68) | | |
| Missing Data | 8 (0.5) | 7 (0.9) | 7.38 (0.91-60.18) | | |
| ***P-value*** | | | 0.162 | | |
| **Highest education attained** | | | | | |
| Tertiary | 1,027 (63.5) | 518 (64.5) | Ref. | | |
| Tafe/Certificate | 388 (24.0) | 200 (24.9) | 1.05 (0.83-1.32) | | |
| School | 195 (12.1) | 84 (10.5) | 0.74 (0.55-1.01) | | |
| Other/PNA | 7 (0.4) | <5 (0.1) | 0.16 (0.02-1.37) | | |
| ***P-value*** | | | 0.076 | | |
| **Prior screening history** | | | | | |
| Regular screeners | 1,279 (79.1) | 581 (72.4) | Ref. | Ref. | |
| Not-Regular screeners | 338 (20.9) | 222 (27.6) | **2.30 (1.79-2.95)** | **2.31 (1.74-3.07)** | |
| ***P-value*** | | | **<0.001** | **<0.001** | |
| **Location of last CST** | | | | | |
| Doctors' clinic | 1,446 (89.4) | 728 (90.7) | Ref. | Ref. | Ref. |
| Women's/Sexual health | 111 (6.9) | 47 (5.9) | 0.72 (0.49-1.07) | 0.80 (0.53-1.21) | 0.76 (0.50-1.15) |
| Gynaecologist/Hospital | 44 (2.7) | 14 (1.7) | **0.46 (0.24-0.88)** | 0.63 (0.31-1.25) | 0.65 (0.33-1.29) |
| Other | 16 (1.0) | 14 (1.7) | **6.90 (1.56-30.49)** | 4.45 (0.98-20.10) | 5.18 (1.16-23.10) |
| ***P-value*** | | | **0.002** | 0.082 | **0.048** |
| **Prompt for last CST** | | | | | |
| Another gynaecological appointment/worried about symptom | 40 (2.5) | 12 (1.5) | **0.40 (0.20-0.81)** | **0.36 (0.17-0.76)** | **0.43 (0.21-0.89)** |
| Can't remember | <5 (0.2) | <5 (0.2) | 1.86 (0.17-20.69) | 2.06 (0.18-23.91) | 2.26 (0.20-25.56) |
| Follow-up previous abnormal screen | 27 (1.7) | <5 (0.5) | **0.16 (0.05-0.48)** | **0.17 (0.06-0.53)** | **0.19 (0.06-0.56)** |
| Health worker told me | 229 (14.2) | 148 (18.4) | **1.70 (1.20-2.40)** | 1.32 (0.92-1.91) | **1.69 (1.19-2.40)** |
| Other | 35 (2.2) | 23 (2.9) | 1.78 (0.86-3.70) | 1.05 (0.49-2.28) | 1.79 (0.86-3.75) |
| Remembered I was due | 185 (11.4) | 93 (11.6) | 0.94 (0.65-1.35) | 0.98 (0.68-1.42) | 0.93 (0.64-1.34) |
| Reminder from clinic | 225 (13.9) | 107 (13.3) | 0.84 (0.60-1.18) | 0.91 (0.64-1.29) | 0.83 (0.59-1.17) |
| Reminder letter from Register | 326 (20.2) | 169 (21.0) | Ref. | Ref. | Ref. |
| Saw something on media | 5 (0.3) | <5 (0.5) | 3.72 (0.41-33.61) | 2.43 (0.26-22.91) | 4.01 (0.44-36.48) |
| Multiple- excluding Gynaecological appointment/symptoms/follow-up | 403 (24.9) | 204 (25.4) | 0.95 (0.71-1.28) | 1.01 (0.75-1.36) | 1.00 (0.74-1.34) |
| Multiple-including Gynaecological appointment/symptoms/follow-up | 139 (8.6) | 37 (4.6) | **0.34 (0.22-0.52)** | **0.34 (0.22-0.54)** | 0.38 (0.24-0.59) |

| | Total N (%) | Chose SC n (%) | Unadjusted OR (95% CI) | Adjusted^ OR (95% CI) | Adjusted^ OR (95%) Sensitivity Analysis[&] |
|---|---|---|---|---|---|
| ***P-value*** | | | **<0.001** | **<0.001** | **<0.001** |

**Notes:** Abbreviations: CI, Confidence Interval; PNA, prefer not to answer; SEIFA, Socio-Economic Indexes for Areas; SC, self-collection, SD, Standard Deviation; SEIFA, Socio-Economic Indexes for Areas.

Total = Column percentages shown, chose SC = row percentages shown.

^Adjusted for; Age group, remoteness of residence, modelled prior screening history (removed for sensitivity analysis), location of last CST, and prompt for last CST.

b Question development and reporting are according to ACON indicators (Recommended Community Indicators, n.d.)

c Analysed and reported on in line with the ABS two-step method (Standard for Sex, Gender, Variations of Sex Characteristics and Sexual Orientation Variables, 2020 | Australian Bureau of Statistics, 2023), Not stated refers to participants who selected prefer not to answer to the gender question.

d Codes are according to the Australian Bureau of Statistics (Australian Statistical Geography Standard (ASGS) Edition 3, July 2021 - June 2026 | Australian Bureau of Statistics, 2024)

[e] Postcode data is missing, invalid or could not be mapped to ABS datafiles. The Postcode question was not mandatory when included in the survey from February 2024

[f] Prior screening history is a modelled variable- see supplementary figure 1

P-value indicate the statistical significance of differences observed among groups. Significant levels P < 0.05 are denoted in bold. P-trend calculated using Cochran–Armitage test.

&Sensitivity analysis was conducted on the adjusted regression model by removing the screening history variable to assess the robustness of the associations.

#### Supplementary Table 5B. Demographic and screening history factors associated with choosing self-collection - complete case analysis

| | Chose SC n (%) complete case | Unadjusted OR (95%) complete case | Adjusted OR (95%) Complete case | Adjusted^ OR (95% CI) – final regression* |
|---|---|---|---|---|
| **Total** | 753 | | | |
| **Mean Age [SD]** | 43.6 [12.1] | | | |
| **Age group (years)** | | | | |
| 20-24 | <5 (0.5) | 1.27 (0.31-5.16) | 2.20 (0.49-9.87) | 1.68 (0.42-6.78) |
| 25-34 | 176 (23.4) | Ref. | Ref. | Ref. |
| 35-44 | 220 (29.2) | 1.24 (0.95-1.63) | 1.08 (0.81-1.44) | 1.03 (0.78-1.37) |
| 45-54 | 184 (24.5) | **1.44 (1.08-1.92)** | 1.21 (0.89-1.65) | 1.19 (0.89-1.59) |
| 55-64 | 121 (16.1) | **1.60 (1.15-2.24)** | 1.34 (0.94-1.91) | 1.31 (0.93-1.85) |
| Over 65 | 48 (6.4) | 1.33 (0.85-2.08) | 1.08 (0.67-1.74) | 1.02 (0.64-1.62) |
| ***P-value*** | | 0.077 | 0.539 | 0.575 |
| ***P-trend*** | | **0.007** | 0.217 | 0.248 |
| **Gender** [b,c] | | | | |
| Cis | 744 (98.8) | Ref. | | |
| Trans | 8 (1.1) | 1.16 (0.42-3.20) | | |
| Not stated | <5 (0.1) | omitted | | |
| ***P-value*** | | 0.784 | | |
| **Intersex** [b] | | | | |
| No | 745 (98.9) | Ref. | | |
| Yes | <5 (0.4) | 1.51 (0.25-9.09) | | |
| PNA/Unsure | 5 (0.7) | 1.26 (0.34-4.72) | | |
| ***P-value*** | | 0.851 | | |
| **Sexuality** [b] | | | | |
| Heterosexual | 670 (90.0) | Ref. | | |
| Gay/Lesbian | 13 (1.7) | 0.93 (0.43-1.99) | | |
| Bisexual | 47 (6.2) | 0.83 (0.55-1.23) | | |
| I use a different term | 18 (1.9) | 1.50 (0.72-3.14) | | |
| Not stated | 5 (0.7) | 1.00 (0.29-3.47) | | |
| ***P-value*** | | 0.706 | | |
| **State** | | | | |
| ACT | 20 (2.7) | 0.72 (0.39-1.32) | | |
| NSW | 217 (28.9) | Ref. | | |
| NT | 19 (2.5) | 0.88 (0.46-1.70) | | |
| QLD | 122 (16.2) | **0.73 (0.54-0.99)** | | |
| SA | 44 (5.9) | 0.95 (0.60-1.51) | | |
| TAS | 39 (5.2) | 1.13 (0.68-1.88) | | |
| VIC | 210 (27.9) | 1.04 (0.79-1.37) | | |
| WA | 82 (10.9) | 0.82 (0.58-1.17) | | |
| ***P-value*** | | 0.336 | | |
| **Remoteness of residence** [d,e] | | | | |
| Major city | 466 (61.8) | Ref. | Ref. | Ref. |
| Inner regional | 213 (27.8) | **1.38 (1.09-1.76)** | 1.32 (1.03-1.70) | **1.30 (1.01-1.67)** |
| Outer regional, remote, very remote | 74 (9.8) | 0.96 (0.68-1.34) | 0.89 (0.62-1.28) | 0.89 (0.62-1.28) |
| Missing data | | | | 0.89 (0.56-1.36) |
| ***P-value*** | | **0.019** | 0.052 | 0.122 |
| **SEIFA** [d,e] | | | | |
| 1 (most disadvantaged) | 67 (8.9) | Ref. | | |
| 2 | 147 (19.5) | **1.61 (1.08-2.38)** | | |
| 3 | 181 (24.0) | **1.56 (1.07-2.28)** | | |

| | Chose SC n (%) complete case | Unadjusted OR (95%) complete case | Adjusted OR (95%) Complete case | Adjusted^ OR (95% CI) – final regression* |
|---|---|---|---|---|
| 4 | 180 (23.9) | 1.31 (0.90-1.93) | | |
| 5 (most advantaged) | 178 (23.7) | 1.31 (0.90-1.90) | | |
| ***P-value*** | | 0.121 | | |
| **Country of birth** | | | | |
| Australia | 621 (82.5) | Ref. | | |
| Other | 132 (17.5) | 1.18 (0.90-1.55) | | |
| ***P-value*** | | 0.243 | | |
| **Language spoken at home** | | | | |
| English | 742 (98.5) | Ref. | | |
| Language Other Than English | 10 (1.3) | 1.12 (0.45-2.78) | | |
| I prefer not to answer | 1 (0.1) | omitted. | | |
| ***P-value*** | | 0.804 | | |
| **Duration of time in Australia** | | | | |
| Australian born | 621 (82.5) | Ref. | | |
| 0-4 years | 3 (0.4) | 0.62 (0.15-2.61) | | |
| 5-9 years | 11 (1.5) | 1.04 (0.45-2.41) | | |
| >10 years | 118 (15.7) | 1.22 (0.91-1.63) | | |
| ***P-value*** | | 0.512 | | |
| **Highest education attained** | | | | |
| Tertiary | 487 (64.7) | Ref. | | |
| Tafe/Certificate | 189 (25.1) | 1.02 (0.80-1.30) | | |
| School | 76 (10.1) | **0.71 (0.51-0.98)** | | |
| Other/PNA | 1 (0.1) | 0.24 (0.03-2.18) | | |
| ***P-value*** | | 0.094 | | |
| **Prior screening history** | | | | |
| Regular screeners | 547 (72.6) | Ref. | Ref. | Ref. |
| Not-Regular screeners | 206 (27.4) | **2.27 (1.75-2.94)** | 2.31 (1.72-3.10) | **2.31 (1.74-3.07)** |
| ***P-value*** | | **<0.001** | **<0.001** | **<0.001** |
| **Location of last CST** | | | | |
| Doctors' clinic | 681 (90.4) | Ref. | Ref. | Ref. |
| Women's/Sexual health | 45 (6) | 0.71 (0.48-1.06) | 0.79 (0.51-1.20) | 0.80 (0.53-1.21) |
| Gynaecologist/Hospital | 13 (1.7) | **0.47 (0.24-0.92)** | 0.63 (0.31-1.29) | 0.63 (0.31-1.25) |
| Other | 14 (1.9) | **6.87 (1.55-30.28)** | 4.52 (1.00-20.43) | 4.45 (0.98-20.10) |
| ***P-value*** | | **0.003** | 0.082 | 0.082 |
| **Prompt for last CST** | | | | |
| Another gynaecological appointment/worried about symptom | 10 (1.3) | **0.37 (0.17-0.79)** | 0.31 (0.14-0.69) | **0.36 (0.17-0.76)** |
| Can't remember | <5 (0.3) | 1.83 (0.16-20.36) | 2.02 (0.17-23.52) | 2.06 (0.18-23.91) |
| Follow-up previous abnormal screen | <5 (0.5) | **0.18 (0.06-0.55)** | 0.19 (0.06-0.59) | **0.17 (0.06-0.53)** |
| Health worker told me | 139 (18.5) | **1.67 (1.16-2.39)** | 1.30 (0.09-1.89) | 1.32 (0.92-1.91) |
| Other | 22 (2.9) | 1.83 (0.86-3.90) | 1.10 (0.49-2.44) | 1.05 (0.49-2.28) |
| Remembered I was due | 85 (11.3) | 0.86 (0.60-1.25) | 0.89 (0.61-1.31) | 0.98 (0.68-1.42) |
| Reminder from clinic | 101 (13.4) | 0.85 (0.60-1.20) | 0.91 (0.64-1.30) | 0.91 (0.64-1.29) |
| Reminder letter from Register | 162 (21.5) | Ref. | Ref. | Ref. |
| Saw something on media | <5 (0.4) | 2.74 (0.28-26.64) | 1.94 (0.19-19.83) | 2.43 (0.26-22.91) |
| Multiple- excluding gynaecological appointment/symptoms/follow-up | 193 (25.6) | 0.96 (0.71-1.30) | 1.03 (0.75-1.40) | 1.01 (0.75-1.36) |
| Multiple-including gynaecological appointment/symptoms/follow-up | 32 (4.3) | **0.31 (0.20-0.49)** | **0.31 (0.19-0.51)** | **0.34 (0.22-0.54)** |
| ***P-value*** | | **<0.001** | **<0.001** | **<0.001** |

**Notes:** Abbreviations: CI, Confidence Interval; PNA, prefer not to answer; SEIFA, Socio-Economic Indexes for Areas; SC, self-collection, SD, Standard Deviation; SEIFA, Socio-Economic Indexes for Areas.
Total = Column percentages shown, chose SC = row percentages shown.
*Adjusted OR and 95% CI from regression analysis included from corresponding table a for easy comparison.
^Adjusted for; Age group, remoteness of residence, modelled prior screening history (removed for sensitivity analysis), location of last CST, and prompt for last CST.

[b] Question development and reporting are according to ACON indicators (Recommended Community Indicators, n.d.)
[c] Analysed and reported on in line with the ABS two-step method (Standard for Sex, Gender, Variations of Sex Characteristics and Sexual Orientation Variables, 2020 | Australian Bureau of Statistics, 2023), Not stated refers to participants who selected prefer not to answer to the gender question.
[d] Codes are according to the Australian Bureau of Statistics (Australian Statistical Geography Standard (ASGS) Edition 3, July 2021 - June 2026 | Australian Bureau of Statistics, 2024)
[e] *Postcode data is missing, invalid or could not be mapped to ABS datafiles. The Postcode question was not mandatory when included in the survey from February 2024*
[f] Prior screening history is a modelled variable- see supplementary figure 1
P-value indicate the statistical significance of differences observed among groups. Significant levels P < 0.05 are denoted in bold. P-trend calculated using Cochran–Armitage test.
[&]Sensitivity analysis was conducted on the adjusted regression model by removing the screening history variable to assess the robustness of the associations.

**Supplementary Table 6A. Demographic and screening history factors associated with using SC at last cervical screen.**

| | Total N (%) | Used SC n (%) | Unadjusted OR (95% CI) | Adjusted^ OR (95% CI) | Adjusted^ OR (95% CI) Sensitivity analysis& |
|---|---|---|---|---|---|
| **Total** | 4,656 | 969 (20.8) | | | |
| **Mean Age [SD]** | 42.7 [11.7] | 44.5 [12.0] | | | |
| **Age group (years)** | | | | | |
| 20-24 | 41 (0.9) | 5 (0.5) | 0.65 (0.25-1.67) | 1.08 (0.38-3.08) | 0.97 (0.34-2.77) |
| 25-34 | 1,291 (27.7) | 228 (23.5) | Ref. | Ref. | Ref. |
| 35-44 | 1,449 (31.1) | 283 (29.2) | 1.13 (0.93-1.37) | 0.97 (0.79-1.19) | 0.99 (0.81-1.22) |
| 45-54 | 1,040 (22.3) | 248 (25.6) | **1.46 (1.19-1.79)** | 1.14 (0.92-1.42) | 1.14 (0.92-1.42) |
| 55-64 | 616 (13.2) | 142 (14.7) | **1.40 (1.10-1.77)** | 1.03 (0.80-1.33) | 1.01 (0.78-1.30) |
| Over 65 | 219 (4.7) | 63 (6.5) | **1.88 (1.36-2.61)** | 1.30 (0.92-1.85) | 1.25 (0.88-1.78) |
| ***P-value*** | | | **<0.001** | 0.071 | 0.616 |
| ***P-trend*** | | | **<0.001** | 0.947 | 0.244 |
| **Gender [b,c]** | | | | | |
| Cis | 4,603 (98.9) | 960 (99.1) | Ref. | | |
| Trans | 49 (1.1) | 8 (0.8) | 0.74 (0.35-1.58) | | |
| Not stated | <5 (0.1) | <5 (0.1) | 1.26 (0.13-12.17) | | |
| ***P-value*** | | | 0.726 | | |
| **Intersex [b]** | | | | | |
| No | 4,611 (99.0) | 960 (99.1) | Ref. | | |
| Yes | 11 (0.2) | <5 (0.3) | 1.43 (0.38-5.39) | | |
| PNA/Unsure | 28 (0.6) | 6 (0.6) | 1.04 (0.42-2.57) | | |
| Missing Data | 6 (0.1) | 0 (0.0) | omitted | | |
| ***P-value*** | | | 0.869 | | |
| **Sexuality [b]** | | | | | |
| Heterosexual | 4,120 (88.5) | 858 (88.5) | Ref. | | |
| Gay/Lesbian | 63 (1.4) | 14 (1.4) | 1.09 (0.60-1.98) | | |
| Bisexual | 352 (7.6) | 67 (6.9) | 0.89 (0.68-1.18) | | |
| I use a different term | 76 (1.6) | 22 (2.3) | 1.55 (0.94-2.56) | | |
| Not stated | 44 (0.9) | 7 (0.7) | 0.72 (0.32-1.62) | | |
| Missing Data | <5 (0.0) | <5 (0.1) | omitted | | |
| ***P-value*** | | | 0.356 | | |
| **State** | | | | | |
| ACT | 153 (3.3) | 21 (2.2) | 0.67 (0.41-1.08) | 0.74 (0.45-1.21) | 0.73 (0.45-1.20) |
| NSW | 1,406 (30.2) | 271 (28.0) | Ref. | Ref. | Ref. |
| NT | 101 (2.2) | 23 (2.4) | 1.23 (0.76-2.00) | 1.52 (0.86-2.71) | 1.50 (0.84-2.66) |
| QLD | 935 (20.1) | 153 (15.8) | 0.82 (0.66-1.02) | 0.89 (0.71-1.13) | 0.89 (0.71-1.12) |
| SA | 269 (5.8) | 62 (6.4) | 1.25 (0.92-1.72) | **1.45 (1.04-2.01)** | **1.43 (1.03-1.98)** |
| TAS | 168 (3.6) | 54 (5.6) | **1.98 (1.40-2.81)** | **1.61 (1.09-2.37)** | **1.58 (1.08-2.32)** |
| VIC | 1,060 (22.8) | 272 (28.1) | **1.45 (1.19-1.75)** | **1.54 (1.26-1.89)** | **1.55 (1.27-1.90)** |
| WA | 564 (12.1) | 113 (11.7) | 1.05 (0.82-1.34) | 1.11 (0.86-1.44) | 1.11 (0.86-1.4) |
| ***P-value*** | | | **<0.001** | **<0.001** | **<0.001** |
| **Remoteness of residence [d,e]** | | | | | |
| Major city | 2,973 (63.9) | 570 (58.8) | Ref. | Ref. | Ref. |
| Inner regional | 971 (20.9) | 257 (26.4) | **1.52 (1.28-1.80)** | **1.38 (1.14-1.68)** | **1.40 (1.16-1.70)** |
| Outer regional, remote, very remote | 436 (9.4) | 92 (9.5) | 1.13 (0.88-1.44) | 1.05 (0.78-1.41) | 1.08 (0.80-1.45) |
| Missing data | 276 (5.9) | 50 (5.3) | 0.93 (0.68-1.28) | 0.93 (0.67-1.30) | 0.94 (0.67-1.32) |
| ***P-value*** | | | **<0.001** | **0.009** | **0.005** |
| **SEIFA [d,e]** | | | | | |
| 1 (most disadvantaged) | 459 (9.9) | 83 (8.6) | Ref. | | |
| 2 | 797 (17.1) | 185 (19.1) | **1.37 (1.03-1.83)** | | |
| 3 | 985 (21.2) | 225 (23.2) | **1.34 (1.01-1.78)** | | |
| 4 | 1,034 (22.2) | 217 (22.4) | 1.20 (0.91-1.59) | | |
| 5 (most advantaged) | 1,105 (23.7) | 209 (21.6) | 1.06 (0.80-1.40) | | |
| Missing Data | 276 (5.9) | 50 (5.2) | 1.00 (0.68-1.48) | | |
| ***P-value*** | | | 0.053 | | |
| **Country of birth** | | | | | |
| Australia | 3,837 (82.4) | 796 (82.1) | Ref. | | |
| Other | 812 (17.4) | 171 (17.6) | 1.02 (0.85-1.23) | | |
| Missing Data | 7 (0.2) | <5 (0.2) | 1.53 (0.30-7.89) | | |
| ***P-value*** | | | 0.864 | | |
| **Language spoken at home** | | | | | |
| English | 4,589 (98.6) | 953 (98.3) | Ref. | | |

| | Total N (%) | Used SC n (%) | Unadjusted OR (95% CI) | Adjusted^ OR (95% CI) | Adjusted^ OR (95% CI) Sensitivity analysis[&] |
|---|---|---|---|---|---|
| Language Other Than English | 64 (1.4) | 15 (1.5) | 1.17 (0.65-2.09) | | |
| PNA | <5 (0.0) | <5 (0.1) | 3.82 (0.24-61.05) | | |
| Missing Data | <5 (0.0) | 0 (0.0) | omitted. | | |
| ***P-value*** | | | 0.558 | | |
| **Duration of time in Australia** | | | | | |
| Australian born | 3,837 (82.4) | 796 (82.1) | Ref. | Ref. | Ref. |
| 0-4 years | 51 (1.1) | <5 (0.3) | 0.24 (0.07-0.77) | 0.28 (0.08-0.93) | **0.29 (0.09-0.96)** |
| 5-9 years | 96 (2.1) | 12 (1.2) | 0.55 (0.30-1.00) | 0.73 (0.38-1.39) | 0.76 (0.40-1.44) |
| >10 years | 649 (13.9) | 151 (15.6) | 1.16 (0.95-1.41) | 1.08 (0.87-1.33) | 1.09 (0.88-1.34) |
| Missing Data | 23 (0.5) | 7 (0.7) | 1.67 (0.69-4.08) | 1.58 (0.63-3.98) | 1.66 (0.67-4.16) |
| ***P-value*** | | | **0.01** | 0.150 | 0.162 |
| **Highest education attained** | | | | | |
| Tertiary | 2,896 (62.2) | 622 (64.2) | Ref. | | |
| Tafe/Certificate | 1,167 (25.1) | 243 (25.1) | 0.96 (0.81-1.14) | | |
| School | 572 (12.3) | 102 (10.5) | 0.79 (0.63-1.00) | | |
| Other/PNA | 21 (0.5) | <5 (0.2) | 0.38 (0.09-1.66) | | |
| ***P-value*** | | | 0.147 | | |
| **Prior screening history [f]** | | | | | |
| Regular screeners | 3,656 (78.5) | 694 (71.6) | Ref. | Ref. | |
| Not-Regular screeners | 1,000 (21.5) | 275 (28.4) | **1.62 (1.38-1.90)** | **1.60 (1.34-1.92)** | |
| ***P-value*** | | | **<0.001** | **<0.001** | |
| **Location of last CST** | | | | | |
| Doctors' clinic | 4,030 (86.6) | 881 (90.9) | Ref. | Ref. | Ref. |
| Women's/Sexual health | 245 (5.3) | 54 (5.6) | 1.01 (0.74-1.38) | 1.08 (0.78-1.51) | 1.08 (0.77-1.50) |
| Gynaecologist/Hospital | 356 (7.6) | 15 (1.5) | **0.16 (0.09-0.27)** | **0.21 (0.12-0.36)** | **0.21 (0.12-0.35)** |
| Other | 24 (0.5) | 19 (2.0) | **13.58 (5.06-36.48)** | **10.06 (3.65-27.75)** | **10.87 (3.95-29.95)** |
| Missing Data | <5 (0.0) | 0 (0.0) | omitted. | omitted. | omitted. |
| ***P-value*** | | | **<0.001** | **<0.001** | **<0.001** |
| **Prompt for last CST** | | | | | |
| Another Gynaecological appointment/Worried about symptom | 227 (4.9) | 16 (1.7) | **0.20 (0.12-0.34)** | **0.20 (0.11-0.34)** | **0.22 (0.13-0.38)** |
| Can't remember | 14 (0.3) | <5 (0.3) | 0.73 (0.20-2.63) | 0.70 (0.19-2.59) | 0.79 (0.21-2.91) |
| Follow-up previous abnormal screen | 191 (4.1) | <5 (0.4) | **0.06 (0.02-0.16)** | **0.07 (0.03-0.20)** | **0.07 (0.03-0.20**) |
| Health worker told me | 638 (13.7) | 181 (18.7) | 1.05 (0.83-1.33) | 0.96 (0.75-1.23) | 1.10 (0.86-1.40) |
| Other | 73 (1.6) | 28 (2.9) | **1.66 (1.01-2.73)** | 1.42 (0.83-2.42) | **1.77 (1.05-2.99)** |
| Remembered I was due | 531 (11.4) | 112 (11.6) | **0.71 (0.55-0.93)** | **0.71 (0.54-0.93)** | **0.72 (0.55-0.95)** |
| Reminder from clinic | 594 (12.8) | 122 (12.6) | **0.69 (0.53-0.89)** | **0.71 (0.55-0.93)** | **0.68 (0.52-0.88)** |
| Reminder letter from Register | 758 (16.3) | 207 (21.4) | Ref. | Ref. | Ref. |
| Saw something on media | 12 (0.3) | 6 (0.6) | 2.66 (0.85-8.35) | 2.33 (0.71-7.64) | 2.94 (0.91-9.57) |
| Multiple- excluding gynaecological appointment/symptoms/follow-up | 891 (19.1) | 238 (24.6) | 0.97 (0.78-1.21) | 0.98 (0.79-1.23) | 0.99 (0.79-1.24) |
| Multiple-including gynaecological appointment/symptoms/follow-up | 727 (15.6) | 52 (5.4) | **0.21 (0.15-0.28)** | **0.23 (0.17-0.32)** | **0.24 (0.17-0.34)** |
| ***P-value*** | | | **<0.001** | **<0.001** | **<0.001** |

**Notes;** Abbreviations: CI, Confidence Interval; PNA, prefer not to answer; SEIFA, Socio-Economic Indexes for Areas; SC, self-collection, SD, Standard Deviation; SEIFA, Socio-Economic Indexes for Areas.

Total = Column percentages shown, used SC = row percentages shown.

^Adjusted for; Age group, state, remoteness of residence, duration of time in Australia, modelled prior screening history (removed for sensitivity analysis), location of last CST, and prompt for last CST.

[b] Question development and reporting are according to ACON indicators (Recommended Community Indicators, n.d.)

[c] Analysed and reported on in line with the ABS two-step method (Standard for Sex, Gender, Variations of Sex Characteristics and Sexual Orientation Variables, 2020 | Australian Bureau of Statistics, 2023), Not stated refers to participants who selected prefer not to answer to the gender question.

[d] Codes are according to the Australian Bureau of Statistics (Australian Statistical Geography Standard (ASGS) Edition 3, July 2021 - June 2026 | Australian Bureau of Statistics, 2024)

[e] *Postcode data is missing, invalid or could not be mapped to ABS datafiles. The Postcode question was not mandatory when included in the survey from February 2024*

[f] Prior screening history is a modelled variable- see supplementary figure 1

P-value indicate the statistical significance of differences observed among groups. Significant levels P < 0.05 are denoted in bold. P-trend calculated using Cochran–Armitage test.

[&]Sensitivity analysis was conducted on the adjusted regression model by removing the screening history variable to assess the robustness of the associations.

#### Supplementary Table 6B. Demographic and screening history factors associated with using SC at last cervical screen – complete case analysis

| | Used SC n (%) complete case | Unadjusted OR (95%) complete case | Adjusted^ OR (95%) complete case | Adjusted^ OR (95% CI) – final regression* |
|---|---|---|---|---|
| **Total** | 912 (20.9) | | | |
| **Mean Age [SD]** | 44.6 [12.0] | | | |
| **Age group (years)** | | | | |
| 20-24 | <5 (0.4) | 0.60 (0.21 - 1.70) | 1.15 (0.38-3.46) | 1.08 (0.38-3.08) |
| 25-34 | 207 (22.7) | Ref. | Ref. | Ref. |
| 35-44 | 274 (30) | 1.18 (0.97 - 1.44) | 1.01 (0.82-1.25) | 0.97 (0.79-1.19) |
| 45-54 | 231 (25.4) | **1.50 (1.22 - 1.85)** | 1.17 (0.94-1.47) | 1.14 (0.92-1.42) |
| 55-64 | 135 (14.8) | **1.45 (1.13 - 1.85)** | 1.08 (0.83-1.40) | 1.03 (0.80-1.33) |
| Over 65 | 61 (6.7) | **2.02 (1.45 - 2.82)** | **1.43 (1.00-2.06)** | 1.30 (0.92-1.85) |
| ***P-value*** | | **<0.001** | 0.340 | 0.457 |
| ***P-trend*** | | **<0.001** | 0.070 | 0.155 |
| **Gender** [b,c] | | | | |
| Cis | 903 (99.0) | Ref. | | |
| Trans | 8 (0.9) | 0.84 (0.39 - 1.81) | | |
| Not stated | <5 (0.1) | 1.26 (0.13 - 12.09) | | |
| ***P-value*** | | 0.886 | | |
| **Intersex** [b] | | | | |
| No | 903 (99.0) | Ref. | | |
| Yes | <5 (0.3) | 1.62 (0.42 - 6.29) | | |
| PNA/Unsure | 6 (0.7) | 1.26 (0.50 - 3.19) | | |
| Missing Data | | | | |
| ***P-value*** | | 0.695 | | |
| **Sexuality** [b] | | | | |
| Heterosexual | 809 (88.7) | Ref. | | |
| Gay/Lesbian | 13 (1.4) | 1.04 (0.56 - 1.94) | | |
| Bisexual | 63 (6.9) | 0.90 (0.67 - 1.19) | | |
| I use a different term | 20 (2.2) | 1.54 (0.91 - 2.60) | | |
| Not stated | 7 (0.8) | 0.80 (0.35 - 1.81) | | |
| Missing Data | | | | |
| ***P-value*** | | 0.470 | | |
| **State** | | | | |
| ACT | 21 (2.3) | 0.70 (0.43 - 1.14) | 0.80 (0.48-1.32) | 0.74 (0.45-1.21) |
| NSW | 255 (28.0) | Ref. | Ref. | Ref. |
| NT | 22 (2.4) | 1.28 (0.78 - 2.12) | 1.58 (0.86-2.89) | 1.52 (0.86-2.71) |
| QLD | 144 (15.8) | 0.80 (0.64 – 1.00) | 0.88 (0.69-1.11) | 0.89 (0.71-1.13) |
| SA | 61 (6.7) | 1.28 (0.93 - 1.76) | **1.49 (1.06-2.09)** | 1.45 (1.04-2.01) |
| TAS | 51 (5.6) | **1.98 (1.38 - 2.85)** | **1.58 (1.06-2.35)** | 1.61 (1.09-2.37) |
| VIC | 251 (27.5) | **1.39 (1.14 - 1.70)** | **1.50 (1.22-1.85)** | 1.54 (1.26-1.89) |
| WA | 107 (11.7) | 1.01 (0.78 - 1.30) | 1.09 (0.83-1.43) | 1.11 (0.86-1.44) |
| ***P-value*** | | **<0.001** | **<0.001** | **<0.001** |
| **Remoteness of residence** [d,e] | | | | |
| Major city | 565 (62.0) | Ref. | Ref. | Ref. |
| Inner regional | 256 (28.1) | **1.52 (1.28 - 1.80)** | **1.38 (1.14-1.68)** | 1.38 (1.14-1.67) |
| Outer regional, remote, very remote | 91 (10.0) | 1.13 (0.88 - 1.45) | 1.05 (0.77-1.42) | 1.05 (0.78-1.41) |
| Missing data | | | | 0.93 (0.66-1.30) |
| ***P-value*** | | **<0.001** | **0.005** | **0.009** |
| **SEIFA** [d,e] | | | | |
| 1 (most disadvantaged) | 82 (9.0) | Ref. | | |
| 2 | 184 (20.2) | **1.38 (1.03 - 1.85)** | | |
| 3 | 223 (24.5) | **1.35 (1.02 - 1.79)** | | |
| 4 | 214 (23.5) | 1.20 (0.91 - 1.60) | | |
| 5 (most advantaged) | 209 (22.9) | 1.08 (0.81 - 1.43) | | |
| ***P-value*** | | 0.055 | | |
| **Country of birth** | | | | |
| Australia | 756 (82.9) | Ref. | | |
| Other | 156 (17.1) | 1.00 (0.82 - 1.22) | | |
| ***P-value*** | | 0.999 | | |
| **Language spoken at home** | | | | |
| English | 898 (98.5) | Ref. | | |
| Language Other Than English | 13 (1.4) | 1.12 (0.60 - 2.09) | | |
| PNA | <5 (0.1) | omitted | | |

| | Used SC n (%) complete case | Unadjusted OR (95%) complete case | Adjusted^ OR (95%) complete case | Adjusted^ OR (95% CI) – final regression* |
|---|---|---|---|---|
| ***P-value*** | | 0.725 | | |
| **Duration of time in Australia** | | | | |
| Australian born | 756 (82.9) | Ref. | Ref. | Ref. |
| 0-4 years | <5 (0.3) | **0.25 (0.08 - 0.81)** | 0.30 (0.09-1.01) | 0.28 (0.08-0.93) |
| 5-9 years | 11 (1.2) | **0.53 (0.28 - 0.99)** | 0.70 (0.36-1.38) | 0.73 (0.38-1.39) |
| >10 years | 142 (15.6) | 1.15 (0.94 - 1.41) | 1.07 (0.86-1.34) | 1.08 (0.87-1.33) |
| Missing data | | | | 1.58 (0.63-3.98) |
| ***P-value*** | | **0.009** | 0.153 | 0.150 |
| **Highest education attained** | | | | |
| Tertiary | 587 (64.4) | Ref. | | |
| Tafe/Certificate | 229 (25.1) | 0.94 (0.79 - 1.12) | | |
| School | 94 (10.3) | **0.79 (0.61 – 0.99)** | | |
| Other/PNA | <5 (0.2) | 0.52 (0.12 - 2.27) | | |
| ***P-value*** | | 0.187 | | |
| **Prior screening history** [f] | | | | |
| Regular screeners | 655 (71.8) | Ref. | Ref. | Ref. |
| Not-Regular screeners | 257 (28.2) | **1.59 (1.34 - 1.87)** | **1.61 (1.33-1.94)** | 1.60 (1.34-1.92) |
| ***P-value*** | | **<0.001** | **<0.001** | **<0.001** |
| **Location of last CST** | | | | |
| Doctors' clinic | 828 (90.8) | Ref. | Ref. | Ref. |
| Women's/Sexual health | 52 (5.7) | 1.02 (0.74 - 1.40) | 1.10 (0.79-1.55) | 1.08 (0.78-1.51) |
| Gynaecologist/Hospital | 14 (1.5) | **0.16 (0.09 - 0.27)** | **0.22 (0.12-0.38)** | 0.21 (0.12-0.36) |
| Other | 18 (2) | **16.01 (5.40 - 47.44)** | **11.50 (3.78-35.00)** | 10.06 (3.65-27.75) |
| Missing data | | | | omitted. |
| ***P-value*** | | **<0.001** | **<0.001** | **<0.001** |
| **Prompt for last CST** | | | | |
| Another Gynaecological appointment/Worried about symptom | 14 (1.5) | **0.19 (0.11-0.34)** | **0.18 (0.10-0.32)** | 0.20 (0.11-0.34) |
| Can't remember | <5 (0.3) | 0.78 (0.21-2.88) | 0.72 (0.19-2.72) | 0.70 (0.19-2.59) |
| Follow-up previous abnormal screen | <5 (0.4) | **0.06 (0.02-0.16)** | **0.07 (0.03-0.21)** | 0.07 (0.03-0.20) |
| Health worker told me | 170 (18.6) | 1.03 (0.81-1.32) | 0.94 (0.73-1.21) | 0.96 (0.75-1.23) |
| Other | 26 (2.9) | 1.58 (0.94-2.64) | 1.34 (0.77-2.32) | 1.42 (0.83-2.42) |
| Remembered I was due | 104 (11.4) | **0.69 (0.52-0.90)** | **0.68 (0.52-0.90)** | 0.71 (0.54-0.93) |
| Reminder from clinic | 116 (12.7) | **0.69 (0.53-0.90)** | **0.72 (0.55-0.94)** | 0.71 (0.55-0.93) |
| Reminder letter from Register | 198 (21.7) | Ref. | Ref. | Ref. |
| Saw something on media | 5 (0.5) | 2.61 (0.75-9.12) | 2.32 (0.63-8.57) | 2.33 (0.71-7.64) |
| Multiple- excluding Gynaecological appointment/symptoms/follow-up | 225 (24.7) | 0.96 (0.77-1.20) | 0.98 (0.78-1.24) | 0.98 (0.79-1.23) |
| Multiple-including Gynaecological appointment/symptoms/follow-up/ | 47 (5.2) | 0.19 (0.14-0.27) | **0.22 (0.15-0.31)** | 0.23 (0.17-0.32) |
| ***P-value*** | | **<0.001** | **<0.001** | **<0.001** |

**Notes:** Abbreviations: CI, Confidence Interval; PNA, prefer not to answer; SEIFA, Socio-Economic Indexes for Areas; SC, self-collection, SD, Standard Deviation; SEIFA, Socio-Economic Indexes for Areas.

Total = Column percentages shown, used SC = row percentages shown.

^Adjusted for; Age group, state, remoteness of residence, duration of time in Australia, modelled prior screening, location of last CST, and prompt for last CST.

*Adjusted OR and 95% CI from regression analysis included from corresponding table a for easy comparison.

[b] Question development and reporting are according to ACON indicators (Recommended Community Indicators, n.d.)

[c] Analysed and reported on in line with the ABS two-step method (Standard for Sex, Gender, Variations of Sex Characteristics and Sexual Orientation Variables, 2020 | Australian Bureau of Statistics, 2023), Not stated refers to participants who selected prefer not to answer to the gender question.

[d] Codes are according to the Australian Bureau of Statistics (Australian Statistical Geography Standard (ASGS) Edition 3, July 2021 - June 2026 | Australian Bureau of Statistics, 2024)

[e] Postcode data is missing, invalid or could not be mapped to ABS datafiles. The Postcode question was not mandatory when included in the survey from February 2024

[f] Prior screening history is a modelled variable- see supplementary figure 1

P-value indicate the statistical significance of differences observed among groups. Significant levels P < 0.05 are denoted in bold. P-trend calculated using Cochran–Armitage test.

#### Survey Tool

What is this survey about?

We are inviting people to complete this 10-15 minute survey about cervical screening.

You can do this survey if you are:

- Currently aged 24 to-74 years old
- Identify as a woman or have a cervix
- Live in Australia

This survey asks about your thoughts and feelings about cervical screening. It does not matter whether you have ever had a cervical screening test or not.

The survey is part of two research projects.

The first project is called *Supporting Choice* and is being led by:

- Dr Claire Nightingale at the University of Melbourne
- o Claire Bavor is involved in this project as part of her studies for the degree of Doctor of Philosophy at The University of Melbourne.
- Associate Professor Megan Smith at The Daffodil Centre (a joint venture between the University of Sydney and Cancer Council NSW).

o Chloe Jennett is involved in this project as part of her studies for the degree of Doctor of Philosophy at The University of Sydney.

The second project is called *Screen Your Way* and is being led by:

- Associate Professor Lisa Whop, a Gumulgal person of Mabuiag Island (Torres Strait Islands) at The Australian National University.

o Emily Phillips is an emerging Aboriginal researcher. She is involved in this project as part of her degree of Master of Epidemiology, at The Australian National University.

Where can I read more about the survey?

You can find out more about the survey by clicking here: <Link to participant information sheet>

You can contact the researchers or the ethics committee if you have any questions. Their details are on the <Link to participant information sheet>

Where do I go if I need help after doing this survey?

If you find this survey to be upsetting at any point, you may contact:

- Lifeline number: 13 11 14
- 13 YARN number: 13 92 76 which is staffed by Aboriginal and Torres Strait Islander peoples.
- QLife number: 1800 184 527, an anonymous and free LGBTIQ+ peer support and referral, available 3pm to midnight, every day.

For any other questions about cancer, you can call the

- Cancer Council Helpline: 13 11 20

What if I want to stop?

To stop the survey at any time, you can click the ‘exit’ button or close your internet browser. You can also re-open the survey later. Your responses will be recorded once you begin the survey.

What do I do now?

Please read the information below before starting the survey:

1. I understand that this research is about cervical screening in Australia
2. It is my own choice to take part in this study
3. I can stop the survey at any point, without a reason for stopping, and that this will not impact how I am treated or any access to services
4. Taking part in this research project requires me to submit an online survey
5. I am aware of the potential benefits and risks of taking part in this research project and they have been explained to me
6. Participation in this research project is voluntary, however, I know that I will not be paid for my participation, but that I may opt-in to a prize draw to win 1 of 24, $100 prepaid Visa gift vouchers
7. The results of this research may be published in a public domain or other forum, but my personal information and data will not be able to be identified.
8. All information gathered in this research project is confidential and will be kept secure for up to 15 years from the date of publication of research articles.
9. This research project is a collaboration between Melbourne School of Population and Global Health, The Daffodil Centre (a joint venture of Cancer Council NSW and the University of Sydney), and The Australian National University.

You can download a copy of the consent form to keep here: <link to consent form>

Q1.1 Would you like to take part in this survey?

- ○ I agree to take part in this survey
- ○ I do not agree, and wish to exit this survey

Q1.2 Please click the box below to help us prevent spam responses.
(captcha)

Q1.3 Where did you find out about this survey? (Please be specific to help us prevent spam responses).

_______________

Part 1. About you

All participants

Q2.1 Are you of Aboriginal or Torres Strait Islander origin?

- ○ Yes, Aboriginal
- ○ Yes, Torres Strait Islander

- Yes, both Aboriginal and Torres Strait Islander
- Neither Aboriginal or Torres Strait Islander

Eligibility criteria (survey ends for those not eligible):

Q2.2 How old are you?

________________________________________________________________

Q2.3 What was your sex recorded at birth?

Note that a separate question on gender is also asked in the survey.

- Female
- Male
- Another term (please specify) ________________________________________
- I prefer not to answer

Q2.4 Do you have a cervix?

The cervix is the lower part of your uterus (or womb) which connects your uterus and vagina. If your sex recorded at birth was female, you were born with a womb and cervix.

The main reason for not having a cervix is if it has been removed by surgery. This is called a hysterectomy. You may still have a cervix if you have had a partial hysterectomy, where only the uterus (womb) has been removed by surgery, leaving the cervix.

- Yes
- No
- Unsure
- I prefer not to answer

All participants- demographics

Q2.5 How do you describe your gender?

Gender refers to current gender, which may be different to your sex recorded at birth and may be different to what is on legal documents.

- Woman or Female
- Man or Male
- Non-binary
- I use a different term (please specify) ________________________________________
- I prefer not to answer

Q2.6 How do you describe your sexual orientation?

- Straight (Heterosexual)
- Gay or lesbian
- Bisexual
- I use a different term (please specify) ________________________________________
- Do not know
- I prefer not to answer

Q2.7 Were you born with a variation of sex characteristics (sometimes called 'intersex')?

- Yes
- No
- I do not know
- I prefer not to answer

Q2.8 What is the highest level of education and training you have completed?

- Did not go to school
- Primary school
- Some high school (i.e Year 7 to Year 11, Form 1 to Form 5)
- Completed high school (i.e. Year 12, Form 6, HSC or equivalent, International Baccalaureate)
- TAFE, Trade Certificate or Diploma but did not complete Year 12 at secondary school
- TAFE or Trade Certificate or Diploma and also completed Year 12 at secondary school
- University, or some other Tertiary Institute degree, including post university (i.e. postgraduate diploma, Master's degree, PhD)
- Other (please specify) ______________________________
- I prefer not to answer

Q2.9a What is your postcode?
(Free text entry, response = optional, not mandatory)

Q2.9b What Australian state or territory do you live in?

- Australian Capital Territory
- New South Wales
- Northern Territory
- Queensland
- South Australia
- Tasmania
- Victoria
- Western Australia

Q2.10 What is your country of birth?

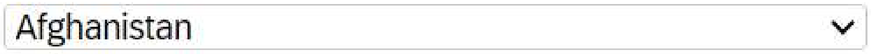

Q2.11 If you were born overseas, how long have you been living in Australia?

- Less than a year
- More than a year (please specify) ______________________________
- Unsure/do not know
- I prefer not to answer

Q2.12 What is the main language you speak at home?

- English
- Aboriginal language
- Torres Strait Islander language (including Creole, Pidgin, Yumplatok)
- Greek
- Italian
- Mandarin
- Cantonese
- Vietnamese
- Arabic
- Punjabi
- Hindi
- Tagalog

- Other (please specify) ___________________________________________
- I prefer not to answer

Q1.4 is hidden using java script, and can only be completed by spam/fraudulent accounts.

Q1.4 Are you a human?

Yes

#### Part 2. Taking part in cervical screening

All participants

Q3.1 Do you remember when your last cervical screening test was?
This might have been called a 'HPV test', 'Pap test' or 'Pap Smear'

- Within the last year (on or after July 2022)
- More than a year ago (Before July 2022)
- I cannot remember when my last cervical screening test was
- I have never had a cervical screening test
- I have never heard of a cervical screening test
- I prefer not to answer

Participants previously screened (Q3.1: Within the last year, more than a year ago):

Q3.2 Where did you go for your most recent cervical screening test?

- A doctor's clinic
- A community health centre
- A women's health centre
- A family planning clinic
- A sexual health clinic
- An Aboriginal Medical Service or Aboriginal Community Controlled Health service
- A gynaecologist
- I prefer not to answer
- Other, please specify:

Q3.3 How often do you attend cervical screening?

- I screen around about the time I am due
- I know that sometimes I leave it for too long between screens
- I have only had one screening test before
- I prefer not to answer

Q3.4 Why did you get your last cervical screening test?
You can select more than 1 option from the list below.

- I remembered I was due for a cervical screening test
- I went to the healthcare provider for another reason and they reminded me
- The health worker told me I was due
- I got a reminder letter, email, SMS (or similar) from my usual clinic
- I got a reminder letter from the National Cancer Screening Register/Pap Test register
- I wanted to follow-up after a previous abnormal cervical screening test
- I was worried about a symptom or health problem
- I saw something in the media (television, radio, social media)
- Other (please specify) ___________________________________________
- I cannot remember

- I prefer not to answer

Participants not previously screened (Q3.1: Cannot remember, never had a CST, never heard of a CST, prefer not to answer):

Q3.5 Can you tell me your reasons for not doing a cervical screening test?
You can select more than 1 option from the list below.

- I do not have time
- I forgot about it
- I cannot get to the healthcare clinic
- It is too expensive
- There is not a healthcare provider near me who can do cervical screening tests
- I am scared of it
- I am embarrassed/ashamed
- I am worried it would be painful
- I need more information about cervical screening before deciding
- I do not want to have a healthcare provider touch me
- I am worried I might be told I have cancer
- I do not think I am at risk of cervical cancer/no family history
- I have no symptoms
- I am not currently sexually active
- I have never been sexually active
- I have had the Human Papillomavirus (HPV) vaccine
- I was not aware I should have cervical screening
- Other (please specify) ________________________________________________
- I do not know why
- I prefer not to answer

All participants:

Q3.6 Have you ever heard of 'self-collection', 'self-sampling', or 'self-testing' as an option for cervical screening?

- Yes
- No
- Unsure
- I prefer not to answer

Heard of self-collection (Q3.6: yes):

Q3.7 How did you first hear about self-collection?
You can select more than 1 option from the list below.

- Family or friend
- Healthcare provider told me about it
- A letter or phone call from my healthcare clinic
- Letter from the National Cancer Screening Register
- Saw something in the media (television, radio, online news, print news)
- Social media
- Website
- Campaign from an organisation such as Cancer Council, or the Government or ACON
- Other (please specify) ________________________________________________
- Unsure/cannot remember

All participants:

About cervical screening

Please read the following information about cervical screening.

At the end of 2017 Australia's National Cervical Screening Program changed from using the Pap test, also known as a 'Pap smear', to a new and better, cervical screening test that looks for human papillomavirus (HPV). HPV is a common virus that causes almost all cervical cancers. The newer cervical screening test for HPV is better at preventing cervical cancer than the Pap test, and also allows for samples to be collected in different ways.

Since July 2022, all people attending cervical screening can choose how they want their sample taken:

**Option 1** is to take your own sample from the vagina using a soft swab which looks like a long cotton bud. This screening method is called 'self-collection'.

A healthcare provider can also help the person take their own sample, without a speculum, if the person would like them to.

**This is a self-collection soft swab**.

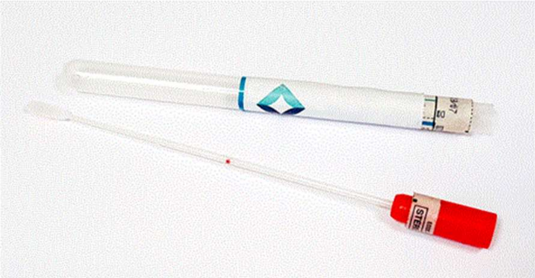

*Image from: The Australian Centre for the Prevention of Cervical Cancer.*

**Option 2** is to have a healthcare provider, like a doctor, nurse, or Aboriginal Health Practitioner, use a speculum. A speculum is a medical device that a healthcare professional uses to open the vagina so they can see the cervix and collect a sample from the cervix. This is the same way the Pap Test or Pap Smear was taken. In this survey, we will call this a 'sample collected by a healthcare provider'.

**This is a speculum. It can be made from metal or plastic.**

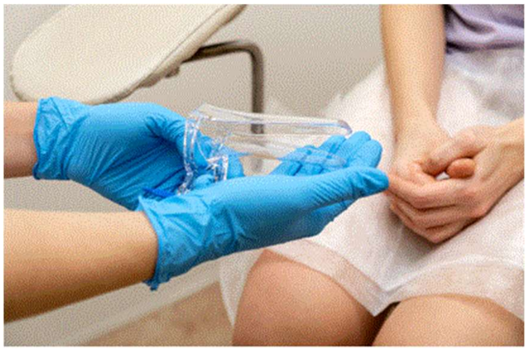

Image from: ShutterStock

**Both of these methods look for HPV and are available to women and people with a cervix for cervical screening.**

**Part 3. Your cervical screening options**

Participants recently screened (Q3.1: Within the last year)

Q4.1 For your last cervical screening test, were you offered the choice between self-collection or having a sample collected by the healthcare provider using a speculum?

- Yes, I was offered a choice
- No, I was not offered a choice
- I cannot remember
- I prefer not to answer

Q4.2 How did you do your last cervical screening test?

- I 'used self-collection' in the clinic
- I 'used self-collection' at home or somewhere else
- A healthcare provider helped me to collect my own sample (without a speculum)
- A healthcare provider collected the sample using a speculum
- I cannot remember
- I prefer not to answer

**Participants recently screened with self-collection (Q4.2: I used SC in the clinic, I used SC at home):**

Q4.3 Can you tell me your reasons for choosing to use self-collection?
You can select more than 1 option from the list below.

- It is less embarrassing
- It is not as scary
- It helps me feel in control of my body
- It is less painful
- I had a bad experience in the past when a sample was collected by a healthcare provider using a speculum
- A friend/family member had a good experience with self-collection
- It is just as accurate as a sample collected by a healthcare provider
- It is more convenient
- My healthcare provider suggested it as an option
- The information I received at my cervical screening appointment made it look like a good option
- Other (please specify) ______________________________________________
- Unsure/cannot remember
- I prefer not to answer

Q4.4 Were you given the following information about self-collection during your last visit for cervical screening?

| | Yes | No | Unsure/cannot remember |
|---|---|---|---|
| That I might need to come back for another test with a healthcare provider if HPV was found | ○ | ○ | ○ |
| That I could take the self-collection swab away and do it myself somewhere else | ○ | ○ | ○ |
| That if I needed to have a follow up test, I might need to have a sample collected by a healthcare provider | ○ | ○ | ○ |
| How I would get my test results (e.g SMS, phone call, letter) | ○ | ○ | ○ |

| | | | |
|---|---|---|---|
| That self-collection was as accurate as a sample collected by a healthcare provider | | | |
| I had enough information to help me decide between self-collection and having a sample collected by a healthcare provider | ○ | ○ | ○ |
| That I could ask for help with collecting the sample if I didn't feel comfortable | | | |

Q4.5 How much do you agree or disagree with the following statements?

| | Agree | Neither disagree nor agree | Disagree |
|---|---|---|---|
| I understood how to collect my own sample | ○ | ○ | ○ |
| I was confident I could do the test correctly | ○ | ○ | ○ |
| The information I was given was easy to understand | ○ | ○ | ○ |
| I was confident the test was accurate | | | |
| It was easy for me to collect the sample | ○ | ○ | ○ |
| I was worried about having to return for a follow up test | ○ | ○ | ○ |
| | ○ | ○ | ○ |
| | ○ | ○ | ○ |

Q4.6 If self-collection was **not** an option, how likely is it that you would have had cervical screening?

- I would not have had cervical screening without self-collection.
- I might have had cervical screening without self-collection.
- I would have had cervical screening anyway

Participants recently screened with clinician-collection (Q4.2: A healthcare provider collected the sample using a speculum):

Q4.7 Can you tell me your reasons for choosing to have a sample collected by the healthcare provider using a speculum?
You can select more than 1 option from the list below.

- I was not told about the option to use self-collection
- I have always had it done by a healthcare provider
- I wanted my healthcare provider to have a look
- My healthcare provider told me they did not offer self-collection
- I am not eligible for self-collection
- I needed more information on self-collection
- I did not think I would take the self-collected sample properly
- I did not think self-collection was as accurate
- I thought I would need to return for another test anyway if I 'used self-collection'

- Other reason, please specify ________________________________________
- I do not know why
- I prefer not to answer

Participants recently screened with clinician-assisted self-collection (Q4.2: A healthcare provider helped me to collect my own sample (without a speculum)):

Q4.8 Can you tell me your reasons for choosing to have your sample collected by a healthcare provider using a self-collection swab?
You can select more than 1 option from the list below.

- It's less embarrassing
- I thought that the healthcare provider could collect a better sample than I could
- I want my healthcare provider to have a look, even without a speculum, and make sure that everything is ok
- I cannot reach, or it is not comfortable for me to reach
- I do not like to touch myself there
- Other (please specify) ________________________________________
- Unsure/do not know
- I prefer not to answer

Participants previously screened (Q3.1: Within the last year, more than a year ago):

Q4.9 How would you prefer to do your next cervical screening test?

- Collected by a healthcare provider using a speculum
- Collected by a healthcare provider using a self-collection swab (but no speculum)
- Self-collection
- I would be okay with either
- I am not planning to screen
- I do not know
- I prefer not to answer

Participants not previously screened (Q3.1: Cannot remember, never had a CST, never heard of a CST, prefer not to answer):

Q4.10 Do you think you are more likely to take part in cervical screening because self-collection is available?

- Yes, I am more likely to have a cervical screening test
- No, I still think I would not have a cervical screening test
- It does not affect my decision to have a cervical screening test or not
- I am not sure
- I prefer not to answer

All participants:

Q4.11 How important are the following things to you when it comes to cervical screening?

| | Very important | Important | Not important | Not sure or not applicable |
|---|---|---|---|---|
| Having the choice over how I can do cervical screening | ○ | ○ | ○ | ○ |

| | | | | |
|---|---|---|---|---|
| Being able to talk with my healthcare provider about cervical screening | ○ | ○ | ○ | ○ |
| Having simple and clear information on cervical screening | ○ | ○ | ○ | ○ |
| Having my healthcare provider explains self-collection to me | ○ | ○ | ○ | ○ |
| Having the healthcare provider available if I needed help doing self-collection | ○ | ○ | ○ | ○ |
| | ○ | ○ | ○ | ○ |
| Feeling safe and comfortable within the clinic | ○ | ○ | ○ | ○ |
| Feeling safe and comfortable with the healthcare provider | ○ | ○ | ○ | ○ |
| Having the choice of healthcare provider (for example, a doctor, nurse of health worker) | ○ | ○ | ○ | ○ |
| Having a female healthcare provider | ○ | ○ | ○ | ○ |
| Having flexible options for how I can access cervical screening (for example, in the clinic or at home) | ○ | ○ | ○ | ○ |

Q4.12 Who would you prefer to talk with about cervical screening?

- My usual doctor (male or female)
- A female doctor
- Nurse
- Midwife
- Health worker
- Aboriginal health worker/practitioner
- Other, please specify:

______________________________

- I do not mind
- I prefer not to answer

Part 4. Taking part in cervical screening in the future

All participants:

The following questions are about self-collection. Even if you would prefer a healthcare provider to collect the sample using a speculum for your cervical screening test, we would like to know more about how you would prefer to pick up a self-collection swab, collect the sample, and return the swab.

Q5.1 Where would you most prefer to do self-collection (collect the sample yourself)?

- At my healthcare provider's clinic during an appointment
- At the place I picked up the swab from (for example, doctor's office, a pathology pick-up point)
- At home
- Somewhere else, please specify:
- I have no preference

Q5.2 How would you prefer to return the self-collection swab for it to be tested?

- o By mail with a prepaid envelope
- o Drop it off at my healthcare provider's clinic
- o Drop it off at a pathology pick-up point (for example, where you go to get a blood test)
- o Somewhere else, please specify:
- o I have no preference

Q5.3 How likely would you be to take part in cervical screening on time compared to now if the following options for self-collection were available?

| | More likely | Same | Less likely |
|---|---|---|---|
| Receiving the self-collection swab in the post after a telehealth appointment (phone or video call with your healthcare provider) | ○ | ○ | ○ |
| Collecting the self-collection swab from the doctor's office or pathology pick-up point after a telehealth appointment | ○ | ○ | ○ |
| Collecting the self-collection swab from my doctor's office/medical service without an appointment | ○ | ○ | ○ |
| Collecting the self-collection swab from a health worker during a routine health assessment | ○ | ○ | ○ |
| Picking up a self-collection swab from my Aboriginal Medical Service | ○ | ○ | ○ |
| Picking up a self-collection swab from a chemist/pharmacy | ○ | ○ | ○ |
| Ordering a self-collection swab online/over the phone when I am due for cervical screening | ○ | ○ | ○ |
| Automatically receiving a self-collection swab in the mail when I am due for screening | ○ | ○ | ○ |
| Picking up a self-collection swab at community group events | ○ | ○ | ○ |
| Picking up a self-collection swab when I attend breast screening (when you are aged 50-74) | ○ | ○ | ○ |
| Getting a self-collection swab when I receive my bowel screening test kit (when you are aged 50-74) | ○ | ○ | ○ |

Q5.4 How would you most prefer to get the swab for self-collection?

- o Receiving the self-collection swab in the post after a telehealth appointment (phone or video call with your healthcare provider)
- o Collecting the self-collection swab from the doctor's office or pathology pick-up point after a telehealth appointment
- o Collecting the self-collection swab from my doctor's office/medical service without an appointment
- o Collecting the self-collection swab from a health worker during a routine health assessment
- o Picking up a self-collection swab from my Aboriginal Medical Service

- Picking up a self-collection swab from a chemist/pharmacy
- Ordering a self-collection swab online/over the phone when I am due for cervical screening
- Automatically receiving a self-collection swab in the mail when I am due for screening
- Picking up a self-collection swab at community group events
- Picking up a self-collection swab when I attend breast screening (when you are aged 50-74)
- Getting a self-collection swab when I receive my bowel screening test kit (when you are aged 50-74)
- During a face-to-face appointment with my healthcare provider
- Other, please specify:

Q5.5 Why is this your most preferred option? You can select more than 1 option from the list below:

- It is closer to home
- I can get it outside of work hours
- I can bring my family or children with me
- It is more convenient
- It is less embarrassing
- It is less expensive
- I prefer to talk to a healthcare provider about cervical screening
- I have a trusted healthcare provider
- I don’t have a trusted healthcare provider
- I am not sure
- Other, please specify: ______________________________________________
- Prefer not to answer

Part 5. End of survey

Q6.1 Would you like to enter the prize draw for a $100 prepaid Visa gift card? You will be asked to provide your email address.

- Yes
- No

Q6.2 Would you like to receive a copy of the survey results? You will be asked to provide your email address.

- Yes
- No

Q1.5 Where did you find out about this survey? (We are asking this a second time to help identify spam responses)

________________

Q6.3 Please enter your email address so the researchers can contact you.
Your email address will only be used to let you know if you have won a voucher or send you a copy of the survey results. It will not be used to identify you or your answers.

___________

_________________________SUBMIT SURVEY_____________________

Thank you for participating in this survey!

If you would like to find out more about cervical screening and self-collection, you can speak to any healthcare provider, like a doctor, nurse, or Aboriginal Health Worker.

**You can get cervical screening at many different places including:**

- a doctor's clinic
- a community health centre
- a women's health centre
- a family planning clinic
- a sexual health clinic
- an Aboriginal Medical Service or Aboriginal Community Controlled Health Service.

**If you do not have a preferred healthcare provider, these websites can help you find one:**

Service directory: https://www.healthdirect.gov.au/australian-health-services

Get Papped: https://get-papped.com/pages/practitioner-directory

**You can find out more about cervical screening on these websites:**

National Cervical Screening Program: https://www.health.gov.au/our-work/national-cervical-screening-program/getting-a-cervical-screening-test

Cancer Council Australia: https://www.cancer.org.au/cervicalscreening

#### Survey Logic Flow Chart

<table>
<tr><td colspan="4">Part 1. About you</td></tr>
<tr><td colspan="4">All participants</td></tr>
<tr><td colspan="4">Q2.1-Q2.12 Demographics</td></tr>
<tr><td colspan="4">Part 2. Taking part in cervical screening</td></tr>
<tr><td colspan="4">All participants</td></tr>
<tr><td colspan="4">Q3.1 Do you remember when your last cervical screening test was?</td></tr>
<tr><td colspan="3">Previously screened (Q3.1: Within the last year, more than a year ago)</td><td>Never screened (Q3.1: cannot remember, never had a CST, never heard of a CST, prefer not to answer)</td></tr>
<tr><td colspan="3">Q3.2 Where did you go for your most recent cervical screening?</td><td rowspan="3">Q3.5 Can you tell me your reasons for not doing a cervical screening test?</td></tr>
<tr><td colspan="3">Q3.3 How often do you attend cervical screening/have a ‘Pap test’?</td></tr>
<tr><td colspan="3">Q3.4 Why did you get your last Cervical Screening Test or Pap Test?</td></tr>
<tr><td colspan="4">All participants</td></tr>
<tr><td colspan="4">Q3.6 Have you ever heard of 'self-collection', 'self-sampling', or 'self-testing' as an option for cervical screening?</td></tr>
<tr><td colspan="3">Heard of cervical screening (Q3.6: Yes)</td><td>Not heard of cervical screening (Q3.6: No, sure, prefer not to answer)</td></tr>
<tr><td colspan="3">Q3.7 How did you hear about the self-collection testing option?</td><td>-</td></tr>
<tr><td colspan="4">All participants: About cervical screening</td></tr>
<tr><td colspan="4">Part 3. Your cervical screening options</td></tr>
<tr><td colspan="3">Recently screened (Q3.1 Within the last year)</td><td>Never screened (Q3.1 Cannot remember, never had a CST, never heard of a CST, prefer not to answer)</td></tr>
<tr><td colspan="3">Q4.1 For your last cervical screening test, were you offered the choice between self-collection or having a sample collected by the healthcare provider using a speculum?</td><td rowspan="6">Q4.10 Do you think you are more likely to take part in cervical screening because self-collection is available?</td></tr>
<tr><td colspan="3">Q4.2 How did you do your last cervical screening test?</td></tr>
<tr><td>Self-collection (Q4.2 SC in the clinic, at home)</td><td>Clinician-collection (Q4.2 healthcare provider)</td><td>Clinician-assisted self-collection (Q4.2 a healthcare provider collected the sample without a speculum)</td></tr>
<tr><td>Q4.3 Can you tell me your reasons for choosing to use self-collection?</td><td rowspan="3">Q4.7 Can you tell me your reasons for choosing to have a sample collected by the healthcare provider</td><td rowspan="3">Q4.8 Can you tell me your reasons for choosing to have your sample collected by a healthcare provider using a self-collection swab?</td></tr>
<tr><td>Q4.4 Were you given the following information about self-collection during your last visit for cervical screening?</td></tr>
<tr><td>Q4.5 How much do you agree or disagree with</td></tr>
</table>

| | |
|---|---|
| the following statements? | using a speculum? |
| Q4.6 If self-collection was not an option, how likely is it that you would have had cervical screening? | |
| Participants previously screened (Q3.1: Within the last year, more than a year ago): | |
| Q4.9 How would you prefer to do your next cervical screening test? | |
| All participants | |
| 4.11 How important are the following things to you when it comes to cervical screening? | |
| 4.13 Who would you prefer to talk with about cervical screening? | |
| Part 4. Taking part in cervical screening in the future | |
| Q5.1 Where would you most prefer to do self-collection (collect the sample yourself)? | |
| Q5.2 How would you prefer to return the self-collection swab for it to be tested? | |
| Q5.3 How likely would you be to take part in cervical screening on time compared to now if the following options for self-collection were available? | |
| Q5.4 How would you most prefer to get the swab for self-collection? | |
| Q5.5 Why is this your most preferred option? You can select more than 1 option from the list below: | |
| Part 5. End of survey | |
| Q6.1 Would you like to enter the prize draw for a $100 prepaid Visa gift card? You will be asked to provide your email address. | |
| Q6.2 Would you like to receive a copy of the survey results? You will be asked to provide your email address. | |
| Q6.3 Please enter your email address so the researchers can contact you. | |
| Submit survey | |
